# Population genomics of four-dimensional cardiac motion reveals non-myocyte regulatory programmes

**DOI:** 10.1101/2025.11.22.25340784

**Authors:** Soodeh Kalaie, Nirmal Vadgama, Jin Zheng, Lara Curran, Kathryn A McGurk, Yue Qin, Mattia Corianò, Khaled Rjoob, Parisa Gifani, Qingjie Meng, Michela Noseda, Wenjia Bai, Christian Bender, Declan P O’Regan

**Affiliations:** MRC Laboratory of Medical Sciences, Imperial College London, Hammersmith Hospital Campus, London, UK; National Heart and Lung Institute, Imperial College London, London, UK; Department of Cardiac, Thoracic, Vascular Science and Public Health, University of Padova, Padova, Italy; Department of Computing, Imperial College London, London, UK; School of Computer Science, University of Birmingham, Birmingham, UK; Department of Brain Sciences, Imperial College London, London, UK; Bayer AG, Research & Development, Pharmaceuticals, Wuppertal, Germany; British Heart Foundation Centre of Research Excellence, Imperial College London, London, UK

## Abstract

Genome-wide studies of cardiac structure and function have relied on global imaging metrics that fail to capture the full complexity of myocardial behaviour, leaving the genetic and environmental architecture of motion largely unexplored. Here we show, using unsupervised deep learning applied to four-dimensional cardiac imaging data in 78,113 adult participants, that latent representations of left-ventricular motion reveal 39 genetic loci, two-thirds undetected by conventional indices, exposing a diversity of cell-type specificity validated across single-cell transcriptional atlases of the human heart. Conventional traits resolve to cardiomyocyte and conduction-system regulatory elements, while motion-specific variants converge on non-myocyte programmes encompassing autonomic neuronal, endocardial, fibroblast, and endothelial compartments. All of Us, FinnGen and GPMap data provided locus-level corroboration. Motion-specific traits further implicate environmental exposures including smoking and air pollution linked to motion profiles, and unsupervised clustering stratifies participants by genetic burden and clinical risk, uncovering disease-relevant endophenotypes invisible to standard clinical assessment.

## Main

Cardiac motion arises from a coordinated sequence of contraction, torsion, relaxation, and untwisting that depends on myocardial fibre architecture, electromechanical coupling, developmental patterning, and loading conditions^1^. Conventional phenotyping of these processes relies on simple global indices such as ejection fraction and volumetric measures derived from two-dimensional imaging. These scalar traits compress spatially and temporally heterogeneous phenotypes and may be insensitive to heritable variation in regional contractile coordination, fibre geometry, and active relaxation^2^. Consequently, efforts to map genetic and environmental determinants of cardiac motion onto the underlying biological programmes that shape ventricular structure and function have been limited.

Advances in computer vision now enable three-dimensional motion estimation from routine cardiac cine imaging^3,4^. Image-to-mesh frameworks reconstruct subject-specific biventricular geometries over time, transforming planar short-axis and long-axis views into digital atlases of whole heart motion that can be aggregated across large populations. Unsupervised deep learning can then compress these high-dimensional motion trajectories into compact latent representations that capture non-linear patterns of variation^5,6^. Motion models have already shown superior prognostic discrimination^7^ and improved disease classification compared to conventional indices^8^, suggesting that they serve as more biologically informative intermediate traits for genetic and environmental association studies.

Here, we combined four-dimensional (4D) analysis of cardiac magnetic resonance (CMR) imaging with unsupervised representation learning to map the genetic and environmental architecture of left-ventricular motion at population scale. Using an image-to-mesh pipeline we reconstructed fine details of motion dynamics of the left ventricle throughout the cardiac cycle for 78,113 UK Biobank participants, aligning each individual’s data to a common reference, and encoding it into latent features. Using these latent features as quantitative traits, we assessed how variants across the allele frequency spectrum contribute to complex patterns of heart motion, applying a hierarchical framework for cell-type-resolved regulatory validation of discovered loci. We also performed an exposome-wide analysis to identify how environmental factors influence cardiac motion. Finally, by distilling 4D cardiac motion into a continuum of diverse trajectories, we uncovered a taxonomy of motion patterns, yielding groups with differing genetic burden, exposure profiles, and risk of major adverse cardiovascular events.

Together, these analyses reveal that comprehensive analysis of human cardiac motion increases the discoverability of genetic, environmental, and biological architecture compared to conventional phenotyping, exposing non-myocyte regulatory programmes, environmentally imprinted motion signatures, and heritable prognostic traits.

## Results

### Study overview

We analysed cardiovascular magnetic resonance (CMR) imaging from 78,113 participants in the UK Biobank^9^. Conventional image-derived phenotypes of ventricular mass and volumes were obtained using deep learning based image segmentation^10^, with cohort characteristics summarised in Supplementary Table 1. To capture cardiac motion at high spatiotemporal resolution, we applied *DeepMesh*^4^, a computer vision framework that reconstructs time-resolved 3D mesh sequences of the left ventricle from cine imaging acquired in both short-and long-axis views. This approach integrates information across imaging planes to provide a unified representation of cardiac dynamics. Each participant’s motion data is then rigidly aligned to a common atlas space to enforce spatial consistency and enable population-level comparison. These mesh sequences were used directly to characterise the spatiotemporal signaturess of cardiomyopathy-associated variants and environmental exposures across the population. We then derived low-dimensional representations of cardiac motion using *Cardio4D-VAE*, a variational autoencoder that captures both spatial deformation and temporal dynamics across the cardiac cycle. These latent features were used as quantitative traits for genome-wide association analyses across the allele frequency spectrum, and to assess associations with environmental exposures. In addition, unsupervised graph-based clustering of motion patterns was used to derive a phenotypic taxonomy of cardiac motion with associated prognostic and genetic risk profiles. An outline of the study design is shown in Figure 1. In this study, *Cardio4D-VAE*-derived motion phenotypes were compared with conventional CMR phenotypes.

**Figure 1.**
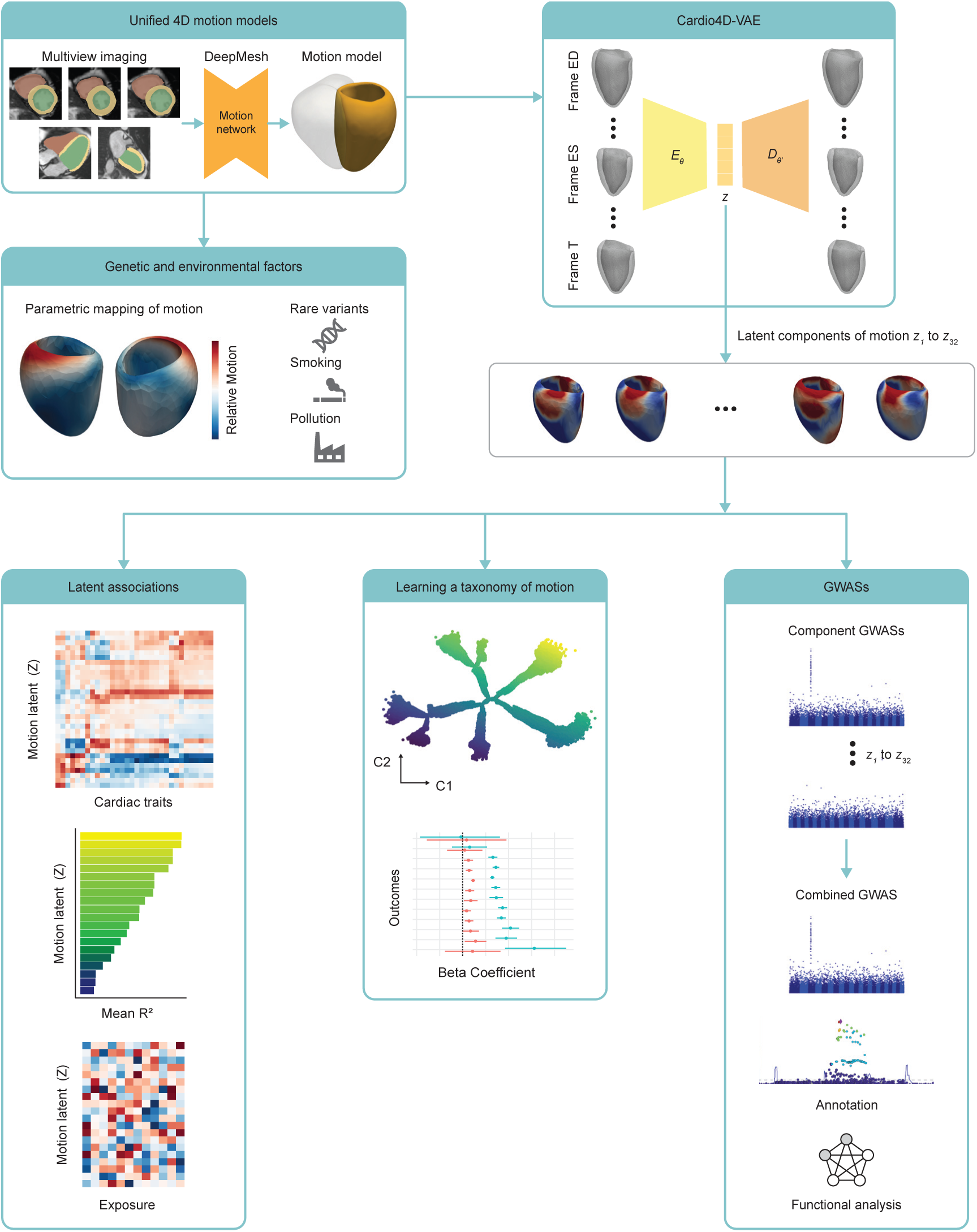
Study flowchart. A single model of three dimensional heart motion was created from multiple long and short axis views using cardiac magnetic resonance imaging. Each was aligned to a common reference space for analysis. The relationship between left ventricular motion and both rare variants and selected environmental exposures was modelled. Variation in heart motion across 78, 113 participants was reduced to 32 latent components using the Cardio4D variational autoencoder. The relationship between the latent components and conventional volumetric parameters was assessed. Associations with environmental exposures was also mapped. A genome wide association study (schematic plots shown) looked at associated common variants across all latent features. An unsupervised ranked clustering was performed on the latent components to define a taxonomy of cardiac motion diversity mapping genetic and environmental features on the tree.

We also benchmarked the nonlinear *Cardio4D-VAE* approach against a linear PCA-based representation derived from the same cardiac motion data.

### Image analysis

We performed automated quality-controlled analysis of CMR cine imaging to assess bi-atrial and bi-ventricular volumes and function, as well as left ventricular mass. Diastolic function was assessed using deformation analysis to derive end-diastolic strain rates^11^. Left ventricular ejection fraction (LVEF), indexed left ventricular end-diastolic volume (LVEDVi) and global longitudinal strain (Ell global) were taken as benchmark comparators for complex motion traits. Central vascular function was assessed by measuring aortic distensibility from central blood pressure estimates and dynamic aortic imaging. In total, 32 quantitative imaging phenotypes characterising structure and function were generated for each participant. For 78,113 participants, *DeepMesh* created 4D spatiotemporal models of cardiac motion mesh, with 50 temporal frames aligned to a common atlas template. Each surface mesh generated by *DeepMesh* consisted of 22,043 vertices and was down-sampled to 1,200 for analysis, providing 4.7 billion spatial coordinates across the cohort. *DeepMesh* achieved a mean surface distance error of 1.7 mm, compared to 3.0 mm for models that only use single-plane imaging – with the greatest improvement in estimating long axis function^4^.

### Latent encoding of structure and function

The 4D cardiac motion data were reduced to 32 latent features (*z*_1_–*z*_32_) using *Cardio4D-VAE*, with the latent dimensionality optimised through hyperparameter tuning. A visualisation of the shape and motion characteristics of each latent component is shown in Supplementary Figure 1. For each latent component, this figure presents the corresponding GWAS results, population-level cardiac motion trajectories, and regional left ventricular shape and motion changes.

We first assessed the relationship between latent components and conventional imaging parameters (see Methods). Taking all components as multivariable predictors, adjusted for age at MRI, sex, ethnicity, and body surface area, we found the highest explained variance for ventricular volumes (*R*^2^ = 0.65 - 0.78) and dynamic traits (*R*^2^ = 0.36 - 0.51). Vascular traits were weakly correlated (*R*^2^ < 0.1) (Figure 2a).

**Figure 2.**
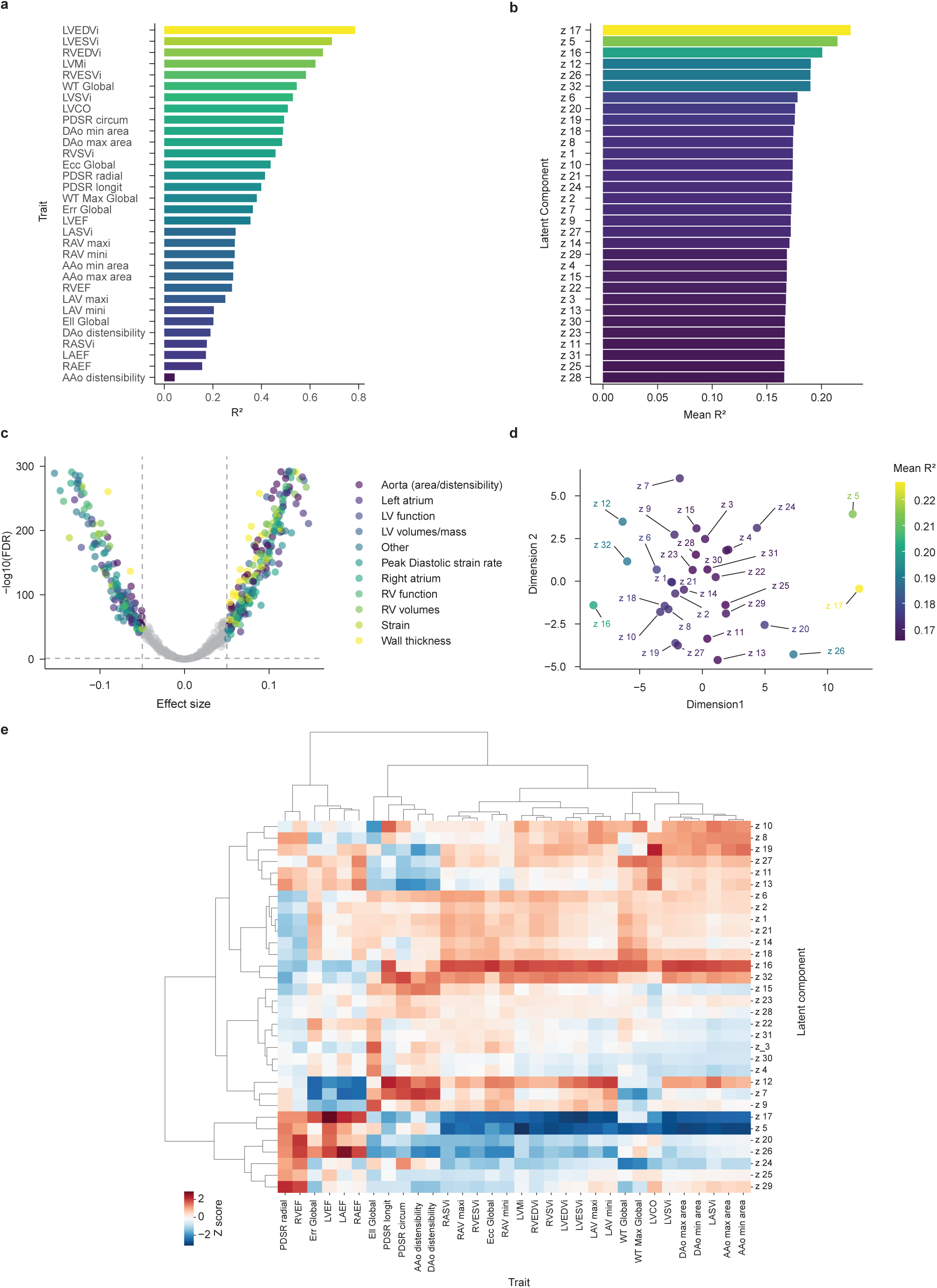
Predictive power and trait associations of motion-derived latent components. Panel **a** is based on the multivariable model including all 32 latent components, whereas panels **b–e** are based on analyses of individual latent components. **a**, Imaging trait predictability from motion-derived latent components, presented by variance explained (*R*^2^). **b**, Mean variance explained (*R*^2^) by each component across traits, each bar represents the mean variance explained by a single latent variable across all CMR traits. **c**, Volcano plot of component–trait associations colored by trait category, revealing numerous robust associations across cardiac and vascular traits. **d**, Two-dimensional projection of standardised effect size profiles across traits, showing the similarity structure of the 32 latent components, colored by mean *R*^2^. **e**, Hierarchically clustered heatmap of covariate-adjusted component–trait associations (Euclidean distance; Ward linkage); (statistical heatmap result is shown in Supplementary Figure 2.) AAo, ascending aorta; DAo, descending aorta; Ell, longitudinal strain; Ecc, circumferential strain; Err, radial strain; LAEF, left atrial ejection fraction; LASVi, left atrial stroke volume indexed; LAVmax, maximum left atrial volume indexed; LAVmin, minimum left atrial volume indexed; LV, left ventricle; LVCO, left ventricular cardiac output; LVCi, left ventricular cardiac index; LVEDVi, left ventricular end-diastolic volume indexed; LVEF, left ventricular ejection fraction; LVESVi, left ventricular end-systolic volume indexed; LVMi, left ventricular mass indexed; LVSVi, left ventricular stroke volume indexed; PDSR, peak diastolic strain rate; RAEF, right atrial ejection fraction; RASVi, right atrial stroke volume indexed; RAVmax, maximum right atrial volume indexed; RAVmin, minimum right atrial volume indexed; RV, right ventricle; RVEDVi, right ventricular end-diastolic volume indexed; RVEF, right ventricular ejection fraction; RVESVi, right ventricular end-systolic volume indexed; RVSVi, right ventricular stroke volume indexed; WT, myocardial wall thickness.

We then assessed the variance explained by individual latent components averaged across all imaging traits adjusted for the same covariates (see Methods). Components *z*_17_, *z*_5_, and *z*_16_, had the most explanatory power across multiple conventional traits, while others (e.g., *z*_28_ and *z*_25_) showed weaker associations (Figure 2b). The strength and significance of individual component–trait associations are summarised in Figure 2c, showing a large number of robust associations across diverse trait categories with FDR-corrected significance and clinically-meaningful effect sizes. A two-dimensional principal components analysis of standardised effect size profiles (Z scores) across traits showed that the latent components evenly captured underlying data variation and explainability with no linear structure or substantial grouping (Figure 2d). Hierarchical clustering of the component–trait association profiles further revealed groups of latent components with similar phenotypic association patterns (Figure 2e).

To benchmark the nonlinear *Cardio4D-VAE* representation against a conventional linear decomposition, we repeated the analysis using the first 32 principal components derived from the same motion data (Supplementary Figure 19). For PCA most of the explanatory power was captured by the first few principal components. In contrast, *Cardio4D-VAE* distributed information more evenly across 32 latent features and exhibited more representative association patterns with conventional traits. Together, these findings indicate that *Cardio4D-VAE* captures a broader spectrum of cardiac phenotypic variation while providing a less redundant representation than PCA.

### GWAS of latent motion traits identifies novel loci

We performed genome-wide association studies (GWAS) of 32 motion-derived latent traits (*z*_1_–*z*_32_) in 65,333 UK Biobank participants of European ancestry. Trait-wise discovery used a conventional genome-wide significance threshold of *P* < 5 ×10^−8^ and, for the combined analysis across latents we applied a Bonferroni-corrected threshold of *P* <5 ×10^−8^∕*m*_eff_, where *m*_eff_ = 7 from the eigen-spectrum of the latent–latent correlation matrix^12^. The combined latent analysis identified 39 independant genomic loci (pan-trait Manhattan plot, Figure 3).

**Figure 3.**
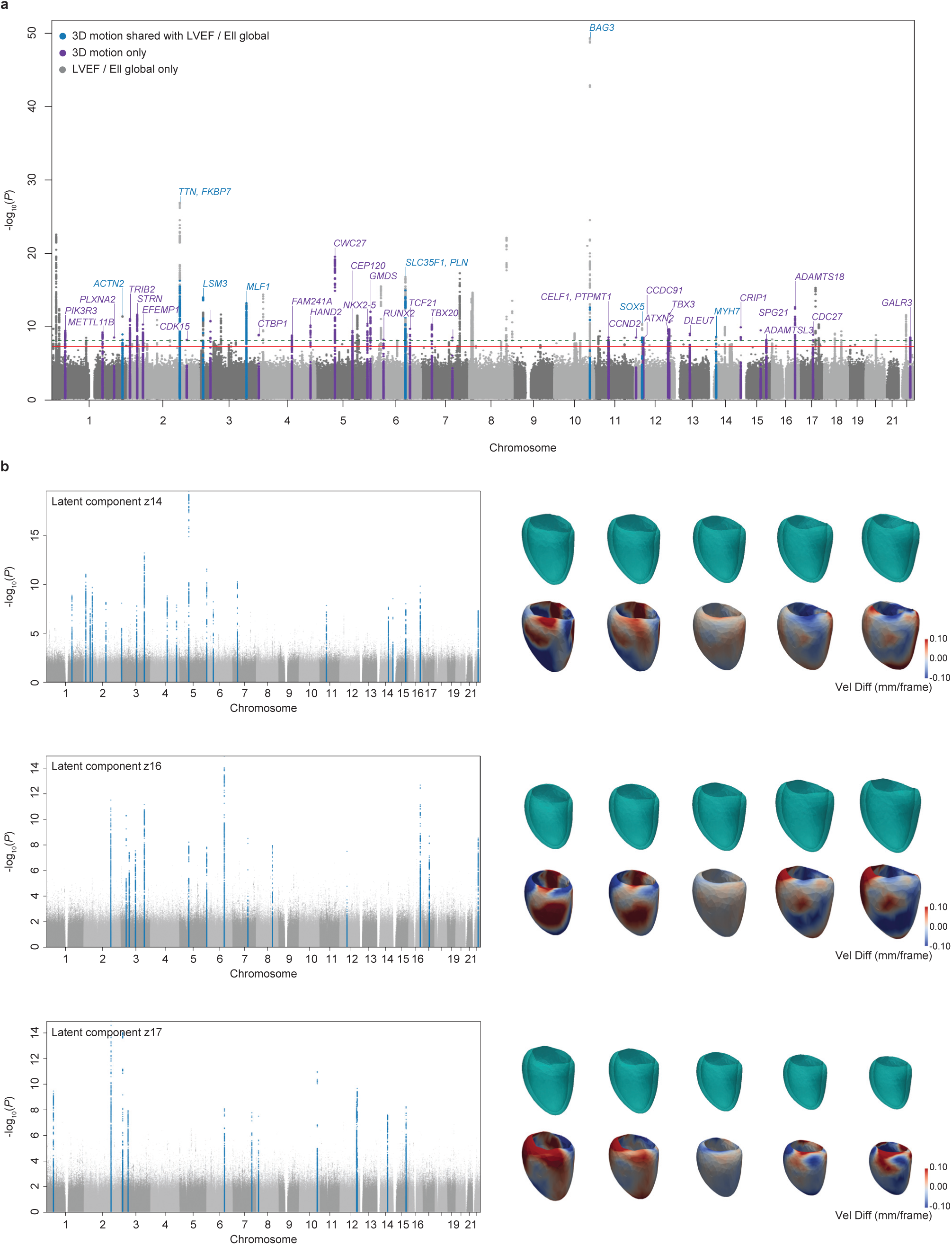
Genome wide association studies of left ventricular motion. **a**, Manhattan plot summarising GWAS of 32 latent components derived from four-dimensional left ventricular (LV) motion. Associations for two conventional traits, left ventricular ejection fraction (LVEF) and global longitudinal strain (Ell global), are shown for comparison. Conventional traits use the standard genome-wide significance threshold of *P* = 5×10^−8^ (− log_10_ *P* = 7.3), while the latent traits apply a multiple-testing–corrected threshold of *P* = 7.14 × 10^−9^ (− log_10_ *P* = 8.15), corresponding to *M*_eff_ = 7.14 effective independent motion components. Loci shared with LVEF or Ell global are indicated in blue, and latent-only loci are shown in purple. **b**, Example GWAS results for three representative latent components, each capturing a distinct spatiotemporal motion pattern. Right panels show reconstructed LV geometries at end-diastole (ED) averaged across quantiles of the latent variable (from left to right: [0.00–0.02], [0.09–0.11], [0.49–0.51], [0.89–0.91], and [0.98–1.00]). Colour maps indicate velocity differences (mm per frame) at ED between the quantile-averaged shape and the corresponding control (non-carrier) cohort, highlighting regional mechanical variation associated with genetic loading.

By comparison, conventional GWAS of global function metrics identified 43 and 13 loci for LVEF and Ell global, respectively, at genome-wide significance. Of the 39 latent loci, 31 (79%) were not detected by LVEF or Ell global (Table 1), indicating that they recover genetic signal largely orthogonal to conventional global indices. We additionally examined LVEDVi to test whether the latent traits captured features beyond chamber volume; it yielded 50 loci at *P* < 5 × 10^−8^, 11 of which overlapped a motion locus. Novelty was assessed against the cumulative union of LVEF, Ell global and LVEDVi: 26 of 39 motion loci (67%) passed the stricter motion-domain threshold (*P* < 7.14×10^−9^; *m*_eff_ = 7) but were not genome-wide significant in any comparator scan (*P* < 5×10^−8^), supporting complementary genetic discovery beyond these prespecified scalar traits (Table 1; Supplementary Data S1).

**Table 1.**
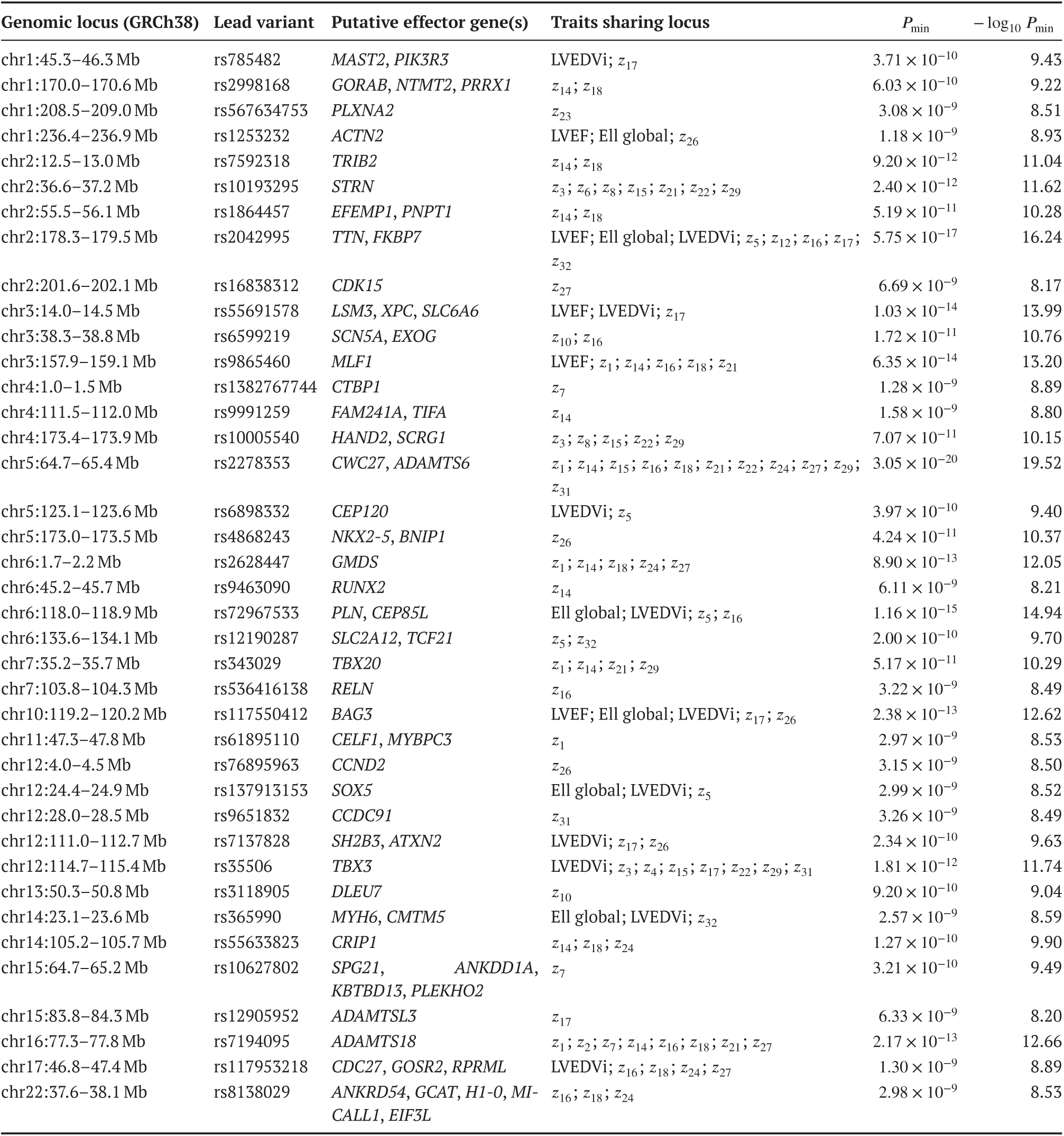
Lead loci from GWAS of LV motion latent traits and shared conventional traits. Each row represents an LD-clumped genomic locus (GRCh38; coordinates in megabases). Variants with the lowest *P*-value across all latent traits within each locus are listed. The *Traits sharing locus* column lists all traits in which a lead SNP lies within the same clumped locus—these may include the sentinel variant itself or distinct trait-specific lead SNPs in high linkage disequilibrium. For each locus, the minimum association *P*-value across traits (*P*_min_) and its − log_10_ transform are reported. All loci surpass the study-wide significance threshold (*P* < 7.14 × 10^−9^) after clumping at *R*^2^<0.1 within ±250 kb. Full base-pair coordinates are provided in Supplementary Data S1.

### Latent motion traits are heritable and localise to active regulatory regions

All traits were heritable (ℎ^2^ ranging from 0.05 to 0.25), establishing a genetic basis for discovery (Supplementary Data S2). Bivariate genetic (*r*_*g*_) and phenotypic (*r*_*p*_) correlations revealed a large contiguous block of positive correlations concordant between levels, indicating shared polygenic architecture anchored within the established biological landscape (Supplementary Figure 3a). Known relationships such as the negative correlation between LVEF and LVEDVi (*r*_*g*_ = −0.41, *r*_*p*_ = −0.23) were recapitulated with correct directionality. LDSC intercepts close to 1.00 confirmed minimal confounding from population structure (Supplementary Figure 3b). S-LDSC (baselineLD v2.2, GRCh38) partitioned heritability across functional annotations, with the strongest positive conditional effects (*τ*^∗^; FDR < 0.05) implicating active *cis*-regulatory DNA and evolutionary constraint. Heritability was depleted at architectural and flanking regions, a pattern recapitulated for conventional traits (Supplementary Figure 4; Supplementary Data S3).

### Functional gene mapping and pathway enrichment

The FUMA framework^13^ prioritised 136 genes for the latent traits through positional proximity, eQTL mapping, and Hi-C-derived chromatin contacts. Combined with manual inspection with LocusZoom^14^, this yielded 157 putative locus-to-gene assignments for downstream analyses (Supplementary Data S4; Supplementary Figure 5). Pathway analysis demonstrated significant enrichment for cardiac muscle patterning and development (Figure 4a). After excluding genes shared with conventional traits, enrichments for chamber and ventricular development persisted (Figure 4b). The motion-only catalogue included regulators of chamber specification and epicardial–fibroblast fate (*TBX20*, *TBX3*, *NKX2-5*, *HAND2*, *FOXC1*, *TCF21*), extracellular-matrix and mechanotransductive genes (*ADAMTS6*, *EFEMP1*), and *SCN5A*. The shared set was dominated by cardiomyocyte-core sarcomeric processes (Figure 4b). MAGMA gene-based analysis^15^ confirmed heart as the principal tissue of action, with greater cardiac selectivity for motion than conventional traits (Figure 4c).

**Figure 4.**
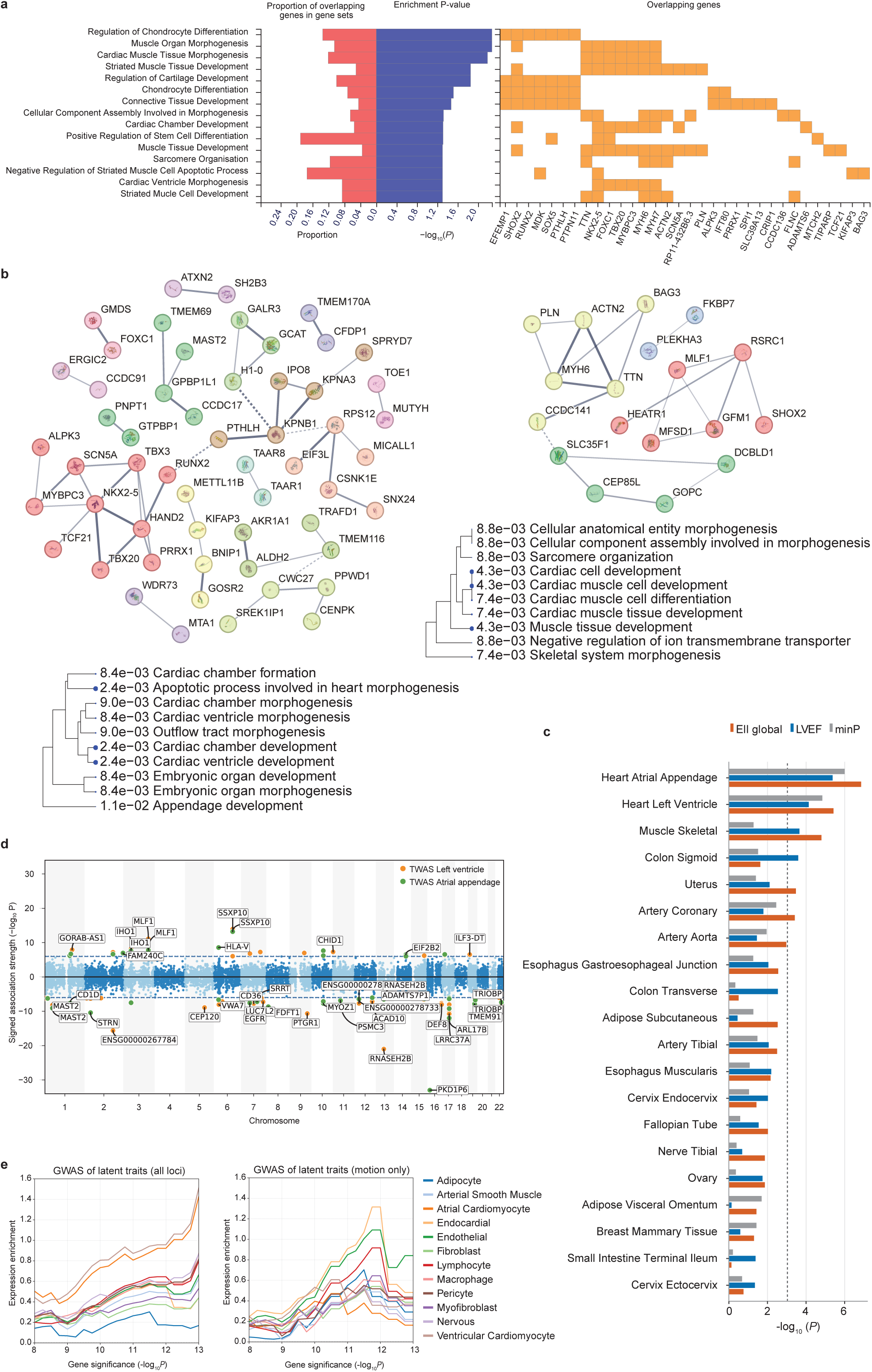
Multi-layer functional and cellular mapping of genetic determinants of LV motion. **a,** Gene Ontology (GO:BP) enrichment for effector genes associated with latent motion loci. Terms converge on muscle and cardiac morphogenesis, chamber development, and sarcomere organisation. **b,** STRING protein–protein interaction networks of latent-only (left) and shared (right) gene sets. Markov clustering (inflation parameter = 2) reveals modules corresponding to cardiac morphogenesis and developmental signalling (latent-only), and sarcomere and excitation–contraction pathways (shared). Disconnected nodes in the network not shown. Corresponding hierarchical clustering trees summarise pathway correlations; blue dots indicate significant *P*-values. **c,** MAGMA tissue enrichment across 54 GTEx tissues for latent min-*P* (orange), LVEF (blue), and Ell global (magenta). Only the top 20 are shown. The Bonferroni-corrected threshold (*P* < 0.05∕54, − log_10_ *P* ≈ 3.03) is indicated by the dashed line. **d,** TWAS of latent traits in LV (orange) and AA (green) using FUSION/GTEx v8 models. The Miami plot shows signed – log_10_ *P* for genes with valid expression models; annotated genes represent the top 20 significant loci. **e,** Sliding-threshold single-nucleus RNA-seq enrichment. Curves represent the standardised mean difference in log-transformed expression between the motion GWAS gene set and background across increasing − log_10_ *P* thresholds. The full latent gene set is enriched in cardiomyocytes (VCM, ACM), whereas latent-only genes show attenuated cardiomyocyte enrichment, uncovering a signal in endocardial, endothelial, and lymphocyte lineages, suggesting complementary non-myocyte programmes.

### TWAS and single-cell integration corroborate a non-myocyte axis

Transcriptome-wide association studies in Heart LV and Atrial Appendage (GTEx v8; FUSION^16^) independently recovered genes concordant with locus-to-gene mapping, including *ALPK3*, *MAST2*, *TCF21* and *TRIOBP* (Figure 4d; Supplementary Data S5), together with *CD36*—wherein a variant was recently identified as a prevalent cause of dilated cardiomyopathy in African-ancestry populations^17^—and *EGFR*, whose signalling drives cardiac fibrosis and hypertrophic remodelling. Sliding-threshold integration with an adult cardiac single-nucleus atlas showed that motion-only genes additionally localised to endocardial, endothelial and lymphocyte populations beyond the cardiomyocyte signal shared with conventional traits (Figure 4e; Supplementary Figure 6). FUMA, MAGMA and TWAS therefore provided first-pass locus, pathway and transcriptomic context; final candidate-gene prioritisation was derived from the fine-mapped hierarchy described below.

### Fine-mapped variants implicate non-myocyte regulators of cardiac motion

Across all 35 traits, SuSiE-RSS fine-mapping retained 87 loci, comprising 19 motion-only, 11 shared and 57 conventional-only loci. Seven motion latents showed at least one significant cell-state enrichment in FDR-corrected PIP-weighted scE2G analyses (Figure 5a; Supplementary Data S6; Supplementary Figure 7). Representative signals included *z*_17_ in parasympathetic neurons (*q* = 1.06 × 10^−8^) and valvular interstitial cells (VIC_5, *q* = 4.66 × 10^−7^); *z*_12_ in valvular interstitial cells (VIC_1, *q* = 4.91 × 10^−7^) and atrial endocardial cells (*q* = 1.78 × 10^−5^); and *z*_27_ in atrial fibroblasts (*q* = 1.24 × 10^−5^). Conventional traits showed the reciprocal cardiomyocyte and conduction profile, led by LVEF enrichment in ventricular and atrial cardiomyocytes (VCM, *q* = 1.16 × 10^−21^; ACM, *q* = 1.86 × 10^−21^).

**Figure 5.**
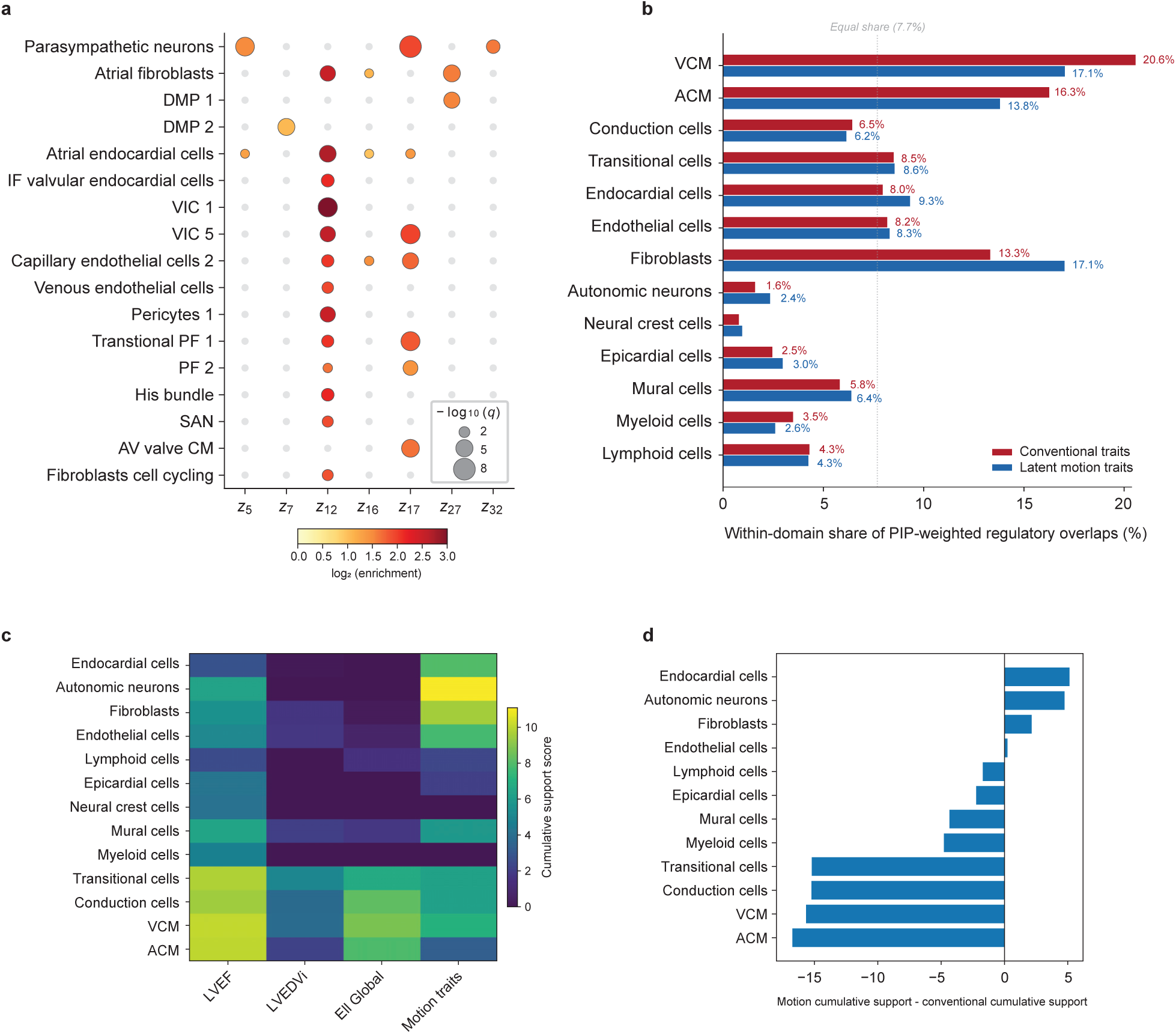
Cell-type-specific regulatory programmes captured by motion-derived versus conventional imaging traits. **a,** Dot plot of cell-state enrichment for the seven motion latents with at least one significant association (*q* < 0.05). Bubble size encodes significance (− log_10_(*q*)); colour encodes effect size (log_2_ enrichment). Each latent resolves to a distinct non-myocyte regulatory axis: *z*_5_, *z*_17_, and *z*_32_ map to parasympathetic neurons; *z*_27_ to atrial fibroblasts and dorsal mesenchymal protrusion (DMP) states; *z*_12_ broadly to endocardial, valvular interstitial, and fibroblast programmes; *z*_7_ to DMP states; and *z*_16_ to endocardial and endothelial cells. Grey dots indicate non-significant enrichments. **b,** Normalised regulatory support within each trait domain, scaled to sum to 100% to enable comparison independent of total signal magnitude. The conventional distribution is the mean of LVEF, LVEDVi, and Ell global; the motion distribution is cumulative across all 32 latent traits. The combined cardiomyocyte share (VCM + ACM) decreases from 36.9% in conventional traits to 30.9% in motion, with the difference redistributed to fibroblasts, endocardial cells, autonomic neurons, mural cells, and epicardial cells. The VCM+ACM share is a deterministic normalisation of cumulative PIP-weighted regulatory support and was not used for statistical inference. **c,** Heatmap of cumulative support scores across cell-type groups for the three conventional traits and the motion cumulative score (sum of positive meta-Z contributions across all 32 latent traits). Conventional traits concentrate support in cardiomyocyte, conduction, and transitional cell programmes, whereas the cumulative motion signal additionally encompasses autonomic neurons, endocardial cells, fibroblasts, and endothelial cells. **d,** Motion-specificity scores (Δ = motion cumulative support − conventional cumulative support) for each cell-type group. Positive values indicate programmes preferentially captured by motion latents; negative values indicate programmes dominated by conventional traits. Panels b–d are descriptive cumulative-support summaries; formal cell-state inference is provided by the FDR-corrected 90-state and 13-group scE2G enrichment tests reported in Supplementary Data S6.

The distinction was therefore not simply that different phenotypes implicated different cell types: conventional scalar traits localised mainly to cardiomyocyte and conduction programmes, whereas motion traits retained this core but additionally implicated cell states that organise the architecture, transmission and timing of ventricular deformation. For descriptive domain-level comparison, cumulative-support panels showed a reduction in cardiomyocyte share from 36.9% for conventional traits to 30.9% for motion (Figure 5b–d).

The tiered variant-to-gene framework resolved 40,973 credible-set variants (21.7%) to 643 priority genes (Figure 6a,b; Supplementary Figure 8a). High-confidence assignments spanning the evidence hierarchy included rs76895963 at *CCND2*, the *CRIP1* missense variant rs55633823 (p.A58V), and rs35506 at *TBX3* (Table 2). Motion-only loci carried a larger Tier 5 unannotated fraction than conventional-only loci (42.4% versus 33.9%), potentially representing a residual regulatory-annotation gap for dynamic cardiac traits (Figure 6a; Supplementary Figure 8b).

**Figure 6.**
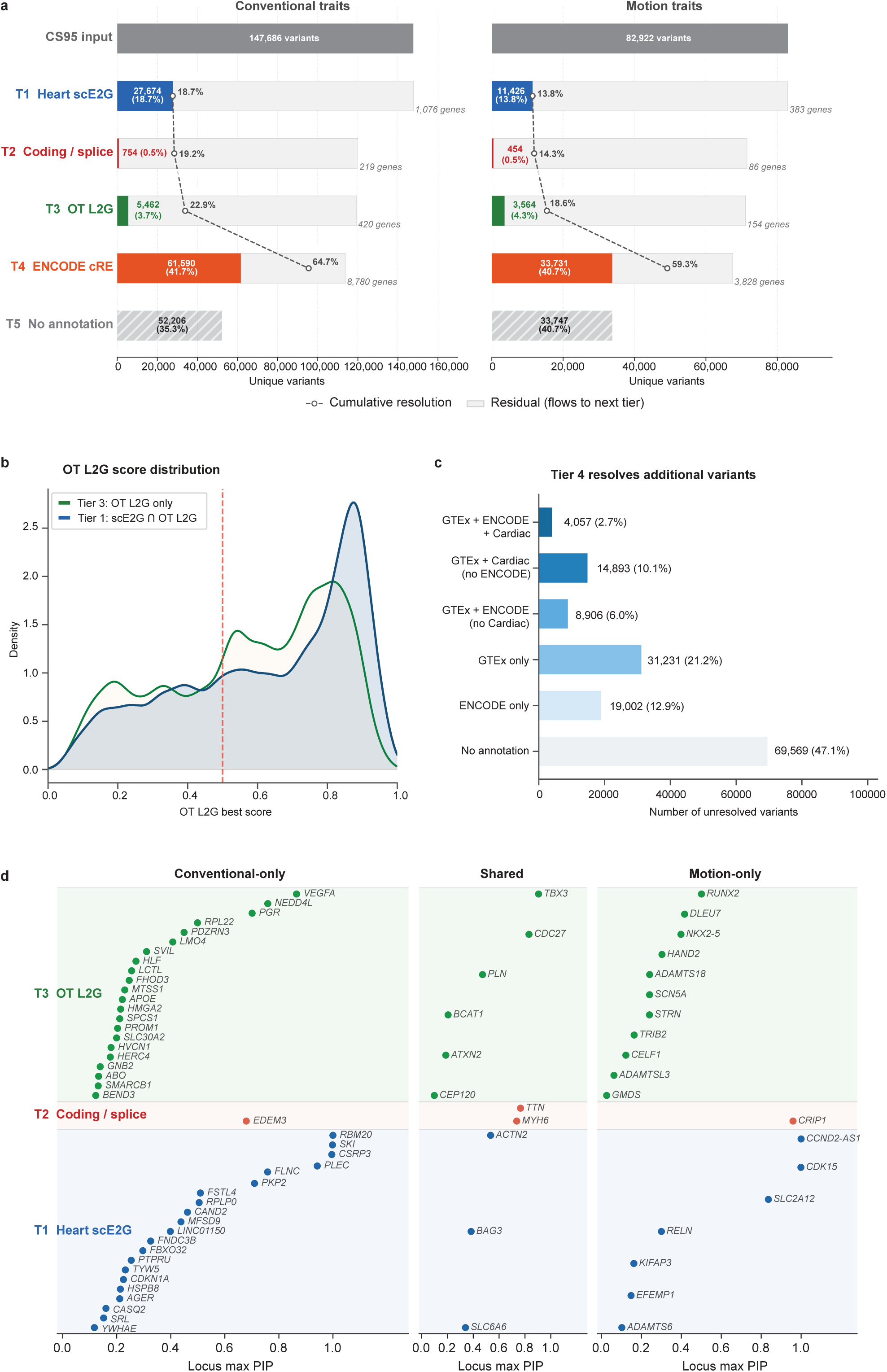
Hierarchical functional annotation resolves the majority of fine-mapped variants and reveals trait-domain differences in regulatory architecture. **a**, Stepwise variant annotation waterfall for conventional traits (left; 147,686 CS95 variants) and motion traits (right; 82,922 variants). Variants were sequentially assigned to the highest-confidence tier: Tier 1, overlap with foetal heart single-cell enhancer-to-gene (scE2G) links; Tier 2, coding or splice-region consequence (VEP HIGH/MODERATE); Tier 3, Open Targets locus-to-gene (L2G) score support; Tier 4, pan-tissue regulatory support (GTEx eQTL and/or ENCODE cCRE overlap); Tier 5, no annotation. Coloured bars show the number (and percentage) of unique variants resolved at each tier; dashed lines trace cumulative resolution. Gene counts denote the number of unique genes implicated per tier. **b**, Density distribution of Open Targets L2G scores for variants resolved by scE2G alone (Tier 1; blue, n = 32,835) versus OT L2G (Tier 3; green, n = 7,271). The confidence in variant scoring is reduced after being solved by the heart regulome. **c**, Breakdown of Tier 4 variants (n = 78,089) by annotation source: GTEx eQTL alone, ENCODE cCRE alone, GTEx eQTL plus cardiac eQTL, GTEx eQTL plus ENCODE cCRE, and GTEx eQTL plus ENCODE cCRE plus cardiac eQTL. **c**, Prioritised genes plotted by locus maximum posterior inclusion probability (PIP), stratified by trait domain: conventional-only (left), shared (centre), and motion-only (right). Genes are coloured by their highest-confidence evidence tier (blue, Tier 1 scE2G; red, Tier 2 coding/splice; green, Tier 3 OT L2G). Each point represents the top-ranked gene at a distinct fine-mapped locus (84 loci total); one gene per locus is shown, selected as the gene carrying the highest individual variant PIP at that locus.

**Table 2.** Fine-mapped variants and prioritised genes across evidence tiers. Selected illustrative examples of high-confidence loci variants ranked by PIP within each trait domain. Tier 1 (T1), foetal heart scE2G regulatory overlap; Tier 2 (T2), protein-altering or splice consequence; Tier 3 (T3), Open Targets Locus-to-Gene ≥ 0.5; Tier 4 (T4), pan-tissue GTEx/ENCODE; Tier 5 (T5), unannotated. Ell, global longitudinal strain; DCM, dilated cardiomyopathy; HCM, hypertrophic cardiomyopathy; ARVC, arrhythmogenic right-ventricular cardiomyopathy; ASD, atrial septal defect; CM, cardiomyocyte; TF, transcription factor; MHC, myosin heavy chain.

| Variant | Trait(s) | PIP | Tier | Gene | Biological context |
| --- | --- | --- | --- | --- | --- |
| **Conventional-only loci** |  |  |  |  |  |
| rs189569984 | LVEF, LVEDVi | >0.999 | T1 | *RBM20* | Missense p.S455L; cardiac RNA splicing regulator; DCM gene |
| rs2503715 | LVEF | 0.998 | T1 | *SKI* | TGF-β repressor; inhibits fibroblast activation and fibrosis |
| rs45550635 | LVEF, Ell | 0.996 | T1 | *CSRP3* | Missense p.W4R; Z-disc mechanosensor; HCM/DCM gene |
| rs11786896 | LVEF | 0.942 | T1 | *PLEC* | Cytoskeletal linker; desmosome–sarcolemma scaffold |
| rs34373805 | LVEF, LVEDVi | 0.758 | T1 | *FLNC* | Actin crosslinker at Z-disc; arrhythmogenic cardiomyopathy gene |
| rs2045172 | LVEF | 0.710 | T1 | *PKP2* | Desmosomal plakophilin; ARVC gene |
| rs78444298 | LVEDVi | 0.679 | T2 | *EDEM3* | Missense; ER mannosidase; protein quality control |
| rs2146324 | LVEDVi | 0.865 | T3 | *VEGFA* | Angiogenesis regulator; coronary vascular remodelling |
| rs11660748 | LVEF | 0.759 | T3 | *NEDD4L* | E3 ubiquitin ligase; targets ENaC and Nav1.5 ion channels |
| rs73044299 | LVEDVi | 0.990 | T4 | *DMWD* | Myotonic dystrophy locus (*DMPK/SIX5*); conduction disease and cardiomyopathy; atrial eQTL |
| rs79675252 | LVEDVi | 0.903 | T5 | — | No annotation; nearest gene *ANKRD10* |
| rs4439609 | Ell | 0.892 | T5 | *MTUS2* | Microtubule scaffold; locus carries lower-PIP T1/T4 variants |
| **Shared loci** |  |  |  |  |  |
| rs35506 | LVEDVi, $z_{3,4,15,22,29,31}$ | 0.905 | T3 | *TBX3* | Conduction-system T-box TF; ulnar–mammary syndrome |
| rs117953218 | LVEDVi, $z_{16,18,24,27}$ | 0.831 | T3 | *CDC27* | APC/C subunit; cardiomyocyte cell-cycle exit regulation |
| rs2042995 | LVEF, LVEDVi, Ell | 0.766 | T2 | *TTN* | Missense p.I10522V; sarcomeric spring; leading DCM gene |
| rs365990 | LVEDVi, $z_{32}$ | 0.736 | T2 | *MYH6* | Missense p.V1101A; α-MHC; ASD and sick sinus syndrome |
| rs4659701 | LVEF, Ell, $z_{26}$ | 0.534 | T1 | *ACTN2* | Z-disc α-actinin; anchors titin and actin; DCM/HCM gene |
| rs72967533 | LVEDVi, Ell, $z_{5,16}$ | 0.472 | T3 | *PLN* | Phospholamban; SERCA2a inhibitor; DCM and HCM gene |
| **Motion-only loci** |  |  |  |  |  |
| rs76895963 | $z_{26}$ | >0.999 | T1 | *CCND2/CCND2-AS1* | Atrial-CM promoter; GATA4 co-factor; drives CM proliferation |
| rs16838312 | $z_{27}$ | >0.999 | T1 | *CDK15* | Atrial-fibroblast context; cyclin-dependent kinase |
| rs55633823 | $z_{14,18,24}$ | 0.959 | T2 | *CRIP1* | Missense p.A58V; LIM-domain protein; TGF-β signalling in fibroblasts |
| rs12190287 | $z_5, z_{32}$ | 0.836 | T1 | *TCF21/SLC2A12* | Parasympathetic-neuron context; epicardial TF and glucose transporter |

### Prioritised genes are transcriptionally active in adult myocardium

The foetal scE2G atlas provided variant-level regulatory localisation, particularly for developmental and morphogenetic loci. As an independent adult expression-validation layer, we analysed single-nucleus RNA-seq from 612,457 left ventricular nuclei across 70 donors^18^ (Supplementary Figure 9a–c).

Motion genes showed preferential detection in fibroblasts, endocardial cells, and autonomic neurons alongside cardiomyocytes—a pattern absent for conventional-trait genes and derived independently of the GWAS and scE2G analyses (Figure 9d).

In DCM, shared genes were broadly suppressed across cell types, whereas motion-only genes remained stable. Within the Motion set, the developmental/morphogenesis subset showed no coordinated DCM dysregulation (*p*_adj_ = 0.23; *p*_adj_ = 0.88 after excluding *NKX2-5*), whereas the adult cardiomyocyte-intrinsic subset was significantly downregulated (*p*_adj_ = 0.048; Supplementary Figure 9f,g). Thus, regulatory mapping and expression analysis independently converged on a multicellular motion programme extending beyond cardiomyocytes.

### External molecular and disease support of fine-mapped loci

External analyses connected the fine-mapped LV imaging loci to coherent molecular and cardiovascular disease biology across three complementary resources (Figure 7a). In the full GPMap fine-mapped locus universe, 17 of 24 motion-only loci contained developmental or congenital genes compared with 9 of 72 conventional-only loci (OR=17.0, FDR=6.1× 10^−7^; Supplementary Data S7). Among scE2G-supported credible sets, motion traits assigned more posterior probability to non-myocyte programmes than conventional traits (62.7% versus 53.6%; difference 9.1 percentage points, 95% CI 4.6–13.5; FDR=2.2 × 10^−5^; Figure 7b). In FinnGen R13, PIP-weighted aggregation across 13 cardiac endpoints supported 40 of 87 loci at FDR< 0.05 on the combined developmental, valvular and conduction axis (Figure 7c,d). Shared loci showed the broadest disease support, whereas motion-only loci were less dominated by myocardial-remodelling associations than conventional-only loci (5/19 versus 34/57; OR=0.24, *P* = 0.0168) while retaining conduction and valvular/endocardial support (Figure 7c). In All of Us, 18 loci were FDR-supported and 27 had nominal or FDR support across 49 EUR phenotypes; cross-ancestry META retained 17 and 29 loci, respectively. AFR, AMR and EAS provided lower-powered ancestry-specific sensitivity, whereas SAS lacked eligible axis endpoints (Figure 7e; Supplementary Data S7). Together, fine-mapping, hierarchical variant-to-gene assignment, adult LV single-nucleus analysis, and external molecular and disease corroboration constitute a causal-regulatory discovery framework that links motion-associated loci to candidate causal variants, effector genes and regulatory cell-state mechanisms (Figure 7a,f).

**Figure 7.**
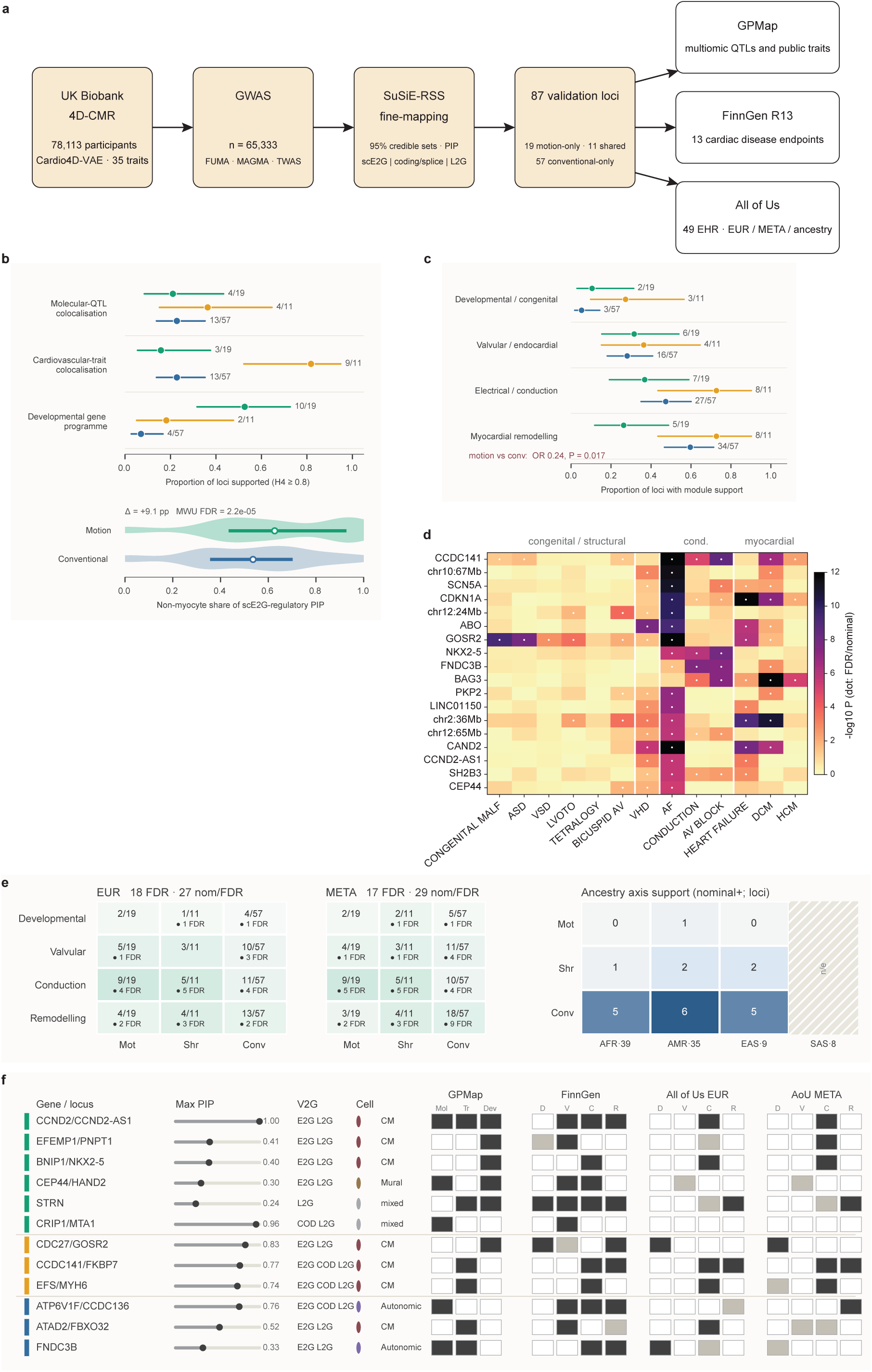
External molecular and disease-context validation of fine-mapped LV motion loci. The same 87-locus validation set (19 motion-only, 11 shared, 57 conventional-only; best-variant PIP ≥ 0.20) is evaluated through three orthogonal external read-outs. Locus classes are coloured motion-only (teal), shared (amber) and conventional-only (blue); in **b** and **c** points show proportions with 95% confidence intervals. **a,** Analytical workflow linking UK Biobank 4D cardiovascular traits to genome-wide association analysis (*n* = 65,333; motion threshold *P* < 7.14 × 10^−9^), conventional annotation with FUMA, MAGMA and TWAS, SuSiE-RSS fine-mapping and hierarchical variant-to-gene prioritisation, yielding the 87-locus set evaluated in GPMap, FinnGen R13 and All of Us. **b,** GPMap support within the common 87-locus set. Upper, proportion of loci colocalising with molecular QTLs or cardiovascular traits at *H*_4_ ≥ 0.8, or carrying a gene assigned to the developmental/congenital programme. Lower, credible-set-level distribution of non-myocyte scE2G-regulatory posterior inclusion probability (PIP); motion credible sets assigned a greater share of regulatory PIP to non-myocyte programmes than conventional credible sets (Δ = 9.1 percentage points; Mann–Whitney *U* test, FDR-adjusted *P* = 2.2 × 10^−5^). **c,** FinnGen disease-module support across four cardiac modules (13 prespecified endpoints). Shared loci showed the broadest disease support, whereas motion-only loci were less frequently associated with myocardial remodelling than conventional-only loci (5/19 versus 34/57; OR= 0.24, *P* = 0.0168). **d,** FinnGen locus–endpoint association landscape for leading externally supported loci. Columns represent the 13 cardiac endpoints grouped into developmental/congenital structural, valvular, electrical/conduction and myocardial-remodelling domains; colour denotes the − log_10_ *P* from PIP-weighted locus-level aggregation and overlaid dots distinguish nominal and FDR-supported associations. **e,** All of Us disease-module validation across 49 curated electronic health-record (EHR) phenotypes. The EUR ancestry-matched primary analysis and cross-ancestry META analysis report the number of loci with nominal-or-FDR support per class, with FDR-supported counts marked (EUR, 18 FDR and 27 nominal-or-FDR; META, 17 and 29). The right matrix shows combined developmental/valvular/conduction-axis support in AFR, AMR and EAS sensitivity analyses (headings give the number of available phenotypes). SAS lacked an eligible primary-axis endpoint and therefore not evaluated. **f,** Integrated evidence matrix for representative high-priority loci spanning all three classes, selected by a prespecified rule (ranked within class by number of supporting external resources, then maximum PIP). Maximum PIP is shown as a lollipop; variant-to-gene evidence indicates cardiac enhancer-to-gene mapping (E2G), coding or splice consequence (COD) and Open Targets locus-to-gene support (L2G), with the predominant cardiac cell programme. GPMap columns give molecular-QTL, cardiovascular-trait and developmental-programme support; FinnGen and All of Us columns give developmental/congenital (D), valvular/endocardial (V), electrical/conduction (C) and myocardial-remodelling (R) support. Black, grey and white squares denote FDR, nominal and no support; coloured sidebars identify locus class.

### Rare variants affect complex motion traits

Outside of familial disease, rare variants in cardiomyopathy-associated genes have an attenuated phenotype that manifests as changes in static geometry^19,20^. Using 4D motion models, we examined the consequence of harbouring these variants on complex motion traits. Here, we considered participants with predicted titin truncating variants (TTNtv, N = 252), associated with a predisposition to dilated cardiomyopathy (DCM), and pathogenic or likely pathogenic variants (P/LP) in sarcomeric genes associated with hypertrophic cardiomyopathy (HCM-PLP, N = 128). Each variant group was compared to non-carriers (N = 77,656) (see Supplementary Table 2 for genotype frequencies).

Figure 8a,b illustrates the absolute local distance maps and velocity differences between variant carriers and non-carriers across the cardiac cycle, while Figure 8c,d summarise global dynamic motion differences (see Methods). For TTNtv carriers, systolic contraction is delayed in a non-uniform pattern, with the septal wall most affected compared to controls, as well as an abnormal pattern of diastolic relaxation. In HCM-PLP carriers, diastolic relaxation is also affected, but there are more pronounced changes in systolic motion dynamics, especially in the basal left ventricle. Dynamic models of regional displacement and velocity differences (Supplementary Movies S1-S4) show that both TTNtvs and HCM-PLPs are associated with spatiotemporal changes in motion and ventricular coordination. This demonstrates that rare variants affect not only influence geometric remodelling but also complex temporal dynamics in the heart.

**Figure 8.**
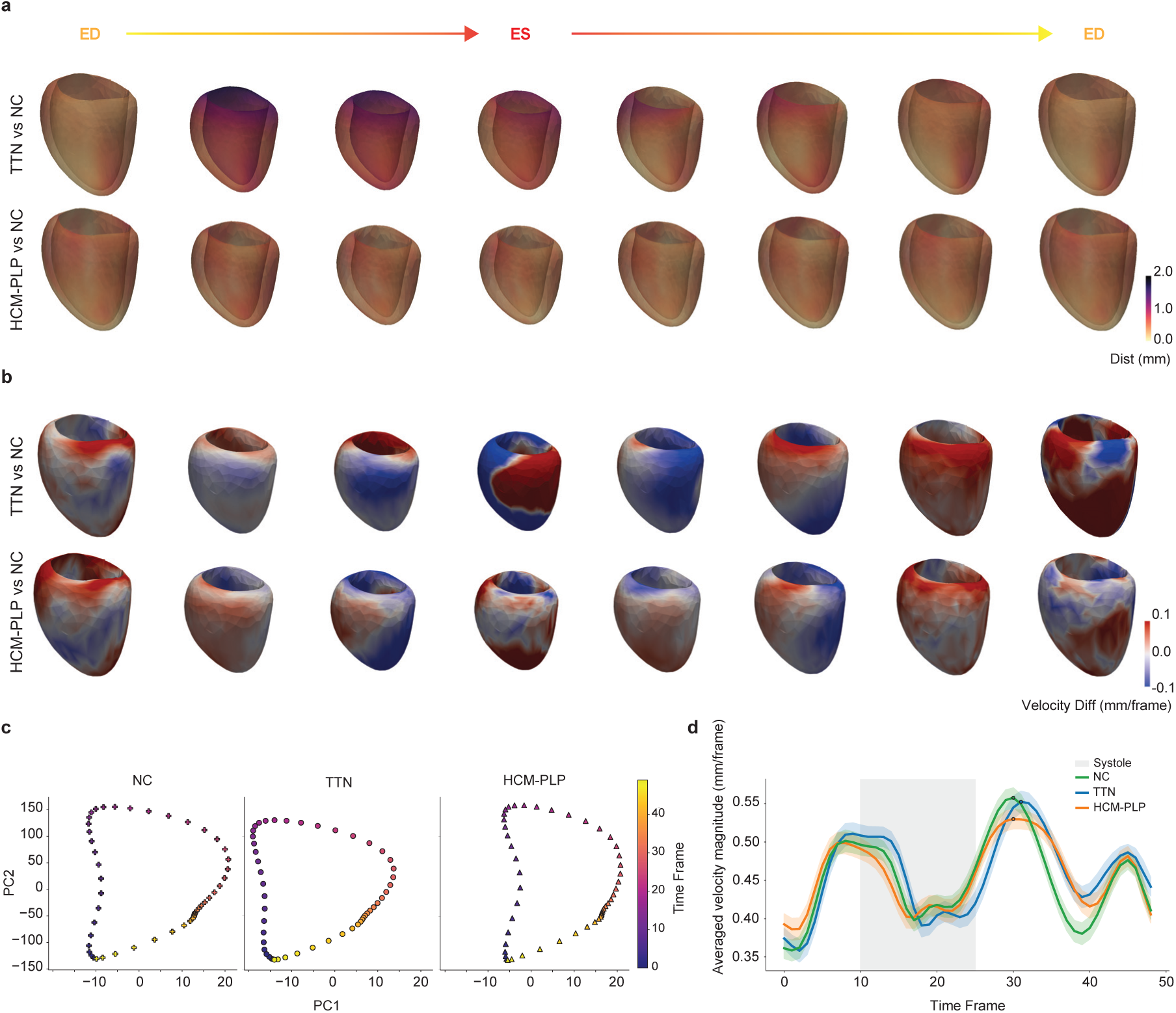
Spatiotemporal differences in left ventricular deformation and motion across genotype groups. Groups include carriers of titin truncating variants (TTN) and carriers of pathogenic or likely pathogenic variants (P/LP) in sarcomeric genes implicated in hypertrophic cardiomyopathy (HCM-PLP). **a**, Local displacement magnitude maps (Dist in mm) comparing variant carriers with non-carriers (NC) across the cardiac cycle. Warmer colours indicate regions where the average LV surface of carriers is more displaced relative to non-carriers at a given phase. **b**, Corresponding maps of local myocardial velocity differences (Velocity Diff in mm/frame). Red regions mark areas where carriers move faster than non-carriers, while blue regions highlight slower motion in carriers compared to non-carriers. **c**, Principal component analysis of average 4D motion. Each group follows a characteristic trajectory in PCA space, with colors indicating progression through the cardiac cycle. **d**, Averaged velocity magnitude curves (±95% CI) for different groups across the cardiac cycle. Gray shading marks systole (frames 10–25); peaks are indicated. This visualisation highlights group-level differences in motion intensity and temporal patterns rather than spatial variation across the genetic groups.

### Environmental exposures linked to ventricular dynamics

Environmental exposures modify cardiovascular disease risk and are associated with alterations in cardiac structure and function^21–23^. We used the 4D motion models to survey the associations of a wide range of exposures on complex motion traits by considering 25 environmental factors known to be linked with cardiovascular risk^21–23^. Figure 9 shows a hierarchically clustered heatmap of covariate-adjusted standardised effect sizes (*Z* scores) for significant associations between exposures and latent features. Alcohol and coffee intake were positively correlated with components *z*_16_, *z*_12_ and *z*_19_. Mobile phone use, which is a proxy for cardiometabolic risk and mental health^24^, also had a strong association with *z*_16_ but a different pattern among other latents. In contrast, an opposite association for these latents was observed for a range of dietary factors, air pollution and smoking. We examined regional motion traits for smoking and air pollution showing that current smokers had larger cavity volumes and impaired diastolic relaxation compared to controls with stronger effect sizes compared to former smokers (Figure 10, Supplementary Figure 10 and Supplementary Table 3). We also observed dose dependent associations on motion from air pollution levels with a similar pattern to smoking. Air pollution is related to ventricular dilatation^25^, potentially due to oxidative stress, and here we extend the associated phenotypes to altered contractile function and diastolic relaxation.

**Figure 9.**
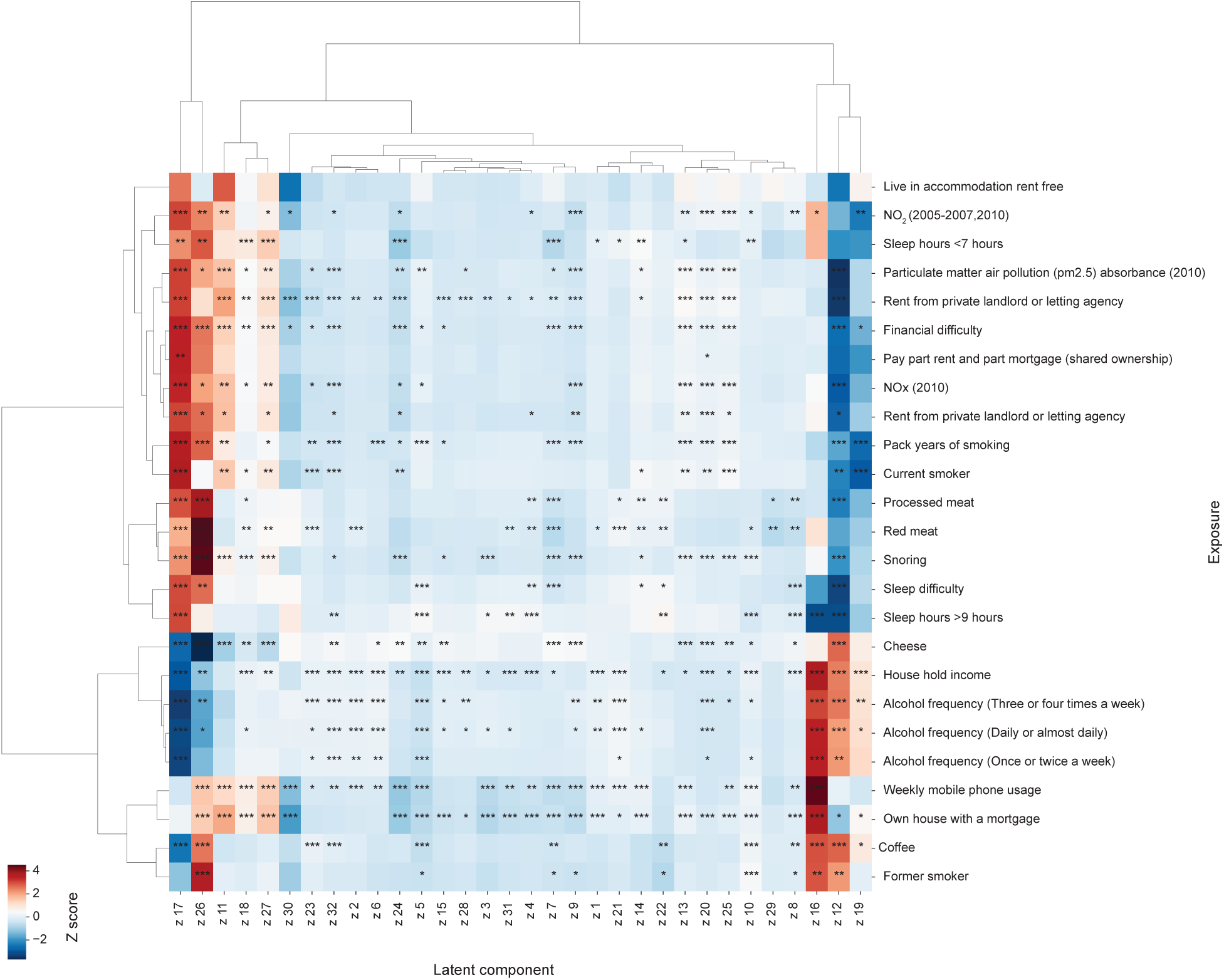
Environmental exposure–motion latent component associations. Hierarchically clustered heatmap depicting associations between 25 environmental factors from the UK Biobank—each previously linked to increased cardiovascular risk—and motion latent components. For categorical variables, dummy variables were derived and analysed separately. Asterisks denote FDR-significant associations (^∗^FDR ≤ 0.05, ^∗∗^FDR≤ 0.01, ^∗∗∗^ FDR ≤ 0.001). Colors indicate standardised beta values (Z scores). FDR, false discovery rate; NO_x_, Nitric oxides; NO_2_, Nitric dioxide.

**Figure 10.**
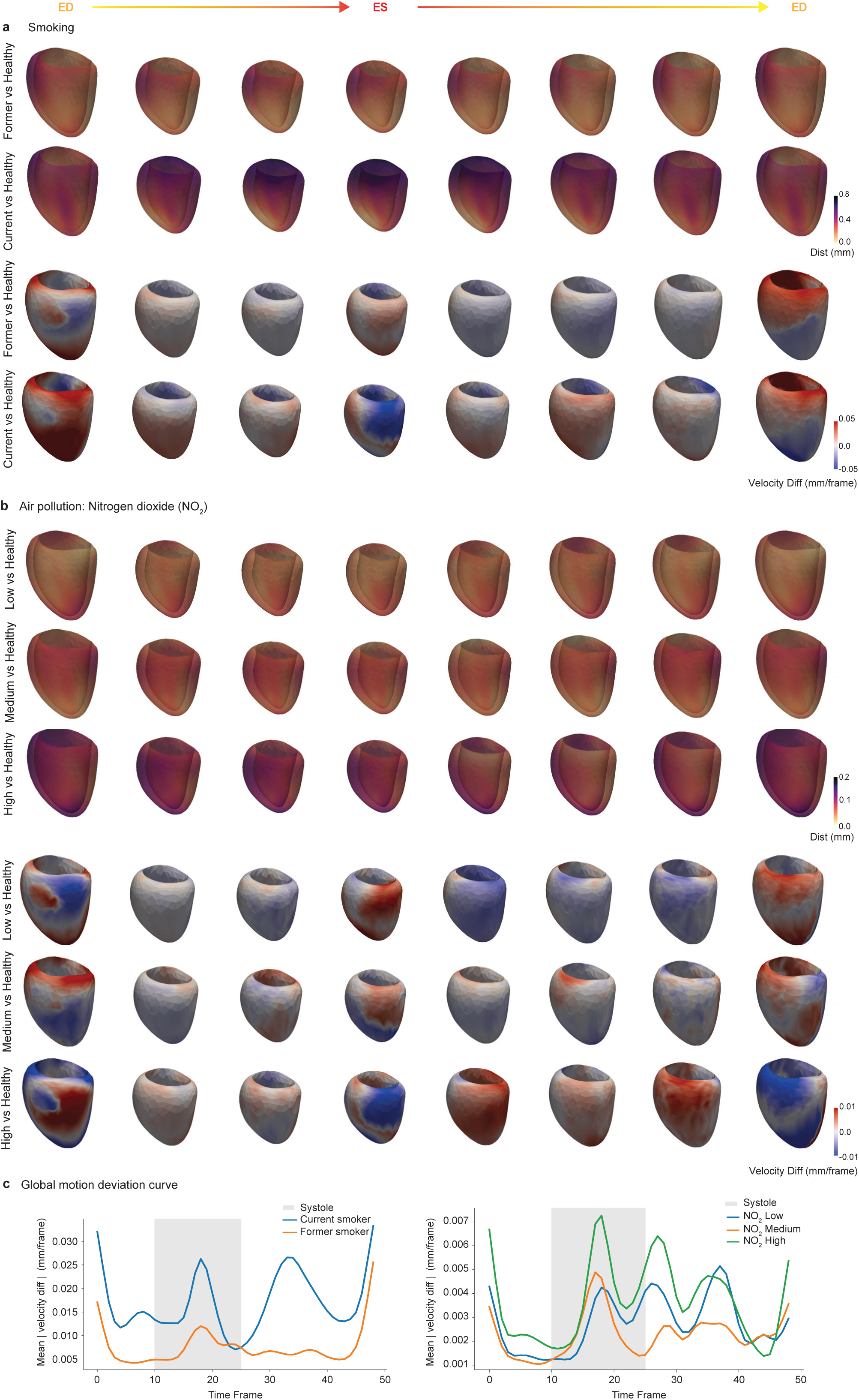
Spatiotemporal differences in left ventricular deformation and motion across environmental exposure groups. **a**, Smoking and **b**, Air pollution (nitrogen dioxide). For each exposure group, Local displacement magnitude maps (Dist in mm) showing pointwise deviations in average left ventricular (LV) geometry relative to the control group across the cardiac cycle. Warmer colours indicate regions of greater displacement, reflecting more pronounced deformation relative to controls. Corresponding maps of local myocardial velocity differences (Velocity Diff in mm/frame) between exposure groups and controls. Red regions indicate faster motion and blue regions indicate slower motion compared with the reference. **c**, Global motion deviation curves showing the average magnitude of vertexwise velocity differences (mm/frame) between each exposure group and the control cohort.

### Learning a taxonomy of cardiac motion traits and biological relevance

Cardiovascular disease classification based on consensus cutoffs does not capture the underlying diversity of pathophysiological mechanisms. Phenotypic expression and adverse risk can instead be represented across continuous axes of variation as clinical states diverge from health. We therefore applied Discriminative Dimensionality Reduction via Learning a Tree (DDRTree)^26^ to the 32-dimensional latent motion representations from *Cardio4D-VAE*, jointly performing dimensionality reduction and unsupervised clustering by mapping each subject to *c*_1_ and *c*_2_ coordinates on a two-dimensional tree-structured manifold (see Methods). The model was trained on 36,856 and validated on 41,257 participants, enabling assessment of stability and reproducibility on unseen data^27,28^.

The tree identified five main phenotypic branches, two of which further subdivided (Figure 11; latent and conventional trait mappings in Supplementary Figures 11 and 12). Probabilistic distributions of genetic, phenotypic, and clinical outcome variables were mapped onto the tree for both continuous and binary traits (Figure 11; Supplementary Tables 4 and 5 and Figures 13 and 14). Diastolic blood pressure and HbA1c were highest in branches 2, 3, and 5 (*P* < 10^−6^), which also showed elevated LV mass and impaired diastolic function. TTNtv were enriched in branch 2 (*P* < 0.05) and HCM P/LP variants in branch 3. Five-year MACE risk was worst in branches 2 (*P* < 10^−4^), 3, and 5 (each *P* < 10^−3^), with lowest risk in branches 6 and 7 (Supplementary Figure 15). Together, these findings illustrate how population diversity in complex motion traits can be efficiently represented, and visualise the interplay between genetic and cardiometabolic risk on outcomes.

**Figure 11.**
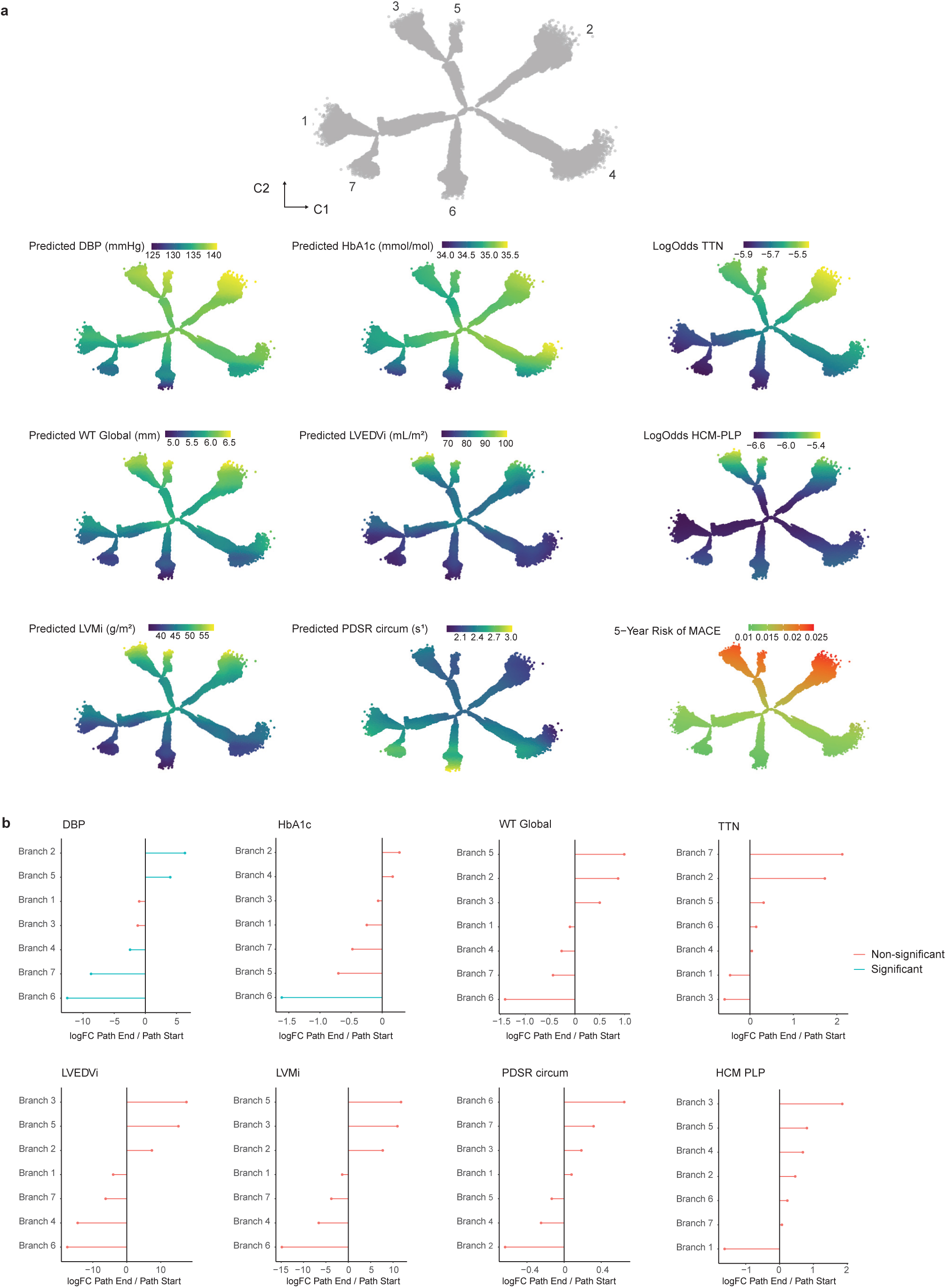
Visualisation of 4D cardiac motion heterogeneity via DDRTree mapping. 32-dimensional latent components (*Z*) were reduced to a 2-dimensional tree structure (*c*) where each point represents one individual. **a**, Each participant in the tree is colored by the model-predicted probability (log-odds) of carrying a pathogenic or likely pathogenic rare variant (TTN, HCM), by the predicted values of continuous cardiac phenotypes (HbA1c, diastolic blood pressure, LVEDVi, LVMi, wall thickness and circumferential PDSR), and the 5-year survival-derived risk of major adverse cardiac events (MACE) estimated from a Cox proportional hazards model. (Statistical branch comparison results are presented in Supplementary Figures 13 and 14). **b**, The differential expression of different clinical, imaging, and outcome phenotypes along each branch. Branches with a significant enrichment. (LogFC change from base to end) of branches are highlighted in blue. Directional changes of latent features along pseudotime for each branch shown in Supplementary Figure 11. DBP, diastolic blood pressure; HbA1c, glycated haemoglobin; HCM, hypertrophic cardiomyopathy; LVEDVi, left ventricular end-diastolic volume indexed; LVMi, left ventricular mass indexed; MACE, major adverse cardiac event; PDSR circum, circumferential peak diastolic strain rate; WT Global, global wall thickness.

In exploratory prognostic analyses, motion-derived coordinates also captured information not fully reflected by conventional imaging markers (see Supplementary Methods). Coordinates *c*_1_ + *c*_2_ alone modestly outper-formed LVEF for 5-year MACE prediction (C-index 0.6014 vs. 0.5810), and combining motion features with LVEF gave the best discrimination (C-index 0.6179; ΔAUC = 0.038, *P* = 4.2 × 10^−6^). Similar patterns were seen for models anchored on LVEDVi (ΔAUC = 0.040, *P* = 5.2 × 10^−6^) and Ell global (ΔAUC = 0.032, *P* = 1.46 × 10^−4^) (Supplementary Table 6). Among the motion coordinates, *c*_2_ carried most of the prognostic signal and remained associated with MACE after adjustment for clinical covariates and either LVEF or global longitudinal strain (Supplementary Table 7; Figure 12). These results should be interpreted as support for biological relevance and complementary risk stratification, rather than evidence of clinical utility.

**Figure 12.**
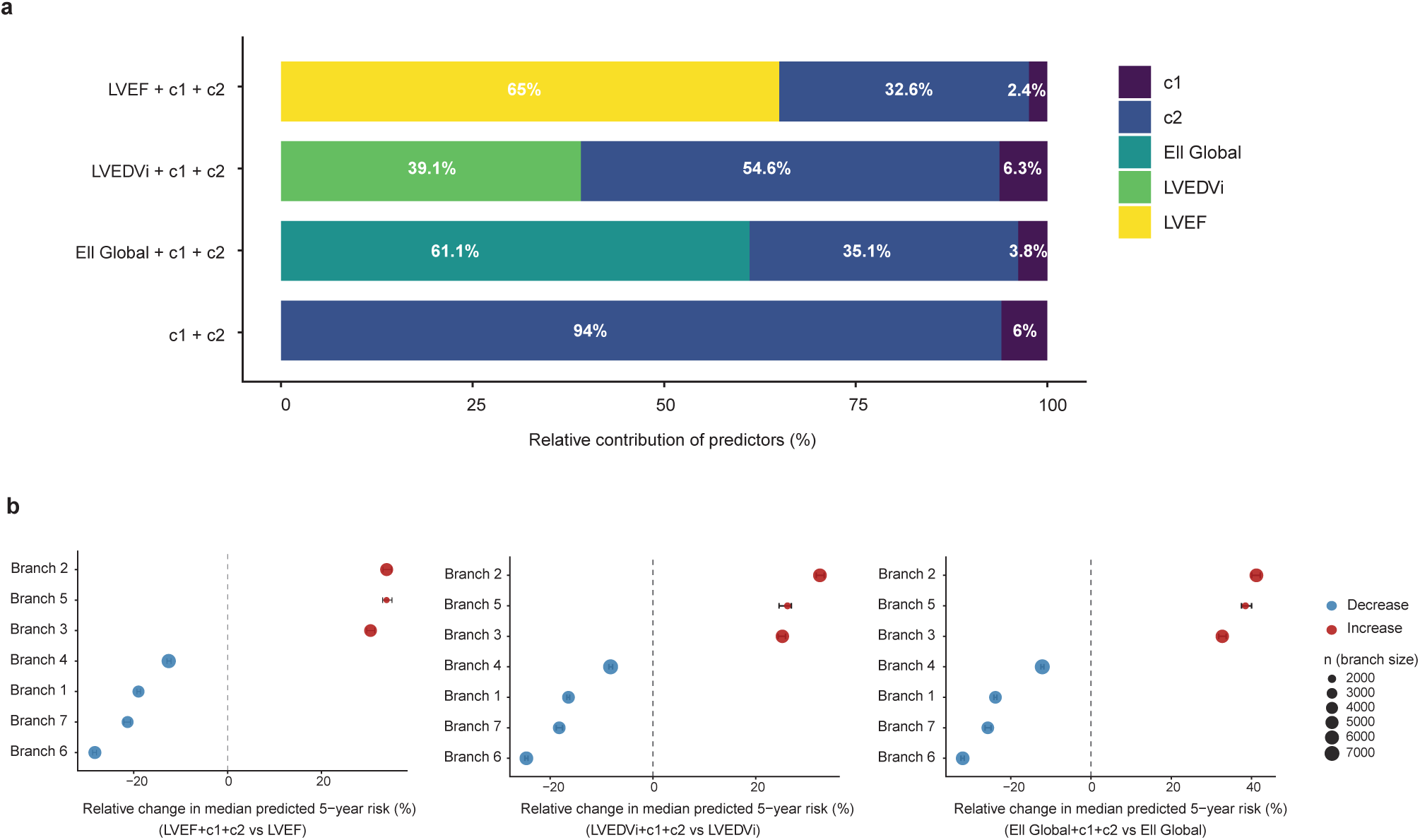
Comparison of prognostic contribution and risk stratification across motion-derived and conventional imaging features. **a**, Relative contribution of individual predictors to prognostic performance. Bars show the fraction of the total prognostic signal contributed by each predictor, quantified using partial Wald *χ* ^2^ statistics from Cox regression models. Motion-derived latent components contribute substantially to model performance, even when conventional imaging traits are included. **b**, Relative change in median predicted 5-year MACE risk across motion-defined branches, after adding motion-derived latent features (*c*1, *c*2) to Cox models using conventional imaging traits. Each panel shows the relative change in median predicted 5-year MACE risk within branches after adding motion-derived latent coordinates (*c*1, *c*2) to a conventional Cox model based on a single imaging biomarker (left: LVEF; middle: LVEDVi; right: Ell Global). Points represent branches, with size proportional to branch sample size. Positive values indicate increased predicted risk following incorporation of motion features, while negative values indicate attenuation of risk. Motion-derived features systematically increased predicted risk in high-risk branches (e.g. branches 2, 3, and 5) and reduced predicted risk in lower-risk branches, demonstrating branch-specific refinement of risk stratification beyond conventional prognostic models.

The learned tree embedding generalised well to unseen participants, with preserved structure across datasets (trustworthiness: train = 0.880, test = 0.877) and closely matched coordinate distributions (Wasserstein < 0.05; JS divergence < 0.004; Supplementary Table 8 and Figure 16).

## Discussion

Ventricular motion reflects the integration of myocardial fibre orientation, electromechanical coupling, developmental patterning, and biophysical constraints imposed by loading conditions. Conventional metrics such as ejection fraction and global strain aggregate spatially heterogeneous processes into scalar measurements that may obscure heritable regional traits and patterns of coordinated function. Realising the full phenotypic content of cardiac motion requires approaches that preserve this spatiotemporal complexity across the genome, the exposome, and the spectrum of cardiovascular risk.

Genome-wide association of the 32 latent motion traits identified 39 loci, most undetected by conventional metrics and only 6 shared by static 3D traits^29,30^. Shared loci recapitulated canonical cardiomyopathy genes involved in sarcomeric assembly (*TTN*, *MYBPC3*, *MYH7*) and excitation-contraction coupling (*PLN*, *SCN5A*). Several effector genes were supported by multiple lines of genomic evidence, implicating microtubule organisation (*MAST2*)^31^, stress-induced cardiomyocyte pyroptosis (*TRIB2*)^32^, epigenetic regulation of myocyte ageing (*MLF1*)^33^, and mitochondrial bioenergetics (*GFM1*)^34^. Latent-only loci implicated secreted modulators of fibrillin-rich extracellular scaffolds (*ADAMTSL3*, *ADAMTS18*, *ADAMTS6*, *EFEMP1*; Supplementary Figure 8e), pointing to ventricular stiffness and elastic recoil^35^, alongside transcriptional regulators of chamber specification and conduction-system architecture (*TBX20*, *TBX3*, *NKX2-5*, *HAND2*)^36,37^. TWAS in adult left ventricle and atrial appendage corroborated these gene-level assignments, including *CD36*, a fatty-acid transporter whose nonsense variant accounts for approximately 20% of excess dilated cardiomyopathy risk in African-ancestry populations^17^. Integration with single-nucleus RNA-seq identified a cardiomyocyte core and an additional non-myocyte axis spanning endocardial, endothelial, and lymphoid lineages for latent-only genes.

Variant-level fine-mapping integrated with a 90-cell-state foetal heart regulatory map sharpened divergence between trait domains: conventional traits localised to cardiomyocyte and conduction-system elements, whereas motion traits redistributed toward fibroblast, endocardial, autonomic-neuronal, and endothelial programmes. A hierarchical framework integrating enhancer-to-gene links, protein-altering annotation, locus-to-gene scores, and pan-tissue regulatory evidence resolved credible-set variants to candidate effector genes (Table 2). The highest-confidence motion-specific signals converged on developmental regulators of cardiac morphogenesis. A prominent signal mapped to an atrial-cardiomyocyte promoter for *CCND2*, a cardiogenic GATA4 cofactor whose overexpression drives cardiomyocyte proliferation in injury models^38^. A second signal linked *CDK15* to an enhancer active in second-heart-field-derived fibroblasts, with murine knockout showing increased heart weight^39^. Additional signals included a missense variant in *CRIP1*, mediating epicardial epithelial-to-mesenchymal transition, and a neural-crest enhancer at *SLC2A12*/*TCF21* coinciding with a coronary heart disease allele implicated in ventricular septal defects^40^. This convergence is consistent with an oligogenic model in which common variants collectively contribute to structural and functional phenotypes through shared developmental programmes.

External analyses corroborated this regulatory interpretation beyond the UK Biobank imaging cohort. GPMap linked motion-associated loci to developmental and non-myocyte molecular programmes, while FinnGen supported 40 of 87 fine-mapped loci across developmental, valvular and conduction disease modules. Shared loci showed the broadest cardiac disease associations, whereas motion-only loci were less dominated by myocardial-remodelling phenotypes than conventional-only loci. In All of Us, the ancestry-matched EUR analysis supported 18 loci at FDR significance, with closely concordant findings in the cross-ancestry META analysis. Together, these results connect the fine-mapped motion loci to independent molecular and disease contexts consistent with developmental, stromal, valvular and conduction mechanisms.

Rare titin truncating and pathogenic sarcomeric variants induced spatially heterogeneous alterations in contraction timing and diastolic relaxation velocity, demonstrating that variably penetrant genetic variation disrupts regional motion coordination before geometric remodelling is detectable^20,41^. Environmental risk factors also imprinted distinct motion signatures orthogonal to traditional metrics^25^. Smoking and air pollution impaired diastolic relaxation and increased cavity volumes dose-dependently, consistent with oxidative stress-mediated dysfunction^42^. Alcohol and coffee intake mapped to similar motion profiles, likely reflecting shared pharmacological effects^43,44^, while mobile phone use showed associations suggesting links between psychosocial stress and subclinical cardiac remodelling^45,46^.

Unsupervised tree-based clustering revealed a branching taxonomy with seven phenotypic trajectories, differentially enriched for pathogenic variants, risk factors, and future major adverse cardiovascular events. This structure represents cardiac function along continuous pathways of differentiation rather than isolated diagnostic categories, placing variant carriers and at-risk individuals within a broader phenotypic landscape that quantitatively links genetic burden, loading conditions, and metabolic status to clinical outcomes^27^. The motion-based taxonomy provided incremental prognostic value beyond conventional imaging biomarkers, offering support for the biological relevance of the latent features. However, unsupervised learning would not be expected to produce a model optimised for outcome prediction.

Together, this work demonstrates that 4D cardiac motion encodes heritable and environmentally responsive features whose regulatory architecture extends into non-myocyte compartments under-represented by conventional phenotyping. Rare variants manifest through spatiotemporal disruption before geometric remodelling is detectable, and motion-derived phenotypes carry prognostic information, supporting the biological relevance of the latent feature space. These findings demonstrate the beating heart as a phenotypic substrate for genetic discovery: one whose molecular regulation spans sarcomeric, developmental, and stromal programmes, and whose population-level variation reflects the joint effect of common variants, rare pathogenic alleles, and modifiable exposures. By integrating computer vision, deep learning, and population genomics, this work provides a framework for extending genetic discovery into a wider spectrum of dynamic cardiac function.

## Methods

### UK Biobank participants

UK Biobank is a large cohort study of half a million individuals in the UK aged 40-69 at recruitment between 2006 and 2010^47^. All participants provided written informed consent for the study, which was also approved by the National Research Ethics Service (11/NW/0382). Our study was conducted under terms of access approval number 40616. A range of phenotypic and genotypic attributes, risk factors and physical measures was used for the analyses. These were collected by touch screen questionnaires, interview, biophysical measurement, hospital episode statistics, and primary care data. CMR images were also acquired for a subset of participants. Details of how each phenotype was acquired are available on the UK Biobank Showcase (http://biobank.ctsu.ox.ac.uk/crystal/).

### Exposures and outcomes

The effect of motion traits on clinical outcomes was assessed by using lifetime risk. The UK Biobank reports the date of first occurrence of a diagnosis, identified from self-reporting, primary care, hospital in-patient, and death register records. This permitted the identification of events preceding recruitment to the UK Biobank.

The primary clinical outcome was a composite of all-cause mortality or major adverse cardiovascular events (MACE) defined as the occurrence of stroke, myocardial infarction (MI), heart failure (HF), cardiac arrest, and cardiovascular mortality. Secondary clinical outcomes were the individual components of the primary clinical outcome. Using Office for National Statistics data (ONS), any death with a primary cause ICD-10 code beginning with ‘*I*’ was classified as cardiovascular. A full list of endpoint definitions and data fields used from the UK Biobank database is in Supplementary Table 9. Outcomes were classified as prevalent or incident if they happened before or after the imaging visit. We could not capture exposure dynamics across the life course, since all exposures were only assessed at one time point in the full UKB cohort. We also have not captured all possible exposures as we were limited to the data available.

### Exposure variables

To evaluate the role of environmental exposures on heart motion, we followed the pipeline proposed by Argentieri et al.^48^. We selected 17 environmental factors related to smoking habits, alcohol use, diet, pollution, and income, all known to be associated with adverse cardiovascular events (Supplementary Table 10)^21^. Variables with fewer than 45% missing values and available across all assessment centers were included. All continuous exposure variables were centered and standardised prior to analysis. Ordinal categorical variables were recoded to test only linear associations, whereas all nominal categorical exposures were analysed using the most common category as the reference. Each level of the variables related to alcohol frequency, sleep duration, housing, and financial difficulty was converted into a dummy variable and analysed separately, resulting in a total of 25 variables. All “mark all that apply” questions were recoded as binary dummy variables. Further details on the preprocessing of exposure variables are provided in the Supplementary Methods.

### Interaction between environmental factors and complex motion traits

Multivariable linear regression analyses were performed with each latent feature as the dependent variable and regressed against 25 derived exposure factors, one at a time. Each model was adjusted for body surface area (BSA), sex, age, and ethnicity (White, Asian, Black, Mixed, or Other). For each model, *β* coefficients were estimated for each exposure-latent feature pair. P values were corrected for multiple testing using the false discovery rate (FDR) according to the Benjamini–Hochberg method^49^, with statistical significance defined as FDR-adjusted at *q* < 0.05.

### Cardiomyopathy-associated rare variant classification

Cardiomyopathy-associated rare variants were identified as previously published^50^. Individuals were classified as genotype-negative if they had no rare protein-altering genetic variation (minor allele frequency <0.001 in the UK Biobank and the Genome Aggregation Database) in any genes that may cause or mimic HCM. These genes represented an inclusive list of genes with definitive or strong evidence of an association with cardiomyopathy, moderate evidence, and genes associated with syndromic phenotypes^51,52^. This group was compared with individuals with disease-associated rare variants in genes with strong or definitive evidence for HCM (*MYBPC3, MYH7, MYL2, MYL3, TNNI3, TNNT2, TPM1*, and *ACTC1*). Analysis was restricted to robustly disease-associated variant classes for each gene and to variants sufficiently rare to cause penetrant disease (filtering allele frequency <0.00004 for HCM^53^). Variants were classified as pathogenic/likely pathogenic if reported as P/LP for cardiomyopathy in ClinVar and confirmed by manual review. Predicted loss-of-function variants in *TTN* were curated to MAF < 0.00004 in gnomAD and UK Biobank. Only TTNtvs in ‘per cent spliced in’ (PSI) ratio > 90% exons predicted to influence gene product level (e.g. frameshift, essential splice, stop gained) were kept. Compound heterozygous carriers of common TTNtvs (same two TTN variants identified in > 10 individuals) were removed from the analysis. The variant list only included variants that would be called P/LP if identified in a patient with DCM, and the variants were manually curated if they had any evidence of pathogenicity.

### Conventional cardiac image segmentation and strain analysis

CMR imaging was performed on participants to capture two-dimensional retrospectively-gated cine imaging on a 1.5T magnet (Siemens Healthineers, Erlangen, Germany)^54^. Left ventricular short-axis plane cine images from base to apex were acquired as well as long-axis cine images in the two and four-chamber views. Cines comprised 50 cardiac frames with a typical temporal resolution of 31 ms. Ascending and descending thoracic aorta images were also acquired with transverse cine imaging.

The two-dimensional cine images in the short and long-axis were segmented using fully convolutional networks, with segmentation quality equivalent to expert human readers^10^. Derived measurements of the left and right ventricles were determined from the two-dimensional images: end-diastolic and end-systolic volumes, stroke volume, and ejection fraction. A density of 1.05 g.ml^-1^ for the left ventricular myocardium was used to derive its mass from its volume. With *A*_2*C*ℎ_ and *A*_4*C*ℎ_ as the atrial areas on the two and four-chamber cine views respectively, and *L* as the averaged longitudinal diameter across two views, the atrial volumes were derived as per the following formula on biplane area-length: 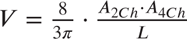. CMR-derived measurements were indexed to body surface area (BSA) calculated with the Du Bois formula: BSA = 0.007184 * Height * Weight^0.425^ with height in cm and weight in kg. The wall thickness of the left ventricular myocardium was measured as the distance between the segmented epicardium and endocardium at end-diastole.

Circumferential and radial strains were calculated with cine short-axis images as per the formula 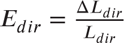 with *dir* the circumferential or radial direction, *L*_*dir*_ the length of a line segment in this direction and Δ*L*_*dir*_ the change of length over time. Longitudinal strain was derived from long-axis four-chamber motion tracking.

A spatio-temporal neural network was used to segment the aorta cine images^55^. Maximum and minimum cross-sectional areas were derived and aorta distensibility was calculated with central blood pressure obtained using peripheral pulse-wave analysis (Vicorder, Wuerzburg, Germany)^56^.

### DeepMesh four dimensional motion models

The left ventricle is represented as a three-dimensional (3D) surface mesh that includes both the endocardial and epicardial surfaces. We adopt a deep learning framework (*DeepMesh*^4^) to estimate 3D cardiac motion on this mesh from multi-view 2D cine cardiac magnetic resonance (CMR) images, including short-axis stacks and long-axis views (2-chamber and 4-chamber). Unlike traditional image-space methods that estimate dense pixel-or voxel-level displacements, *DeepMesh* operates directly in mesh space and learns per-vertex displacements, preserving one-to-one vertex correspondence across time and subjects. This mesh-based formulation avoids interpolation artefacts when projecting image-based motion fields onto anatomy, simplifies population-level analysis, and enables direct computation of clinically relevant volumetric and regional functional biomarkers on anatomically consistent meshes.

The *DeepMesh* framework consists of two stages: (i) Reconstruction of a subject-specific end-diastolic mesh from a population template; and (ii) Estimation of per-vertex motion across the cardiac cycle by deforming the end-diastolic mesh over time. Both stages are supervised using a differentiable rasteriser that projects the evolving 3D mesh into soft 2D contours on short-axis and long-axis planes, enabling the use of standard 2D annotations for training. The inclusion of long-axis views in this supervision is critical to improving motion estimation along the heart’s long axis. In the first stage, a subject-specific end-diastolic mesh is reconstructed by deforming a fixed-topology population template mesh. A convolutional neural network processes the multi-view end-diastolic images to predict a 3D voxel-wise displacement field. This field is sampled at the template’s vertex locations to produce the deformed mesh for that subject. As a result, all end-diastolic meshes are topologically consistent (i.e., share the same vertex and face structure) and directly comparable across individuals.

In the second stage, *DeepMesh* estimates the left ventricle motion over time by predicting per-vertex displacements from the end-diastolic frame to any time point t in the cardiac cycle. A motion network processes the multi-view images images at the end-diastolic frame and frame t, and predicts a 3D voxel-wise motion field Φ_0→*t*_ that captures how the myocardium deforms over time. Sampling this field at the end-diastolic mesh vertex locations yields the per-vertex displacement Δ*V*_0→*t*_, which is used to deform the end-diastolic mesh to the mesh at time t: 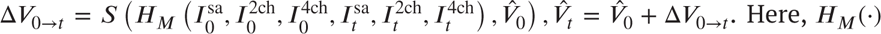 denotes the motion network, which outputs the intermediate motion field Φ_0→*t*_, and *S*(⋅) samples it at the mesh vertices. This results in a time-resolved sequence of anatomically consistent meshes that capture dynamic left ventricle motion across the entire cardiac cycle.

To supervise training using standard 2D annotations, *DeepMesh* employs a differentiable rasteriser that projects 3D meshes into 2D probability maps representing myocardial contours. The rasteriser slices the predicted mesh along the short-axis and long-axis planes and computing each vertex’s likelihood of lying on the image plane based on its distance to the plane. The resulting 2D soft contours are then compared to ground-truth 2D annotations using a weighted Hausdorff distance, a robust loss function that handles sparse or noisy annotations. This rasterisation strategy provides strong supervision from all three anatomical views. In particular, the short-axis view contributes dense in-plane anatomical coverage across slices, while the long-axis views provide high-resolution through-plane constraints that are critical for accurate reconstruction and motion estimation along the long axis.

In addition to the contour-based loss, model training incorporates auxiliary constraints to ensure anatomical plausibility and motion coherence. Laplacian smoothness promotes surface regularity, Huber loss encourages coherent motion fields, and a image similarity loss provides self-supervision by enforcing consistency between warped and reference short-axis stack. All losses are fully differentiable and jointly optimised in an end-to-end manner, enabling robust learning from multi-view cine CMR using only standard 2D annotations.

*DeepMesh* was trained and evaluated on 530 subjects from the UK Biobank. On the challenging end-diastolic to end-systolic transition, it achieved state-of-the-art motion tracking performance, particularly along the long axis. On the 2-chamber view, the method reached a Hausdorff distance of 5.75 ± 1.81 mm and a Boundary F-score of 89.26%, while on the 4-chamber view, it achieved 6.21 ± 2.56 mm and 88.69%, respectively. These results substantially outperform existing techniques, including non-rigid image registration (e.g., 15.17 ± 4.52 mm / 77.6% on 2-chamber view and 15.95 ± 4.84 mm / 77.79% on 4-chamber view), demonstrating the benefits of explicit multi-view image fusion and supervision.

DeepMesh offers state-of-the-art performance on cardiac motion estimation through combining multiple views of the heart and using direct image-to-mesh generation. Residual errors due to image quality, mis-registration, and displacement estimation may propagate into the modelled traits.

After model training, *DeepMesh* analysed CMR data from participants in UK Biobank. For each, the model generated 50-frame sequences of left ventricular meshes, all with identical vertex count across time and individuals.

### Data pre-processing

As a preprocessing step, each subject mesh’s end-diastolic (ED) frame was rigidly aligned to the atlas via symmetric rigid surface registration implemented in the Medical Image Registration ToolKit (MIRTK). The estimated transform was applied to all frames, yielding temporally consistent atlas-space meshes while preserving anatomical geometry. This atlas was defined from over 1,000 healthy adults which enforced spatial and temporal consistency^57^. Meshes were then down-sampled using surface simplification using quadric error metric^58^, from 22,043 to 1,200 vertices. Following pre-processing and quality control (QC), subjects with one or more missing frames (*T* < 50) were excluded from all downstream analyses. To identify outliers and failure cases in mesh data, we extracted subject-level features using *Cardio4D-VAE* model (described below). The embeddings were projected to two dimensions for visualisation. A k-nearest-neighbours (k=5) distance was computed for each subject to quantify its local neighbourhood structure in the latent space. A percentile threshold of this distance distribution was then used as a screening criterion to identify candidate outliers for visual quality control.Candidate cases were subsequently inspected visually, and exclusions were restricted to subjects with clear mesh-generation failures (e.g., collapsed meshes, and holes resulting from failures in the upstream mesh-generation pipeline). After all quality control steps, 78,113 participants were retained for analysis.

### Cohort-level comparison of ventricular shape and motion

To assess cohort-level differences in cardiac shape and motion, we performed a groupwise comparison of left ventricular dynamics. For each cohort, we generated a spatiotemporal (4D) average mesh of left ventricular motion by averaging meshes across subjects at each phase of the cardiac cycle. The absolute local distance of each cohort’s average left ventricular (LV) surface relative to the non-carrier reference cohort was then computed at each vertex and cardiac frame as:

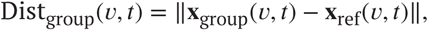

where **x**_group_(*v*, *t*) and **x**_ref_(*v*, *t*) denote the 3D coordinates of vertex *v* at frame *t* for the group-averaged and reference meshes, respectively. This metric captures regional shape deviations throughout the cardiac cycle (e.g., Figure 8a)

For each subject group, framewise myocardial velocity fields Vel_group_(*v*, *t*) were computed as the absolute pointwise displacement per frame interval:

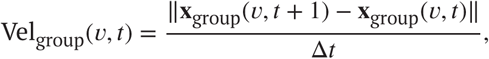

where *v* indexes mesh vertices and Δ*t* = 1 frame. The resulting fields capture the temporal evolution of myocardial motion throughout the cardiac cycle.

To visualise regional motion deviations relative to the reference cohort, vertexwise velocity differences were computed as

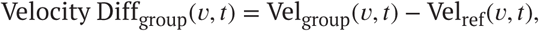

where Vel_ref_(*v*, *t*) denotes the reference group’s velocity at vertex *v* and frame *t*. Positive values indicate faster motion and negative values slower motion relative to the reference. Representative maps (e.g., Figure 8b) highlight spatially localised differences in ventricular wall motion across the cardiac cycle.

To summarise global myocardial motion dynamics, the spatially averaged velocity magnitude for each group was computed as

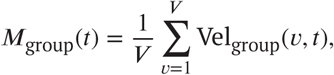

yielding a group-specific temporal profile of motion amplitude over the cardiac cycle. This representation captures overall motion intensity and timing rather than spatial deviations (e.g.,Figure 8d).

### Unsupervised latent representation of cardiac motion with Cardio4D-VAE

As illustrated in Figure 1, the proposed *Cardio4D-VAE* model is a probabilistic generative model trained in an unsupervised learning setting to capture a compact latent representation of dynamic left-ventricular motion. The extracted left-ventricular mesh sequences from *DeepMesh* framework, can be represented as *M*_0∶*T* −1_ = *m*, *m*,…, *m* where *T* denotes the number of time frames. Each LV mesh *m_t_* = (*X_t_*, *A_t_*) is a graph with a node feature matrix *X*_*t*_ = [*x*_*t*0_, *y*_*t*0_, *z*_*t*0_]…[*x*_*tV* −1_, *y*_*tV* −1_, *z*_*tV* −1_] ∈ ℝ^*V* ×3^ and *A*_*t*_ ∈ {0, 1}^*V* ×*V*^ is the mesh adjacency matrix. Atlas-based spatial alignment yields a fixed adjacency matrix *A* across subjects and time frames, ensuring consistent vertex-to-vertex correspondence.

Given the dynamic 3D left-ventricular mesh sequences, our objective is to train a model that learns a probabilistic low-dimensional representation capturing both anatomical and temporal variations, through an unsupervised learning framework based on a variational autoencoder (*Cardio4D-VAE*). Mesh sequences were then projected into this latent space, yielding compact shape descriptors (latent variables) for each cardiac cycle. We adopt a *β*-VAE formulation^59^ to encourage a compact and interpretable latent space. In this setting, the KL term is weighted by an adjustable coefficient, promoting disentanglement while not enforcing strict independence among latent dimensions. This flexibility is appropriate for cardiac physiology, where ventricular motion patterns are biomechanically coupled. The resulting latent space therefore captures coordinated modes of left-ventricular deformation while allowing physiologically meaningful dependencies to persist.

In this framework, the encoder *E_**θ**_* ∶ ℝ^3*VT*^ → ℝ^*d*^ maps the input dynamic meshes into a latent space, while the decoder *D_θ′_* ∶ ℝ^*d*^ → ℝ^3*VT*^ reconstructs the meshes, with ***θ*** and ***θ***^′^ representing the learnable parameters. Inspired by prior advances in spatiotemporal representation learning^60^, we designed an architecture that jointly captures anatomical structure and temporal dynamics across the cardiac cycle. The model employed a convolutional encoder composed of hierarchical three-dimensional residual blocks with batch normalisation and rectified linear (ReLU) activations, capturing localised deformation patterns across spatial and temporal dimensions. Input data consisted of voxelized representations of dynamic surface meshes, enabling efficient learning within a structured volumetric domain. Spatial and temporal resolution were progressively reduced through strided convolutions to extract multiscale features. The resulting feature map was aggregated by global average pooling and projected to the parameters of a multivariate Gaussian posterior, yielding mean *μ* ∈ ℝ^*d*^ and log-variance log *σ*^2^ ∈ ℝ^*d*^ vectors that define the approximate distribution *q*(*Z* ∣ *X*) =  (*μ*, diag(*σ*^2^)). Latent samples *Z* ∈ ℝ^*d*^ were obtained via the reparameterisation trick, *Z* = *μ* + *σ* ⊙ *ε*, with *ε* ∼  (0, *I*), enabling differentiable stochastic sampling during training.

Finally, latent vectors were decoded using a lightweight fully connected network consisting of two linear layers with a sigmoid activation function. The decoder generated dynamic left-ventricular surface meshes, providing anatomically interpretable reconstructions of cardiac motion from the latent space. This generative formulation enabled compact, low-dimensional representations of population-level deformation patterns.

To enforce spatiotemporal geometric and physiological consistency, the model was trained using a combined loss function:

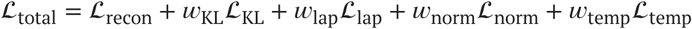

where *w*_KL_, *w*_lap_, *w*_norm_, and *w*_temp_ are weighting coefficients that balance the contribution of each regularisation term. 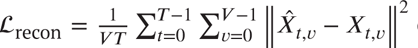 denotes the mean squared error between the reconstructed and original vertex coordinates, with *X̂*_*t*_ representing the decoded mesh coordinates at time frame *t*. The term G_KL_ corresponds to the Kullback–Leibler divergence between the approximate posterior *q*(*Z* ∣ *X*) and an isotropic unit Gaussian prior *p*(*Z*) =  (0, *I*), as in the evidence lower bound (ELBO) formulation of variational autoen-coders. The Laplacian loss G_lap_ serves as a regularisation term that enforces coherent move among neighboring vertices, thereby reducing mesh self-intersections and promoting smoother surface reconstructions^61^. It penalises the deviation of each vertex position from the mean of its neighbors. In addition, the normal consistency loss G_norm_ encourages smooth alignment of surface normals across adjacent faces. Following the PyTorch3D implementation, this is computed as the cosine similarity between the normals of neighboring triangles, ensuring that reconstructed meshes preserve locally smooth and realistic surfaces. To enforce frame-to-frame motion smoothness, we define the temporal smoothness loss as 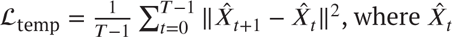 denotes the decoded mesh coordinates at time frame *t*..

### Experimental settings

Model development was performed using a randomly selected cohort of 10, 000 UK Biobank participants. To ensure generalisability across datasets, we applied a random split strategy, allocating approximately 80% of the data to training/validation and 20% to testing. Training includes 7000 samples, validation 1000, and held-out test 2000 sets. Reconstruction accuracy was assessed on the test set using the root mean squared error (RMSE) between the predicted and ground-truth meshes in physical space, yielding RMSE = 2.14 ± 0.38 mm (*mean* ± *SD*).

For each participant *i*, motion data were represented as a tensor *X*_*i*_ ∈ ℝ^*T* ×*V* ×*F*^, where *T* is the number of time frames, *V* the number of nodes, and *F* the number of features per node. In our setting, *T* = 50, *V* = 1200, *F* = 3. We applied dataset-level min-max normalisation to preserve relative shape size across subjects and time. Let

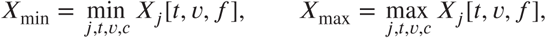

computed once over the entire training cohort {*X*_*j*_ } and subsequently applied unchanged to the held-out test and remaining UKB cohort. Each sample was normalised as

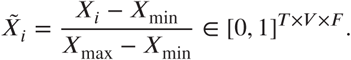

This pre-processing allows the preservation of intra-population size variability while translating all data points between 0 and 1.

Training was performed with a batch size of 32 and a learning rate of 10^−4^. All experiments were implemented in Python v3.8.10 using PyTorch v1.13.0 (CUDA 11.7) and PyTorch Geometric v2.5.3^62^, model training was performed on an NVIDIA Tesla V100 GPU with 32 GB of memory. The model was trained for 30 epochs using the Adam optimiser^63^ with a learning rate of 10^−4^.

### Hyperparameter optimisation

To optimise the weighting coefficients of the loss terms *w*_KL_, *w*_lap_, *w*_norm_, *w*_temp_ and the latent dimensionality of the encoder, we used Bayesian hyperparameter search with Optuna v4.0.0^64^. Each trial sampled *w*_KL_, *w*_lap_, *w*_norm_ and *w*_temp_ from a log-uniform distribution in the range [10^−5^, 1] and selected the latent dimensionality *d* from {16, 32, 64, 128}. For each candidate configuration, the model was trained and evaluated on the validation set, and the final validation loss was used as the optimisation objective. To reduce sensitivity to model initialization, the optimisation was performed across five independent random seeds (1701, 1993, 2023, 42, and 1994). This strategy allowed us to automatically identify the hyperparameters that yielded the best trade-off between reconstruction accuracy, smoothness, and regularisation.

The optimal configuration resulted in weighting coefficients of *w*_KL_ = 2 × 10^−5^, *w*_lap_ = 10^−4^, *w*_norm_ = 10^−4^, and *w*_temp_ = 10^−4^, with a latent dimensionality of *d* = 32. The encoder weights were initialised from a ResNet-3D-18 backbone^60^. The decoder consisted of a lightweight fully connected network with a hidden layer of size 1024, mapping the latent representation to the input space, with ReLU activation and a final sigmoid output. The model was jointly optimised with all trainable components. Under these settings, training the model required approximately 5.2 hours.

The latent trait distributions were highly consistent between the model-development cohort and the remaining UK Biobank participants, indicating that the phenotypic variation present in the full cohort was well represented during model development. Only a small proportion of unseen participants fell outside the latent-value range observed during model development (maximum 0.12% across latent dimensions; mean 0.024%; Supplementary Figure 18 and Supplementary Table 12).

### Latent–phenotype association analysis

To quantify associations between the learned latent cardiac motion features and conventional imaging phenotypes, we employed two complementary regression frameworks. We first assessed the global association between all latent components and each imaging-derived trait using a multivariable regression model:

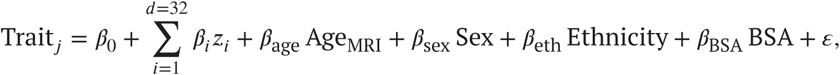

where *z*_*i*_ denotes the *i*^th^ latent component and *ε* is the model residual. This model evaluates the overall predictive power of the latent motion space for each phenotype, adjusting for demographic and anthropometric covariates. To further quantify the effect of individual motion-derived components, we fitted the following multivariable linear regression models for each component–trait pair:

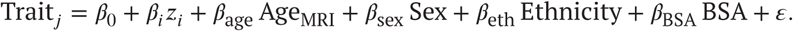

This analysis identified specific latent component (*z*_*i*_) significantly associated with structural or functional imaging traits, providing interpretable links between latent motion features and physiological phenotypes.

### Genetic quality control in the UK Biobank

Analyses were conducted in the UK Biobank resource under approved application. Participants were retained if they had no evidence of sex-chromosome aneuploidy or quality-control (QC) outlier status for heterozygosity or missingness, had complete information for autosomal phasing and principal component analysis (PCA), and passed stringent per-sample QC (genotyping call rate/Cluster_CR ≥ 0.95; derived QC score dQC ≥ 0.82). To minimise population stratification, analyses were restricted to the UK Biobank genetic ethnic grouping “Caucasian,” with full adjustment for genetic PCs in downstream models.

Cine cardiovascular magnetic resonance (CMR) was processed using a previously validated deep-learning four-dimensional pipeline that reconstructs subject-specific left-ventricular (LV) meshes across the cardiac cycle. A spatiotemporal variational autoencoder (*Cardio4D-VAE*) encoded LV motion into 32 latent features (*z*_1_–*z*_32_) that capture independent modes of spatiotemporal variation. Because the VAE objective includes a Kullback–Leibler divergence term that regularises the latents towards a standard normal prior, the resulting features were approximately Gaussian and were analysed without additional transformation. Conventional comparators were LV ejection fraction (LVEF) and global circumferential strain (Ell global), whereas indexed LV end-diastolic volume (LVEDVi) served as a structural control trait. The final analysis set comprised *N* = 65,333 participants with both imaging and genetic data passing QC.

### Genome-wide association studies

Genome-wide association studies (GWAS) were performed using REGENIE (v4.1) in its two-step ridge-regression framework^65^. In Step 1, whole-genome predictors were trained on directly genotyped array variants after variant-level QC using PLINK (v2.0)^66^: minor allele frequency (MAF) > 0.01, minor allele count (MAC) ≥ 100, missingness < 10%, and Hardy–Weinberg equilibrium *P* > 1 × 10^−15^. Per-chromosome leave-one-chromosome-out (LOCO) predictors were then generated.

In Step 2, we tested the association between imputed variants and each trait using the chromosome-specific LOCO predictors from Step 1 as covariate offsets. Genotypes were drawn from the Genomics England (GEL)-UK Biobank imputation reference panel, which combines the UK Biobank array data with the high-coverage GEL whole-genome sequencing dataset. This panel contains approximately 342 million autosomal variants imputed using data from 78,195 individuals and provides markedly improved coverage of low-frequency alleles compared with previous reference sets. We restricted analyses to autosomal variants with minor allele count (MAC) ≥ 20 and imputation INFO ≥ 0.4. Within the predominantly White British subset, the GEL reference panel demonstrates reliable imputation of variants with allele frequencies as low as 1 in 10,000, enabling the discovery of rare-variant associations while maintaining high genotype quality across the allele-frequency spectrum^67^. All models were adjusted for age at MRI, sex, age^2^, age×sex, body surface area (BSA), and the first ten genetic principal components. Because LVEDVi is already indexed to BSA, BSA was not included as a covariate for this trait. LVEF, Ell global, and LVEDVi were inverse-rank normalised prior to analysis. The 32 latent motion traits derived from *Cardio4D-VAE* were analysed on their native scale, as they are inherently approximately normally distributed due to the KL-divergence term in the variational loss that constrains the latent space towards a standard normal prior.

Prevalent cardiovascular and cardiometabolic diagnoses were not included as GWAS covariates because these heritable downstream phenotypes may mediate genetic effects on cardiac structure and function; conditioning on them can introduce overadjustment or collider bias^68,69^.

Across the 32 latents, multiplicity was controlled using an eigenvalue-based effective number of tests. Let **R** denote the latent–latent correlation matrix and {*λ*_*i*_} its eigenvalues; the effective number of independent traits was *m*_eff_ = 7. This yielded a study-wide significance threshold:

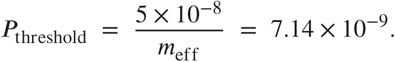

For the pan-latent minimum-*P* scan, a Manhattan plot was produced for locus visualisation. LVEF and Ell global were selected as the primary comparators because they represent the most widely used conventional indices of ventricular function and therefore provide a direct benchmark for the latent motion traits. In contrast, LVEDVi reflects chamber size rather than dynamic performance and was included as a secondary check to ensure that associations with the latent traits were not driven by static geometry. For the novelty comparison, motion loci passing the motion-domain threshold (*P* < 7.14 × 10^−9^) were compared with the cumulative union of loci reaching *P* < 5 × 10^−8^ in LVEF, LVEDVi or Ell global. A motion locus was classified as not detected by the prespecified conventional comparators when no overlapping comparator locus reached genome-wide significance.

### LD score regression (LDSC)

We used LD Score Regression (LDSC v1.0.1) for univariate and bivariate analyses to quantify SNP heritability, assess confounding and estimate genetic correlations among the 32 latent motion traits *z*_1_–*z*_32_ and three conventional measures (LVEF, Ell global, LVEDVi). For each trait, we started from GWAS summary statistics derived with REGENIE whole-genome regression, and harmonised them using the munge_sumstats.py script from the LDSC package, restricting to well-imputed HapMap3 variants, aligning alleles to the 1000 Genomes Phase 3 European reference panel, and excluding strand-ambiguous SNPs and variants with extreme allele frequency discrepancies. Harmonised summary statistics were processed with ldsc.py using the baselineLD v2.2 (GRCh38) annotation model to yield observed-scale SNP heritability, standard errors, and LDSC intercepts. Bivariate LDSC (ldsc.py –rg) was performed for all trait pairs using the same LD scores and regression weights to generate the 35 × 35 genetic correlation matrix. The resulting genetic-correlation matrix was then combined with Spearman’s rank phenotypic correlations *r*_*p*_ computed using latent scores (*z*_1_–*z*_32_) and rank-inverse-normal transformed conventional traits (LVEF, Ell global, LVEDVi). To facilitate comparison, hierarchical clustering was performed on the genetic correlation matrix using the distance metric *D* = (1 − *r*_*g*_)∕2. Agglomerative clustering with average linkage determined the dendrogram leaf order, which was applied to both *r*_*g*_ and *r*_*p*_ matrices. Data were visualised as a composite heatmap displaying *r*_*p*_ (lower triangle), *r*_*g*_ (upper triangle), and ℎ̂^2^ (diagonal).

### Stratified LD score regression (S-LDSC)

We used stratified LD score regression (S-LDSC; baselineLD v2.2, GRCh38) to partition SNP heritability across functional annotations. For each of the 32 latent traits and three conventional measures, summary statistics were harmonised and regressed on baselineLD v2.2 LD scores, and we extracted the rescaled regression coefficient *τ*^∗^ wherever available (or the raw coefficient *τ* when *τ*^∗^ was not reported) as the primary measure of the conditional per-SNP contribution of each annotation to heritability.

To facilitate biological interpretation, baseline tracks were mapped *a priori* into functional families, defined by genomic position and chromatin state: coding sequence; 5^′^ and 3^′^ UTRs; intronic and intron-flanking regions; promoters and transcription start sites (TSS); enhancers (distal and super-enhancer); histone marks (H3K27ac, H3K4me1, H3K4me3, H3K9ac); open chromatin (DNase I hypersensitive sites and digital genomic footprints); transcription factor binding sites; CTCF insulators; transcribed, Polycomb-repressed, and bivalent chromatin; and evolutionary conservation and ancient sequence-age metrics. For relevant features we modelled the 500 bp flanking windows (”flanking.500”) as distinct families. Baseline “nuisance” annotations (e.g., MAF bins, recombination rate, nucleotide diversity, CpG content, and baseL2_0), which capture background selection and LD structure, were retained in all models but not interpreted as biological categories.

Within each trait we first collapsed multiple baseline tracks assigned to the same functional family using inverse-variance weighted (IVW) meta-analysis to obtain a single family-level effect estimate and standard error per trait. We then performed a second-stage IVW meta-analysis across the 32 latent motion traits to quantify shared functional architecture, computing *Z*-scores as *μ̂*_*f*_ ∕SE(*μ73×0302;*_*f*_) for each family and deriving two-sided *P*-values under a normal approximation. Across-family false discovery rate was controlled using the Benjamini-Hochberg procedure, and we report these meta-analytic family-level coefficients as the primary summary of how latent-trait heritability is distributed across regulatory, coding and conserved genomic domains.

### Locus-to-gene mapping and functional annotation

Independent genetic loci were defined by linkage-disequilibrium (LD) clumping of GWAS summary statistics. For trait-wise GWAS we used--clump-p1 5e-8 --clump-p2 1e-2 --clump-r2 0.1 --clump-kb 250. For the pan-latent minimum-*P* scan, the primary threshold was tightened to 7.14 × 10^−9^ to reflect the study-wide cut-off. Lead variants and locus boundaries from clumping were propagated to downstream functional mapping. Effector genes were prioritised using FUMA (v1.8.1)^13^, integrating three evidence streams: (i) positional mapping based on genomic proximity and predicted functional consequence; (ii) expression quantitative trait loci (eQTLs) from GTEx v8 in heart–left ventricle (LV) and heart–atrial appendage (AA); and (iii) LV Hi-C chromatin interactions linking distal regulatory elements to promoters. FUMA was configured with a lead-SNP threshold of *P* ≤ 5× 10^−8^ (or *P* ≤ 7.14 × 10^−9^ for the pan-latent scan), a candidate-SNP threshold of *P* < 0.05, LD thresholds of *R*^2^ ≥ 0.6 (independent significant SNPs) and *R*^2^ ≥ 0.1 (lead SNPs), and the UK Biobank release-2b 10k Europeans reference panel.

Because FUMA operates on GRCh37/hg19 whereas discovery GWAS used GRCh38, a subset of rare or solitary imputed lead variants were manually harmonised and annotated using the OpenTargets Locus-to-Gene resource and LocusZoom. When neither resource provided a confident target, the nearest gene was recorded. Gene identities were normalised to Ensembl identifiers and duplicates aggregated. The union of FUMA-derived and harmonised gene assignments defined the latent-trait gene catalogue for downstream analyses.

### MAGMA gene-based and tissue-property analyses

MAGMA (v1.08)^15^ was used to obtain an orthogonal gene-level view that adjusts for gene length and local LD. SNP-to-gene assignment followed MAGMA defaults, and gene association statistics were computed with the SNP-wise mean model. When reporting gene-level discoveries we applied Bonferroni correction over the number of tested genes.

To assess whether association strength concentrated within particular tissues, we ran MAGMA’s gene-property analysis against GTEx v8 expression profiles for 54 tissues. For each tissue *t*, the model regressed gene-level Z scores on expression in *t*, conditioning on gene size and SNP density. Significance across tissues was controlled at a family-wise level of *α* = 0.05∕54 ≈ 9.3 × 10^−4^. The same procedure was applied to the pan-latent scan, LVEF, and Ell global to compare cardiac specificity among phenotypes.

MAGMA analysis adds three layers of value to the GWAS. First, it prioritises genes within multi-gene loci by harnessing aggregate signal while controlling for gene-level confounds. Second, it demonstrates that the genetic architecture of ventricular motion coheres into biologically interpretable pathways centred on sarcomere function, cardiac development and electrical conduction. Third, it localises the tissue context to the heart, thereby strengthening causal inference and complementing the left-ventricle-specific regulatory evidence used for variant-to-gene mapping.

### Transcriptome-wide association studies

Transcriptome-wide association studies (TWAS) were performed using the FUSION framework on the FarmGTEx TWAS-server^16^ with GTEx v8 expression prediction models for LV and AA. The server was accessed on 10 November 2025. TWAS provides an axis of evidence complementary to traditional GWAS locus discovery and MAGMA gene-based analysis. Whereas MAGMA aggregates SNP-level associations to the gene level while conditioning on gene size and local LD, TWAS tests whether *cis*-eQTL-driven variation in gene expression within specific cardiac tissues mediates phenotypic variation. As such, TWAS can elevate distal regulatory targets at multi-gene loci and de-emphasise large coding loci with limited *cis*-eQTL architecture. For the motion traits, we analysed the pan-latent minimum-*P* GWAS while carrying forward the multiple-trait correction to account for the effective number of independent motion components. Conventional traits were analysed in standard single-trait mode. Summary statistics were harmonised to the FUSION input format, including allele alignment and removal of strand-ambiguous variants. LD information was drawn from the GTEx v8 reference (European ancestry) or a matched European LD reference panel. For each trait-tissue combination, associations between genetically predicted gene expression and the phenotype were tested, and per-gene *Z* and *P* statistics were reported.

Primary TWAS significance was determined using a Bonferroni threshold within each trait–tissue combination:

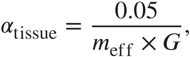

where *G* is the number of genes with valid prediction models in that tissue and *m*_eff_ = 7 for the pan-latent analysis (reflecting the effective number of independent motion latents) and *m*_eff_ = 1 for LVEF and Ell global. Benjamini–Hochberg (BH) FDR at *q* < 0.05 was also reported as a sensitivity analysis. Because LV and AA were pre-specified tissue contexts for the heart, we did not impose an additional across-tissue Bonferroni correction in the primary analysis.

### Cellular context of genetic associations

To resolve the cellular context of trait-associated genes, we leveraged a single-nucleus RNA-seq atlas of the adult human heart^70^. This dataset profiles over half a million nuclei from all four chambers and major vascular compartments, providing quantitative expression across the dominant myocardial, vascular, and stromal populations. Cell types comprised ventricular and atrial cardiomyocytes, fibroblast, myofibroblast, endothelial, endocardial, arterial smooth muscle, pericyte, macrophage, lymphocyte, adipocyte and Schwann cell. For downstream analyses, we obtained the absolute expression matrix, aggregated duplicate Ensembl IDs by their mean, and applied a natural log transformation, log(1 + *x*), to stabilise variance and mitigate the influence of highly expressed genes. The transformed matrix thus represents the per-cell-type expression intensity of each gene on a continuous, variance-stabilised scale suitable for statistical comparison across gene sets. This approach preserves absolute abundance information necessary for enrichment testing, while controlling for heteroscedasticity inherent to single-cell transcript counts. To examine how cell-type specificity evolved with increasing strength of genetic evidence, we implemented a sliding-threshold analysis of ranked gene lists from TWAS (LV and AA; ranked by − log_10_*P*_TWAS_), and proximity/functional mapping (ranked by − log_10_*P*_GWAS_). For a threshold *u*, let *S*_*u*_ denote genes with score ≥ *u*. For each cell type *c* we computed a standardised mean difference,

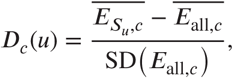

where *E* denotes log(1 + *x*) expression. Curves of *D*_*c*_(*u*) versus *u* were drawn on fixed *u* grids. Calculations were done with the background restricted to heart-expressed genes (mean absolute expression > 0.1), to avoid inflating contrasts by including unexpressed genes in the denominator.

### Summary-statistics fine-mapping

We performed summary-statistics fine-mapping across all 35 cardiac imaging traits, comprising 32 latent motion traits (*z*_1_–*z*_32_) and three conventional traits (*LVEF*, *LVEDVi*, and *Ell* global). Fine-mapping was carried out using SuSiE-RSS with locus-specific LD matrices computed from reference-based UK Biobank genotype data, using reference-allele-oriented LD to ensure consistency between effect estimates and correlation structure. SuSiE-RSS was run with *L* = 10, refine = TRUE, estimate_residual_variance = FALSE, max_iter = 500, and tol = 10^−3^. For each retained trait–locus pair, we extracted posterior inclusion probabilities (PIPs) for all variants and defined 95% credible sets. Downstream analyses were restricted to loci with *lead* PIP ≥ 0.1, while retaining the full 95% credible set within each retained locus. Thus, locus selection was based on minimum fine-mapping resolution, whereas variant-level overlap analyses preserved the full posterior support structure within each selected locus.

For downstream summarisation, loci were classified according to their sharing pattern across trait domains. A locus was labelled *conventional-only* if it was fine-mapped in at least one conventional trait and in no motion latent, *motion-only* if it was fine-mapped in at least one motion latent and in no conventional trait, and *shared* if it was fine-mapped in both domains.

### Overlapping fine-mapped variants with the foetal heart regulome

To map fine-mapped variants to regulatory elements, candidate target genes, and cardiac cell states, we intersected credible-set variants with the single-cell enhancer-to-gene (scE2G) atlas of the developing human heart^71,72^. The scE2G framework integrates activity-by-contact (ABC) score^73^, chromatin accessibility, and single-cell co-variation in accessibility and expression to predict enhancer–gene interactions. We defined high-confidence regulatory links using the validated scE2G score threshold of 0.171 and mapped variants to the 90 available foetal-heart cell states.

For primary analyses, we restricted scE2G overlaps to enhancer elements (class ≠ promoter) to maximise cell-state specificity. Analyses including all regulatory elements (enhancers and promoters) were retained as sensitivity analyses and for gene-linking support. Variant overlaps were defined at single-base resolution in GRCh38. To avoid inflating counts when the same regulatory element was linked to multiple genes, scE2G intervals were first deduplicated to unique element coordinates within each cell state for overlap calculations; gene-level links were then restored after overlap assignment.

For higher-level summaries, each of the 90 foetal-heart states was assigned to one of 13 broad cell-programme groups: atrial cardiomyocytes, ventricular cardiomyocytes, conduction cells, transitional cells, endocardial cells, endothelial cells, fibroblasts, mural cells, epicardial cells, autonomic neurons, neural crest cells, lymphoid cells, and myeloid cells. The full 90-state atlas remained the primary resolution for biological interpretation; the 13-group collapse was used only for summarisation and figure panels.

### PIP-weighted enrichment analysis

For a given trait, locus, and annotation, overlap was quantified using PIP-weighted credible-set support. The observed overlap for locus *ℓ* was defined as

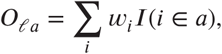

where *w*_*i*_ is the PIP of variant *i*, and *I*(*i* ∈ *a*) indicates whether variant *i* overlaps annotation *a*. The expected overlap under a within-locus null was defined as

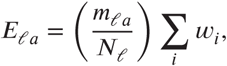

where *m*_*ℓa*_ is the number of variants in locus *ℓ* that overlap annotation *a*, and *N*_*ℓ*_ is the total number of variants in the locus. This formulation controls for locus-specific annotation density while preserving posterior support from fine-mapping. Linkage disequilibrium is handled upstream by SuSiE-RSS rather than by the overlap statistic itself.

For each trait and annotation, effect size was reported as

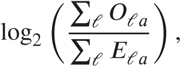

and statistical significance was assessed using pooled within-locus residual *Z*-statistics. Resulting *P*-values were adjusted across tests using the Benjamini–Hochberg false discovery rate (FDR) procedure. Trait-resolved enrichment analyses were performed both at full 90-state resolution and after collapse to the 13 broad groups.

### Combined motion summaries for visualisation

To facilitate visualisation across the 32 latent motion traits, we generated combined motion summaries for figure panels. These summaries were used for visualisation and domain-level comparison only and did not replace the primary per-trait enrichment analyses. In grouped panels derived from the 90-state analysis, member-state *P*-values were combined within each of the 13 broad cell-programme groups using the aggregated Cauchy association test (ACAT), and the resulting group-level *P*-values were FDR-corrected across groups. Separately, Figure 5b shows the within-domain distribution of cumulative PIP-weighted regulatory support, normalised to 100%, whereas Figure 5c,d summarise positive meta-*Z* contributions across motion traits and their difference from conventional-trait support. These cumulative displays are descriptive and were not used for statistical inference; formal cell-state inference is provided by the FDR-corrected 90-state and 13-group scE2G enrichment tests. FUMA, MAGMA and TWAS were used as first-pass locus, pathway and transcriptomic annotation layers. The final candidate-gene set was then derived from the mutually exclusive fine-mapped hierarchy below, prioritising cardiac scE2G links for noncoding regulatory variants, coding or splice consequences for direct molecular effects, Open Targets L2G for unresolved noncoding variants, and GTEx/ENCODE evidence as secondary pan-tissue support.

### Hierarchical variant-to-gene prioritisation

We applied a five-tier hierarchical evidence framework to all 188,631 credible-set variants passing the lead-PIP locus filter, assigning each variant to the highest-supported tier in a mutually exclusive manner.

#### Tier 1 — heart scE2G regulatory overlap

Variants overlapping scE2G enhancer elements (score ≥ 0.171, class ≠ promoter) were assigned their target-gene links, cell types, broad cell-programme groups, and quantitative scE2G metrics. When multiple genes were linked to the same variant, the priority gene was defined as the gene with the highest scE2G score in the supplied table. This tier resolved 32,835 variants (17.4%) to 486 unique priority genes across 81 loci.

#### Tier 2 — coding and splice-altering variants

Variants not resolved by Tier 1 were assessed for direct functional consequence using Ensembl VEP v113^74^ against the GRCh38 reference. We defined rescued coding/splice variants with HIGH or MODERATE VEP impact (comprising missense, splice, stop-loss/gain, frameshift, in-frame indel, start-loss, and related consequences). This resolved 867 variants (0.5%) to 277 unique priority genes across 69 loci. Where a Tier 2 variant already existed in Open Targets Variant dataset, or also carried GTEx eQTL or ENCODE cCRE support, those annotations were retained as secondary evidence rather than replacing the primary coding/splice label.

#### Tier 3 — Open Targets Locus-to-Gene (L2G)

Variants not resolved by Tiers 1 or 2 were assigned a priority gene using the Open Targets L2G machine-learning model^75^, which integrates colocalisation, distance, molecular QTL, and chromatin interaction evidence to predict the most likely causal gene at each GWAS locus. This tier resolved 7,271 variants (3.9%) to 491 unique priority genes across 83 loci. Together, Tiers 1–3 resolved 40,973 variants (21.7%) to 643 unique priority genes across all 87 loci.

#### Tier 4 — pan-tissue regulatory cross-reference

Variants unresolved after Tiers 1–3 were cross-referenced against two pan-tissue resources: GTEx v11 significant eQTLs across 50 tissues^76^ and ENCODE SCREEN v4 candidate cis-regulatory elements (cCREs)^77^. The GTEx cross-reference recorded all significant variant–gene–tissue associations and summarised, for each variant, the linked genes, number of tissues, best eQTL *P*-value, and strongest effect size. The ENCODE cross-reference recorded cCRE type (distal enhancer-like, proximal enhancer-like, promoter-like, or CTCF-bound), accessions, and the number of overlapping elements. Variants with evidence from at least one of these sources were classified as Tier 4 and stratified into five mutually exclusive sub-categories by evidence source: GTEx eQTL only (31,231; 40.0%), ENCODE cCRE only (19,002; 24.3%), GTEx and cardiac eQTL (14,893; 19.1%), GTEx and ENCODE (8,906; 11.4%), and GTEx, ENCODE, and cardiac eQTL (4,057; 5.2%), yielding 78,089 Tier 4 variants in total (41.4%). Pan-tissue eQTL and cCRE information was retained as secondary evidence for interpretation of the residual unresolved set; Tier 4 gene assignments were not used as primary causal nominations.

#### Tier 5 — unannotated

The 69,569 variants (36.9%) lacking evidence from any of the above layers were classified as Tier 5. These variants remain compatible with developmental-stage-specific regulatory activity, transient or low-abundance cell populations under-sampled in current atlases, and regulatory mechanisms operating in cell contexts not yet resolved at single-cell resolution. The Tier 5 fraction therefore reflects the boundary of current functional annotation resources rather than an absence of biological function, and is expected to diminish as cardiac cell-state atlases expand across developmental stages and rare populations.

### Gene-level prioritisation and exemplar variant extraction

For gene-level prioritisation, genes were ranked first by evidence tier and then by the maximum PIP of any supporting variant. Where scE2G evidence was present, the scE2G-linked target gene was treated as the highest-confidence cardiac-context assignment; coding/splice genes (Tier 2) and L2G genes (Tier 3) served as rescue layers when scE2G support was absent.

To prioritise high-confidence exemplars for detailed narrative discussion, we extracted variants with PIP > 0.8, considered near-certain single-variant causal nominations. In addition, several loci contained multiple independently supported credible-set variants within the same tier, each at individually moderate PIP but collectively accounting for a large fraction of the posterior mass within a single causal signal (cumulative PIP across the credible set > 0.8). Under the SuSiE-RSS framework, such distributed posterior support arises when linkage disequilibrium prevents resolution to a single variant but the aggregate evidence for a causal effect at the locus remains strong^78,79^. Exemplar variants at these loci were therefore retained using a cumulative-PIP criterion to avoid penalising loci where LD limits variant-level resolution despite coherent locus-level evidence. We additionally retained a small number of variants with lower PIP but strong orthogonal support (Tier 2 coding/splice, or high-confidence Tier 3 L2G assignments) as biologically informative exemplars, in recognition that posterior inclusion probability quantifies statistical fine-mapping confidence rather than biological relevance.

Variants without scE2G overlap were retained in the exemplar table, as these may act through protein-altering or otherwise non-regulatory mechanisms not captured by the foetal enhancer map. Loci were classified as Shared if the genomic window contained fine-mapped credible-set variants (lead PIP > 0.1, 95% credible set) for both conventional and motion trait domains. This classification is based on locus-level co-occurrence of fine-mapped signals and does not imply that the same causal variant underlies both trait domains.

### Cell-type enrichment of prioritised gene sets in non-failing and dilated cardiomyopathy hearts

To examine cell-type specificity and disease-state behaviour of the prioritised gene sets in an independent adult dataset, we analysed a published snRNA-seq atlas of the human heart^18^, restricted to left ventricular samples (including interventricular septum) from non-failing donors (Normal, *n* = 18) and dilated cardiomyopathy patients (DCM, *n* = 52). The resulting dataset comprised 612,457 nuclei across ten annotated cell populations (ventricular cardiomyocytes, fibroblasts, mural, endothelial, endocardial, adipocyte, neuronal, myeloid, lymphoid, and mast cells). Three gene sets were tested, defined by the hierarchical variant-to-gene framework: Conventional (loci fine-mapped only in LVEF, Ell global, or LVEDVi), Shared (loci fine-mapped in both domains), and Motion (loci fine-mapped only in the 32 latent motion traits). Gene identifiers were harmonised against the h5ad gene model using MyGene.info and HGNC aliases; genes absent from GRCh38.98 were excluded, retaining 368 Conventional, 83 Shared, and 75 Motion entries.

Cell-type preference in Normal hearts was quantified as the detection rate ratio: the mean percentage of nuclei expressing a gene set within a cell type, divided by the mean across all ten cell types (Figure 9d; values > 1 indicate preferential detection).

Per-donor gene-set Z-scores (Figure 9f) were computed as *Z* = (*x̄*_set_ − *μ*_null_)∕*σ*_null_, where *x̄*_set_ is the mean normalised expression of set genes for a given donor and cell type, and *μ*_null_, *σ*_null_ are the mean and standard deviation of 1,000 random gene sets of matched size sampled from the same donor–cell-type group. This matched-size background controls for set size and expression level. Differences between Normal and DCM Z-scores were tested by two-sided Wilcoxon rank-sum tests across the 30 gene-set × cell-type combinations, with Benjamini–Hochberg correction.

Pseudobulk differential expression analysis was performed with DESeq2 (v1.44) using mean counts per cell scaled to a common library size of 10,000 per donor × cell-type combination (≥ 10 nuclei), which decouples DESeq2 size factors from cell-recovery differences between conditions (Figure 9c). The design formula was ∼ sex + condition, with apeglm log-fold-change shrinkage. Genes were called differentially expressed at Benjamini–Hochberg-adjusted *p* < 0.05 and log_2_ FC > 0.25. Enrichment of prioritised gene sets among cell-type DEGs was assessed by Fisher’s exact test against all tested genes in each cell type (Figure 9e), with Benjamini–Hochberg correction across the 30 tests; dashes indicate combinations with no overlap.

UMAP feature plots (Figure 9g) show log-normalised expression of highly detected Motion-set genes on a 100,000-nucleus random subsample of Normal donors, using pre-computed UMAP coordinates from the h5ad object.

### External molecular and disease validation

External validation was designed to test whether the fine-mapped LV imaging loci were corroborated by independent molecular and cardiovascular-disease evidence. The strict cross-resource validation manifest contained 560 credible-set variants from 87 loci, comprising 19 motion-only, 11 shared and 57 conventional-only loci. Trait-domain classifications were inherited from the UK Biobank SuSiE-RSS analysis, and UK Biobank-derived posterior inclusion probabilities were retained as variant weights. All variants and loci were represented on GRCh38.

#### GPMap colocalisation

Per-trait 95% credible sets from 23 motion latent traits and the three conventional traits with available fine-mapped signals were used as the LV study-extraction layer for GPMap. One extraction was defined for each trait–locus–credible-set combination, retaining all credible-set variants, alleles, association statistics and PIPs. Variant-anchored GPMap queries were performed without preselection of cardiovascular, developmental, molecular-QTL or other external phenotype categories. External colocalisation support was defined as posterior probability *H*_4_ ≥ 0.8. The complete GPMap credible-set universe comprised 109 unique physical loci (24 motion-only, 13 shared and 72 conventional-only); the matched strict 87-locus subset was used for integrated cross-resource displays.

Developmental and congenital gene enrichment in GPMap was tested at the unique-locus level. Locus-linked genes were intersected with a prespecified developmental and congenital-heart-disease gene dictionary, and the proportion of motion-only and conventional-only loci containing at least one category gene was compared by Fisher’s exact test, with Benjamini–Hochberg correction across gene categories. For regulatory localisation, scE2G-linked PIP was summed within each trait–locus credible set and partitioned between cardiomyocyte and non-myocyte cardiac programmes. Among credible sets with non-zero scE2G-regulatory PIP, the non-myocyte regulatory-PIP share was compared between motion and conventional traits using Mann–Whitney *U* tests; FDR correction was applied across localisation metrics, and bootstrap resampling was used to derive 95% confidence intervals for mean motion-minus-conventional differences.

#### FinnGen disease-module validation

The strict 87-locus manifest was tested against FinnGen R13 sum-mary statistics for 13 prespecified cardiac endpoints. Myocardial-remodelling endpoints comprised heart fail-ure, dilated cardiomyopathy and hypertrophic cardiomyopathy (I9_HEARTFAIL, I9_DILATED_CARDIOMYOPATHY, I9_HYPERTROCARDMYOP); electrical/conduction endpoints comprised atrial fibrillation, conduction disorders and atrioventricular block (I9_AF, I9_CONDUCTIO, I9_AVBLOCK); valvular/endocardial disease was represented by I9_VHD; and developmental/congenital structural endpoints comprised congenital malformations of the circula-tory system, atrial septal defect, ventricular septal defect, left-ventricular outflow-tract obstruction, tetralogy of Fallot and bicuspid aortic valve (Q17_CONGEN_MALFO_CIRCULATO_SYSTEM, Q17_ASD, Q17_VENTRICULAR_SEPTAL_DEFECT, Q17_LVOTO_BROAD, Q17_TETRALOGY_OF_FALLOT, Q17_BICUSP_AORTIC_VALVE).

Manifest variants were matched to FinnGen R13 records by GRCh38 position and reference and alternate alleles. Allele orientation was harmonised to a common reference/alternate coding, and effect directions were sign-reversed wherever the effect and non-effect alleles were swapped relative to the manifest. Where FinnGen reported a probability-scale *P* of exactly zero (numerical underflow), *P* was recovered from the − log_10_ *P* field as 10^− min(− log^10 ^*P*, 300)^ and floored at 10^−300^. Within each locus–endpoint combination, matched variant *P*-values were combined using PIP-weighted ACAT, with UK Biobank PIPs as relative weights, as the primary statistic; a parallel statistic restricted to lead and high-confidence variants was computed as a sensitivity analysis. Endpoint-level evidence was combined by ACAT into developmental/congenital, valvular/endocardial, electrical/conduction and myocardial-remodelling modules. The prespecified primary disease axis combined the developmental, valvular and conduction modules. Benjamini–Hochberg correction was applied separately across locus–module and locus–axis tests.

#### All of Us EHR-genetic validation

The same 87-locus manifest was evaluated in the All of Us All-by-All v8 ACAF resource across 49 prespecified cardiovascular EHR phenotypes. Phenotypes were assigned to developmental/congenital structural, valvular/endocardial, electrical/conduction, myocardial-remodelling, ischaemic/MACE, loading/cardiometabolic and stromal/vascular/ECM modules. Variant matching, allele harmonisation and within-locus PIP-weighted ACAT aggregation followed the FinnGen procedure. Endpoint statistics were then combined into disease modules and into the prespecified developmental/valvular/conduction axis, with Benjamini–Hochberg correction applied separately at the locus–module and locus–axis levels.

The European-ancestry analysis was prespecified as the primary ancestry-matched All of Us analysis. The All-by-All cross-ancestry META analysis was treated as an aggregate sensitivity analysis, and AFR, AMR, EAS and SAS strata were evaluated separately to assess ancestry-specific transferability and phenotype coverage. In the META analysis, *P*-values were reported on the natural-log scale and were exponentiated to the probability scale before aggregation, then subjected to variant-, locus-, module- and axis-level range checks. An ancestry-specific axis statistic was calculated only when at least one developmental/congenital, valvular/endocardial or electrical/conduction endpoint was available. SAS contained no eligible primary-axis endpoint and was therefore classified as not evaluable rather than as having no supported loci.

Across FinnGen and All of Us, FDR support was defined as *q* < 0.05, and nominal support as *P* < 0.05 without FDR significance. Locus-class proportions were reported with 95% Wilson confidence intervals; class and annotation contrasts were tested with Fisher’s exact test, signal-strength contrasts with the Mann–Whitney *U* test, and the lead-versus-PIP FDR contrast with the exact McNemar test. These analyses test external molecular and disease-context corroboration of the fine-mapped LV imaging loci; FinnGen and All of Us do not contain comparable 4D CMR motion phenotypes and were therefore not interpreted as direct replication of the imaging traits.

### Tree-based unsupervised taxonomy of motion

To enable interpretable visualisation and unsupervised stratification of cardiac motion phenotypes, we applied a graph-based dimensionality reduction method to the population-scale latent space. Specifically, we used Discriminative Dimensionality Reduction via Learning a Tree (DDRTree)^26^, a manifold learning algorithm that jointly learns (i) a low-dimensional embedding, (ii) a tree-structured graph capturing the global geometry of the data and (iii) a soft clustering structure that preserves local neighbourhood coherence. DDRTree projects the high-dimensional motion representations *Z* ∈ ℝ^*d*^ extracted from the *Cardio4D-VAE* into a two-dimensional space while enforcing a minimum spanning tree constraint, preserving both local neighborhood continuity and global branching structure.

The resulting tree topology provides a biologically interpretable representation of population-level diversity in left-ventricular motion and supports downstream analyses such as clustering, genetic association, and trajectory modeling. The algorithm operates by jointly minimising a reconstruction loss, a graph smoothness term, and a soft clustering objective, enabling both local geometry preservation and global structure recovery.

Given latent features 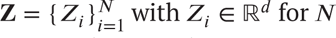 for *N* individuals, DDRTree learns a low-dimensional embedding 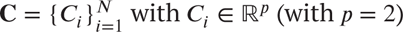, by minimising the following objective:

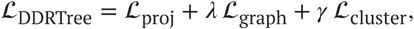

where 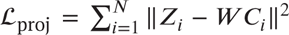 ensures accurate reconstruction of the original latent features from their low-dimensional projections via a linear mapping *W* ∈ ℝ^*d*×*p*^. To incorporate the discriminative information, the method introduces another set of latent points 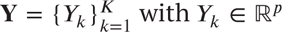, as the centers of 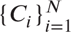. The graph regularisation term, 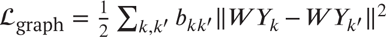, enforces smoothness over a minimum spanning tree defined on cluster centers {*Y*_*k*_}, with adjacency matrix *B*. The clustering term, 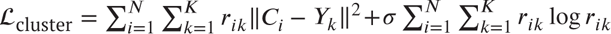, encourages soft association of each embedding *C*_*i*_ to the cluster centers, regularised by an entropy penalty. The optimisation is subject to the constraints 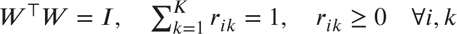; where *W* ∈ ℝ^*d*×*p*^ is an orthonormal projection matrix, **Y** = {*Y*_*k*_} ⊂ ℝ^*p*^ are cluster centers in the embedding space, *B* ∈ ℝ^*K*×*K*^ encodes the tree structure over the clusters, and *R* ∈ ℝ^*N*×*K*^ is a soft assignment matrix. The hyperparameters *λ*, *γ*, and *σ* control the relative weights of the graph regularisation, clustering fidelity, and entropy penalty, respectively.

This optimisation enables DDRTree to produce a two-dimensional manifold embedding (*C*) of the 32-dimansional latent cardiac motion features (*Z*), in which individuals are organized along continuous phenotypic trajectories and branching structures. We defined the root node of the tree model as the subject located closest to the global mean in the low-dimensional embedding space, serving as the origin point for all subsequent branches. For each individual, a continuous pseudotime value was calculated to represent their relative progression along a given branch from the root to its terminal node (Supplementary Figure 17a). The minimum spanning tree (MST) was constructed using the root as the origin, with each branch corresponding to the shortest path from the root to a leaf node, representing a terminal subject within that trajectory. The closest points to the MST nodes were then identified to build the lineage graph. Disconnected subgraphs were iteratively connected to allow for a single joined component. This process was repeated over multiple iterations, testing different internal parameters of algorithm, in order to optimise the stability of the final tree structure. Differential analyses were performed to identify significant changes in the 32-dimensional latent cardiac motion features between the start and end points of each lineage, using the R package *TradeSeq* v 4.5.2 (Supplementary Figure 11).

#### Experimental settings

The optimisation of DDRTree model was implemented using the R package *Monocle* v4.5.2^80^, which incorporates the DDRTree algorithm for manifold learning and pseudotemporal trajectory inference. Hyperparameters were set empirically to balance projection fidelity, graph regularisation, and clustering robustness. The parameters *λ*, *γ*, and *σ* were tuned to ensure stability of the embedded topology across iterations. Specifically, *λ* was set to 5 based on model stability analysis, while *γ*, and *σ* were maintained at their default values as defined in the DDRTree implementation.

### Stability analysis

We tested the stability of the tree by evaluating the preservation of nearest neighbours (NN) in the model, with varying *γ* values (2, 5, 10). The parameter *γ* influences the balance between fitting the data and maintaining the simplicity of the partitioned structure. For each *γ* value, 500 replicates were generated by randomly sampling 90% of the data and dimensionality reduction was performed. At each set level of *γ*, we tested the median preservation (%) of 50 nearest neighbours in the tree’s structure across the replicates. The chosen value of *γ* for our final model was set at 5, as this was felt to reflect the optimum between over-fitting the model and maintaining an acceptable (%) median NN at 75% (Supplementary Figure 17b).

## Author contributions

S.K. and D.P.O. conceived the project and designed the experiments. N.V. led the genetic component of the study, including genome-wide association analyses, fine-mapping, single-cell multiomic integration, and functional annotation. Y.Q. and M.N. collaborated on the single cell data, and K.A.M. performed the rare variant classification analysis. L.C., M.C., K.R., P.G., and C.B. performed data analysis and provided guidance on experiments. J.Z., Q.M., and W.B. performed the image analysis. S.K., N.V. and D.P.O. prepared the manuscript with input from all authors.

## Data availability

All raw and derived data in this study is available from UK Biobank (http://www.ukbiobank.ac.uk/). GWAS summary level data will be publicly available through the GWAS catalogue.

## Code availability

The code used in this analysis is available from GitHub (https://github.com/ImperialCollegeLondon/cardio4d) as well as the pipeline for image analysis (https://github.com/ImperialCollegeLondon/DeepMesh).

## Supplementary materials

Supplementary Methods

Figures S1 to S15

Tables S1 to S10

Movies S1 to S4

Data S1 to S6

## References

1. Buckberg G, Hoffman JIE, Mahajan A, Saleh S, and Coghlan C. Cardiac Mechanics Revisited: The Relationship of Cardiac Architecture to Ventricular Function. Circulation. 2008;118:2571–87. DOI: 10.1161/CIRCULATIONAHA.107.754424.

2. Cikes M and Solomon SD. Beyond ejection fraction: an integrative approach for assessment of cardiac structure and function in heart failure. Eur Heart J. 2016;37:1642–50. DOI: 10.1093/eurheartj/ehv510.

3. Meng Q, Qin C, Bai W, Liu T, de Marvao A, O’Regan DP, and Rueckert D. MulViMotion: Shape-Aware 3D Myocardial Motion Tracking From Multi-View Cardiac MRI. IEEE Trans Med Imaging. 2022;41:1961–74. DOI: 10.1109/TMI.2022.3154599.

4. Meng Q, Bai W, O’Regan DP, and Rueckert D. DeepMesh: Mesh-Based Cardiac Motion Tracking Using Deep Learning. IEEE Trans Med Imaging. 2024;43:1489–500. DOI: 10.1109/TMI.2023.3340118.

5. Biffi C, Oktay O, Tarroni G, Bai WJ, De Marvao A, Doumou G, Rajchl M, Bedair R, Prasad S, Cook S, et al. Learning Interpretable Anatomical Features Through Deep Generative Models: Application to Cardiac Remodeling. Medical Image Computing and Computer Assisted Intervention – MICCAI 2018. 2018;11071:464–71. DOI: 10.1007/978-3-030-00934-2_52.

6. Bonazzola R, Ferrante E, Ravikumar N, Xia Y, Keavney B, Plein S, Syeda-Mahmood T, and Frangi AF. Unsupervised ensemble-based phenotyping enhances discoverability of genes related to left-ventricular morphology. Nat Mach Intell. 2024;6:291–306. DOI: 10.1038/s42256-024-00801-1.

7. Bello GA, Dawes TJW, Duan J, Biffi C, de Marvao A, Howard L, Gibbs JSR, Wilkins MR, Cook SA, Rueckert D, et al. Deep learning cardiac motion analysis for human survival prediction. Nat Mach Intell. 2019;1:95–104. DOI: 10.1038/s42256-019-0019-2.

8. Qiao M, McGurk KA, Wang S, Matthews PM, O’Regan DP, and Bai W. A personalized time-resolved 3D mesh generative model for unveiling normal heart dynamics. Nat Mach Intell. 2025;7:800–11. DOI: 10.1038/s42256-025-01035-5.

9. Bycroft C, Freeman C, Petkova D, Band G, Elliott LT, Sharp K, Motyer A, Vukcevic D, Delaneau O, O’Connell J, et al. The UK Biobank resource with deep phenotyping and genomic data. Nature. 2018;562:203–9. DOI: 10.1038/s41586-018-0579-z.

10. Bai W, Sinclair M, Tarroni G, Oktay O, Rajchl M, Vaillant G, Lee AM, Aung N, Lukaschuk E, Sanghvi MM, et al. Automated cardiovascular magnetic resonance image analysis with fully convolutional networks. J Cardiovasc Magn Reson. 2018;20:65. DOI: 10.1186/s12968-018-0471-x.

11. Thanaj M, Mielke J, McGurk KA, Bai W, Savioli N, de Marvao A, Meyer HV, Zeng L, Sohler F, Lumbers RT, et al. Genetic and environmental determinants of diastolic heart function. Nat Cardiovasc Res. 2022;1:361–71. DOI: 10.1038/s44161-022-00048-2.

12. Li J and Ji L. Adjusting multiple testing in multilocus analyses using the eigenvalues of a correlation matrix. Heredity (Edinb). 2005;95:221–7. DOI: 10.1038/sj.hdy.6800717.

13. Watanabe K, Taskesen E, van Bochoven A, and Posthuma D. Functional mapping and annotation of genetic associations with FUMA. Nat Commun. 2017;8:1826. DOI: 10.1038/s41467-017-01261-5.

14. Pruim RJ, Welch RP, Sanna S, Teslovich TM, Chines PS, Gliedt TP, Boehnke M, Abecasis GR, and Willer CJ. LocusZoom: regional visualization of genome-wide association scan results. Bioinformatics. 2010;26:2336–7. DOI: 10.1093/bioinformatics/btq419.

15. De Leeuw CA, Mooij JM, Heskes T, and Posthuma D. MAGMA: generalized gene-set analysis of GWAS data. PLoS Comput Biol. 2015;11:e1004219. DOI: 10.1371/journal.pcbi.1004219.

16. Zhang Z, Chen Z, Teng J, Liu S, Lin Q, Wu J, Gao Y, Bai Z, Li B, Liu G, et al. FarmGTEx TWAS-server: An Interactive Web Server for Customized TWAS Analysis. Genomics Proteomics Bioinformatics. 2025;23. DOI: 10.1093/gpbjnl/qzaf006.

17. Huffman JE, Gaziano L, Al Sayed ZR, Judy RL, Raffield LM, Biddinger KJ, Charest B, Chopra A, Gagnon D, Guo X, et al. An African ancestry-specific nonsense variant in CD36 is associated with a higher risk of dilated cardiomyopathy. Nat Genet. 2025;57:2682–90. DOI: 10.1038/s41588-025-02372-2.

18. Reichart D, Lindberg EL, Maatz H, Miranda AMA, Viveiros A, Shvetsov N, Gartner A, Nadelmann ER, Lee M, Kanemaru K, et al. Pathogenic variants damage cell composition and single cell transcription in cardiomyopathies. Science. 2022;377:eabo1984. DOI: 10.1126/science.abo1984.

19. Pirruccello JP, Bick A, Chaffin M, Aragam KG, Choi SH, Lubitz SA, Ho CY, Ng K, Philippakis A, Ellinor PT, et al. Titin Truncating Variants in Adults Without Known Congestive Heart Failure. J Am Coll Cardiol. 2020;75:1239–41. DOI: 10.1016/j.jacc.2020.01.013.

20. De Marvao A, McGurk KA, Zheng SL, Thanaj M, Bai W, Duan J, Biffi C, Mazzarotto F, Statton B, Dawes TJW, et al. Phenotypic Expression and Outcomes in Individuals With Rare Genetic Variants of Hypertrophic Cardiomyopathy. J Am Coll Cardiol. 2021;78:1097–110. DOI: 10.1016/j.jacc.2021.07.017.

21. Visseren FLJ, Mach F, Smulders YM, Carballo D, Koskinas KC, Bäck M, Benetos A, Biffi A, Boavida JM, Capodanno D, et al. 2021 ESC Guidelines on Cardiovascular Disease Prevention in Clinical Practice. Eur Heart J. 2021;42:3227–337. DOI: 10.1093/eurheartj/ehab484.

22. Lelieveld J, Pozzer A, Pöschl U, Fnais M, Haines A, and Münzel T. Loss of Life Expectancy from Air Pollution Compared to Other Risk Factors: A Worldwide Perspective. Cardiovasc Res. 2020;116:1910–7. DOI: 10.1093/cvr/cvaa025.

23. Czernin J, Sun K, Brunken R, Böttcher M, Phelps M, and Schelbert H. Effect of Acute and Long-Term Smoking on Myocardial Blood Flow and Flow Reserve. Circulation. 1995;91:2891–7. DOI: 10.1161/01.cir.91.12.2891.

24. Andersen TO, Sejling C, Jensen AK, Dissing AS, Severinsen ER, Drews HJ, Sorensen TIA, Varga TV, and Rod NH. Self-reported and tracked nighttime smartphone use and their association with overweight and cardiometabolic risk markers. Sci Rep. 2024;14:4861. DOI: 10.1038/s41598-024-55349-2.

25. Aung N, Sanghvi MM, Zemrak F, Lee AM, Cooper JA, Paiva JM, Thomson RJ, Fung K, Khanji MY, Lukaschuk E, et al. Association Between Ambient Air Pollution and Cardiac Morpho-Functional Phenotypes: Insights From the UK Biobank Population Imaging Study. Circulation. 2018;138:2175–86. DOI: 10.1161/CIRCULATIONAHA.118.034856.

26. Mao Q, Wang L, Goodison S, and Sun Y. Dimensionality reduction via graph structure learning. In: Proceedings of the 21th ACM SIGKDD international conference on knowledge discovery and data mining. 2015:765–74.

27. Nair ATN, Wesolowska-Andersen A, Brorsson C, Rajendrakumar AL, Hapca S, Gan S, Dawed AY, Donnelly LA, McCrimmon R, Doney ASF, et al. Heterogeneity in phenotype, disease progression and drug response in type 2 diabetes. Nat Med. 2022;28:982–8. DOI: 10.1038/s41591-022-01790-7.

28. Curran L, de Marvao A, Inglese P, McGurk KA, Schiratti PR, Clement A, Zheng SL, Li S, Pua CJ, Shah M, et al. Genotype-Phenotype Taxonomy of Hypertrophic Cardiomyopathy. Circ Genom Precis Med. 2023;16:e004200. DOI: 10.1161/circgen.123.004200.

29. Burns R, Young WJ, Aung N, Lopes LR, Elliott PM, Syrris P, Barriales-Villa R, Sohrabi C, Petersen SE, Ramirez J, et al. Genetic basis of right and left ventricular heart shape. Nat Commun. 2024;15:9437. DOI: 10.1038/s41467-024-53594-7.

30. Lu C, McGurk KA, Zheng SL, de Marvao A, Inglese P, Bai W, Ware JS, and O’Regan DP. New genetic loci implicated in cardiac morphology and function using three-dimensional population phenotyping. Circ Genom Precis Med. 2025;18:e005116.

31. Fathima I, Mahin A, Priyanka P, Naurah Vattoth N, Nishana A, Perunelly Gopalakrishnan A, Soman S, and Raju R. Cellular phospho-signaling map of the enigmatic serine/threonine kinase MAST2. Biochem Biophys Rep. 2025;44:102277. DOI: 10.1016/j.bbrep.2025.102277.

32. Liu W, Cai X, Duan S, Shen J, Wu J, Zhou Z, Yu K, He C, and Wang Y. E3 ubiquitin ligase Smurf1 promotes cardiomyocyte pyroptosis by mediating ubiquitin-dependent degradation of TRIB2 in a rat model of heart failure. Int Rev Immunol. 2025;44:165–79. DOI: 10.1080/08830185.2024.2434058.

33. Lv J, Chen Q, Wang J, Guo N, Fang Y, Guo Q, Li J, Ma X, Zhan H, Chen W, et al. Downregulation of MLF1 safeguards cardiomyocytes against senescence-associated chromatin opening. Nucleic Acids Res. 2025;53. DOI: 10.1093/nar/gkae1176.

34. Ahmad B, Dumbuya JS, Li W, Tang JX, Chen X, and Lu J. Evaluation of GFM1 mutations pathogenicity through in silico tools, RNA sequencing and mitophagy pahtway in GFM1 knockout cells. Int J Biol Macromol. 2025;304:140970. DOI: 10.1016/j.ijbiomac.2025.140970.

35. Hubmacher D and Apte SS. ADAMTS proteins as modulators of microfibril formation and function. Matrix Biol. 2015;47:34–43. DOI: 10.1016/j.matbio.2015.05.004.

36. Tang Y, Aryal S, Geng X, Zhou X, Fast VG, Zhang J, Lu R, and Zhou Y. TBX20 Improves Contractility and Mitochondrial Function During Direct Human Cardiac Reprogramming. Circulation. 2022;146:1518–36. DOI: 10.1161/CIRCULATIONAHA.122.059713.

37. De Sena-Tomas C, Aleman AG, Ford C, Varshney A, Yao D, Harrington JK, Saude L, Ramialison M, and Targoff KL. Activation of Nkx2.5 transcriptional program is required for adult myocardial repair. Nat Commun. 2022;13:2970. DOI: 10.1038/s41467-022-30468-4.

38. Yamak A, Latinkic BV, Dali R, Temsah R, and Nemer M. Cyclin D2 is a GATA4 cofactor in cardiogenesis. Proc Natl Acad Sci U S A. 2014;111:1415–20. DOI: 10.1073/pnas.1312993111.

39. Dickinson ME, Flenniken AM, Ji X, Teboul L, Wong MD, White JK, Meehan TF, Weninger WJ, Westerberg H, Adissu H, et al. High-throughput discovery of novel developmental phenotypes. Nature. 2016;537:508–14. DOI: 10.1038/nature19356.

40. Yang L, Gao X, Luo H, Huang Q, Su D, Tan X, and Lu C. TCF21 rs12190287 Polymorphisms Are Associated with Ventricular Septal Defects in a Chinese Population. Genet Test Mol Biomarkers. 2017;21:312–5. DOI: 10.1089/gtmb.2016.0324.

41. Schafer S, de Marvao A, Adami E, Fiedler LR, Ng B, Khin E, Rackham OJ, van Heesch S, Pua CJ, Kui M, et al. Titin-truncating variants affect heart function in disease cohorts and the general population. Nat Genet. 2017;49:46–53. DOI: 10.1038/ng.3719.

42. Khudhur ZO, Smail SW, Awla HK, Ahmed GB, Khdhir YO, Amin K, and Janson C. The effects of heavy smoking on oxidative stress, inflammatory biomarkers, vascular dysfunction, and hematological indices. Sci Rep. 2025;15:18251. DOI: 10.1038/s41598-025-03075-8.

43. Piano MR, Marcus GM, Aycock DM, Buckman J, Hwang CL, Larsson SC, Mukamal KJ, Roerecke M, on behalf the American Heart Association Council on L, Cardiometabolic H, et al. Alcohol Use and Cardiovascular Disease: A Scientific Statement From the American Heart Association. Circulation. 2025;152:e7–e21. DOI: 10.1161/CIR.0000000000001341.

44. Van Dam RM, Hu FB, and Willett WC. Coffee, Caffeine, and Health. N Engl J Med. 2020;383:369–78. DOI: 10.1056/NEJMra1816604.

45. Moukarzel M, Soroush K, Zhang Y, Gouin JP, Friedrich MG, and Luu JM. Sex Differences in the Relationship Between Psychosocial Stress and Myocardial Tissue Characteristics: A CMR Imaging Study. Circ Cardiovasc Imaging. 2025:e017667. DOI: 10.1161/CIRCIMAGING.124.017667.

46. Ong AD, Mann FD, and Kubzansky LD. Cumulative social advantage is associated with slower epigenetic aging and lower systemic inflammation. Brain Behav Immun Health. 2025;48:101096. DOI: 10.1016/j.bbih.2025.101096.

47. Littlejohns TJ, Holliday J, Gibson LM, Garratt S, Oesingmann N, Alfaro-Almagro F, Bell JD, Boultwood C, Collins R, Conroy MC, et al. The UK Biobank imaging enhancement of 100,000 participants: rationale, data collection, management and future directions. Nat Commun. 2020;11:2624. DOI: 10.1038/s41467-020-15948-9.

48. Argentieri MA, Amin N, Nevado-Holgado AJ, Sproviero W, Collister JA, Keestra SM, Kuilman MM, Ginos BNR, Ghanbari M, Doherty A, et al. Integrating the environmental and genetic architectures of aging and mortality. Nat Med. 2025;31:1016–25. DOI: 10.1038/s41591-024-03483-9.

49. Benjamini Y and Hochberg Y. Controlling the False Discovery Rate: A Practical and Powerful Approach to Multiple Testing. J R Stat Soc Series B Stat Methodol. 1995;57:289–300. DOI: 10.1111/j.2517-6161.1995.tb02031.x.

50. McGurk KA, Curran L, Sau A, Ng FS, Halliday B, Ware JS, and O’Regan DP. Circulating Cardiovascular Proteomic Associations With Genetics and Disease. Circ Genom Precis Med. 2025:e005005. DOI: 10.1161/CIRCGEN.124.005005.

51. Josephs KS, Seaby EG, May P, Theotokis P, Yu J, Andreou A, Sinclair H, Morris-Rosendahl D, Thomas ERA, Ennis S, et al. Cardiomyopathies in 100,000 genomes project: interval evaluation improves diagnostic yield and informs strategies for ongoing gene discovery. Genome Med. 2024;16:125. DOI: 10.1186/s13073-024-01390-9.

52. Ingles J, Goldstein J, Thaxton C, Caleshu C, Corty EW, Crowley SB, Dougherty K, Harrison SM, McGlaughon J, Milko LV, et al. Evaluating the Clinical Validity of Hypertrophic Cardiomyopathy Genes. Circ Genom Precis Med. 2019;12:e002460. DOI: 10.1161/CIRCGEN.119.002460.

53. Whiffin N, Minikel E, Walsh R, O’Donnell-Luria AH, Karczewski K, Ing AY, Barton PJR, Funke B, Cook SA, MacArthur D, et al. Using high-resolution variant frequencies to empower clinical genome interpretation. Genet Med. 2017;19:1151–8. DOI: 10.1038/gim.2017.26.

54. Petersen SE, Matthews PM, Francis JM, Robson MD, Zemrak F, Boubertakh R, Young AA, Hudson S, Weale P, Garratt S, et al. UK Biobank’s cardiovascular magnetic resonance protocol. J Cardiovasc Magn Reson. 2016;18:8. DOI: 10.1186/s12968-016-0227-4.

55. Bai W, Suzuki H, Qin C, Tarroni G, Oktay O, Matthews PM, and Rueckert D. Recurrent Neural Networks for Aortic Image Sequence Segmentation with Sparse Annotations. Medical Image Computing and Computer Assisted Intervention – MICCAI 2018. 2018:586–94. DOI: 10.1007/978-3-030-00937-3_67.

56. Bai W, Suzuki H, Huang J, Francis C, Wang S, Tarroni G, Guitton F, Aung N, Fung K, Petersen SE, et al. A population-based phenome-wide association study of cardiac and aortic structure and function. Nat Med. 2020;26:1654–62. DOI: 10.1038/s41591-020-1009-y.

57. Bai W, Shi W, de Marvao A, Dawes TJ, O’Regan DP, Cook SA, and Rueckert D. A bi-ventricular cardiac atlas built from 1000+ high resolution MR images of healthy subjects and an analysis of shape and motion. Med Image Anal. 2015;26:133–45. DOI: 10.1016/j.media.2015.08.009.

58. Garland M and Heckbert PS. Surface simplification using quadric error metrics. In: Proceedings of the 24th annual conference on Computer graphics and interactive techniques. 1997:209–16.

59. Higgins I, Matthey L, Pal A, Burgess C, Glorot X, Botvinick M, Mohamed S, and Lerchner A. beta-VAE: Learning basic visual concepts with a constrained variational framework. In: International conference on learning representations. 2017.

60. Tran D, Wang H, Torresani L, Ray J, LeCun Y, and Paluri M. A closer look at spatiotemporal convolutions for action recognition. In: Proceedings of the IEEE conference on Computer Vision and Pattern Recognition. 2018:6450–9.

61. Nealen A, Igarashi T, Sorkine O, and Alexa M. Laplacian mesh optimization. In: Proceedings of the 4th international conference on Computer graphics and interactive techniques in Australasia and Southeast Asia. 2006:381–9.

62. Fey M and Lenssen JE. Fast graph representation learning with PyTorch Geometric. *arXiv*:1903.02428. 2019. DOI: 10.48550/arXiv.1903.02428.

63. Kingma DP and Ba J. Adam: A method for stochastic optimization. *arXiv:1412*.6980. 2014. DOI: 10.48550/arXiv.1412.6980.

64. Akiba T, Sano S, Yanase T, Ohta T, and Koyama M. Optuna: A Next-Generation Hyperparameter Optimization Framework. In: The 25th ACM SIGKDD International Conference on Knowledge Discovery & Data Mining. 2019:2623–31.

65. Mbatchou J, Barnard L, Backman J, Marcketta A, Kosmicki JA, Ziyatdinov A, Benner C, O’Dushlaine C, Barber M, Boutkov B, et al. Computationally efficient whole-genome regression for quantitative and binary traits. Nat Genet. 2021;53:1097–103. DOI: 10.1038/s41588-021-00870-7.

66. Purcell S, Neale B, Todd-Brown K, Thomas L, Ferreira MA, Bender D, Maller J, Sklar P, de Bakker PI, Daly MJ, et al. PLINK: a tool set for whole-genome association and population-based linkage analyses. Am J Hum Genet. 2007;81:559–75. DOI: 10.1086/519795.

67. Shi S, Rubinacci S, Hu S, Moutsianas L, Stuckey A, Need AC, Palamara PF, Caulfield M, Marchini J, and Myers S. A Genomics England haplotype reference panel and imputation of UK Biobank. Nat Genet. 2024;56:1800–3. DOI: 10.1038/s41588-024-01868-7.

68. Hernán MA, Hernández-Díaz S, and Robins JM. A structural approach to selection bias. Epidemiology. 2004;15:615–25. DOI: 10.1097/01.ede.0000135174.63482.43.

69. Schisterman EF, Cole SR, and Platt RW. Overadjustment bias and unnecessary adjustment in epidemiologic studies. Epidemiology. 2009;20:488–95. DOI: 10.1097/EDE.0b013e3181a819a1.

70. Hocker JD, Poirion OB, Zhu F, Buchanan J, Zhang K, Chiou J, Wang TM, Zhang Q, Hou X, Li YE, et al. Cardiac cell type-specific gene regulatory programs and disease risk association. Sci Adv. 2021;7. DOI: 10.1126/sciadv.abf1444.

71. Sheth MU, Qiu WL, Rosa Ma X, Gschwind AR, Jagoda E, Tan AS, Einarsson H, Gorissen BL, Dubocanin D, McGinnis CS, et al. Mapping enhancer-gene regulatory interactions from single-cell data. bioRxiv. 2024. DOI: 10.1101/2024.11.23.624931.

72. Ma XR, Conley SD, Kosicki M, Bredikhin D, Cui R, Tran S, Sheth MU, Qiu WL, Chen S, Kundu S, et al. Molecular convergence of risk variants for congenital heart defects leveraging a regulatory map of the human fetal heart. medRxiv. 2024. DOI: 10.1101/2024.11.20.24317557.

73. Fulco CP, Nasser J, Jones TR, Munson G, Bergman DT, Subramanian V, Grossman SR, Anyoha R, Doughty BR, Patwardhan TA, et al. Activity-by-contact model of enhancer-promoter regulation from thousands of CRISPR perturbations. Nat Genet. 2019;51:1664–9. DOI: 10.1038/s41588-019-0538-0.

74. McLaren W, Gil L, Hunt SE, Riat HS, Ritchie GR, Thormann A, Flicek P, and Cunningham F. The Ensembl Variant Effect Predictor. Genome Biol. 2016;17:122. DOI: 10.1186/s13059-016-0974-4.

75. Mountjoy E, Schmidt EM, Carmona M, Schwartzentruber J, Peat G, Miranda A, Fumis L, Hayhurst J, Buniello A, Karim MA, et al. An open approach to systematically prioritize causal variants and genes at all published human GWAS trait-associated loci. Nat Genet. 2021;53:1527–33. DOI: 10.1038/s41588-021-00945-5.

76. GTEx Consortium. The GTEx Consortium atlas of genetic regulatory effects across human tissues. Science. 2020;369:1318–30. DOI: 10.1126/science.aaz1776.

77. Moore JE, Pratt HE, Fan K, Phalke N, Fisher J, Elhajjajy SI, Andrews G, Gao M, Shedd N, Fu Y, et al. An expanded registry of candidate cis-regulatory elements. Nature. 2026. DOI: 10.1038/s41586-025-09909-9.

78. Zou Y, Carbonetto P, Wang G, and Stephens M. Fine-mapping from summary data with the “Sum of Single Effects” model. PLoS Genet. 2022;18:e1010299. DOI: 10.1371/journal.pgen.1010299.

79. Wang G, Sarkar A, Carbonetto P, and Stephens M. A simple new approach to variable selection in regression, with application to genetic fine mapping. J R Stat Soc Series B Stat Methodol. 2020;82:1273–300. DOI: 10.1111/rssb.12388.

80. Trapnell C, Cacchiarelli D, Grimsby J, Pokharel P, Li S, Morse M, Lennon NJ, Livak KJ, Mikkelsen TS, and Rinn JL. The dynamics and regulators of cell fate decisions are revealed by pseudotemporal ordering of single cells. Nat Biotechnol. 2014;32:381–6. DOI: 10.1038/nbt.2859.

