## Supplementary Materials for "Population genomics of four-dimensional cardiac motion reveals non-myocyte regulatory programmes"

### Supplemental Methods

#### Exposure factors

**Variable coding.** Variable recoding was performed before multiple imputation. All variable responses of "Prefer not to answer", "Do not know", and "Not applicable (NA)" were recoded to NA. Following multiple imputation, any variable values where the participant responded "Prefer not to answer" in the original dataset were recoded to NA. All nominal categorical variables were coded as unordered factors, with the reference level set as the most frequent response reported in the UK Biobank dataset. Ordered categorical variables were coded with the reference level set as the lowest response (e.g., "Never", "Rarely", or "None"). Dichotomous categorical variables were coded with the reference level set as "No." All variables with "mark all that apply" response categories were converted into multiple dummy variables, with each unique response option used to create a yes/no dichotomous variable.

For all numeric diet intake variables and number of hours spent on the mobile phone, participants who responded "less than one" were recoded as 0.5 hours. For smoking pack years (field ID 20161), missing values were recoded as 0 if the respondent was coded as "No" in response to the derived ever smoked variable available in the UK Biobank (field ID 20160), but otherwise left as NA if respondents had ever smoked.

**Imputation.** Missing data were imputed using the R package *missRanger*, which combines random forest imputation with predictive mean matching. The dataset was imputed with a maximum of 10 iterations, 200 trees per random forest, and 16 threads, while all other hyperparameters were kept at their default settings. Variables were recoded or derived after imputation.

**Recoding after imputation.** For processed meat (field ID 1349), the top three frequencies were combined to obtain four categories: never, < 1.0 time per week, 1.0 time per week, and ≥ 2.0 times per week. For cheese intake (field ID 1408), the bottom two and the top two frequencies were combined to get four categories: < 1.0 time per week, 1.0 time per week, 2.0–4.9 times per week, and ≥ 5.0 times per week. For daily coffee intake (field ID 1498), participants were grouped as: 0 cups/day, 0.5–1.9 cups per day, 2.0–2.9 cups per day, and ≥ 3.0 cups per day.

For alcohol intake frequency (field ID 1558), all participants who responded as "Never" or "Previous" drinkers to the alcohol status variable (field ID 20117) were coded as NA. In addition, we coded participants who responded as drinking on "Special occasions only" as NA. The final variable was coded as a nominal variable with responses for "One to three times a month", "Once or twice a week", "Three or four times a week", and "Daily or almost daily", with "One to three times a month" set as the reference.

**Derived variables.** All derived variables were calculated after imputation, and the original variables used to create each of the derived variables were then excluded from the multivariable regression analysis.

**Sleep.** A categorical variable for hours of sleep was derived using the UK Biobank hours of sleep continuous measure (field ID 1160). Category levels used for hours of sleep were: <7 hours, 7-9 hours, and >9 hours, with the reference set as 7-9 hours.

**Total red meat consumption.** A total red meat consumption variable was created by summing the frequencies for beef (field ID 1369), pork (field ID 1389), and lamb/mutton (field ID 1379). and coded into 4 categories of red meat consumption: <1 time per week, 1.0-1.9 times per week, 2.0-2.9 times per week, and ≥ 3.0 times per week.

**Air pollution.** The UK Biobank provides data on local environmental exposures. Annual average concentrations of air pollutants in the UK Biobank were estimated by the land use regression model where geographic information system data is used to estimate the spatial variation of annual average concentrations of air pollutants around the home addresses of participants. For this study, we considered particulate matter with an aerodynamic diameter smaller than 2.5 $\mu$m (field ID 24004), nitrogen oxide levels in 2010 (field ID 24004) and nitrogen dioxide air pollution levels in 2005 (field ID 24016), 2006 (field ID 24017), 2007 (field ID 24018), and 2010 (field ID 24003). Nitrogen dioxide values from the four years were summed and evaluated as a continuous variable to assess their association with each latent feature. Additionally, the summed values were divided into tertiles and analyzed as a categorical variable to explore their effects on heart shape and motion (Supplementary Table 3).

##### Risk factor enrichment.

Cardiovascular events and outcomes were collected from participants' linked healthcare records, obtained from primary and secondary healthcare sources. Cardiovascular events were derived from ICD-9, ICD-10, "first occurrence ICD-10" and operational (OPSC-4) datasets, and self-reported patient questionnaires. Each of these datasets were mapped to pre-defined diagnostic terms (Supplementary Table 9), which were used for our analyses. Logistic and linear regression models were fitted to explore the relationship between the reduced dimensions of the tree ($c1$ and $c2$ coordinates) and clinical features of interest. Linear models were fitted to predict the risk distribution across the tree (supplementary Figure 15).

#### DDTree modelling of motion latent components

**Statistical modeling and association analyses of DDRTree embedding.** The distribution of nine key phenotypic variables was overlaid onto the tree structure to visualize their relative enrichment (Supplementary Figure 12a). Phenotypic structure was further quantified using Moran's I and linear regression analyses (Supplementary Figure 12b,c), defining a quantitative taxonomy of 4D cardiac motion diversity across imaging-derived phenotypes.

The predictive power of the DDRTree mapping was evaluated using generalised additive models (GAMs) fitted on the DDRTree coordinates ($c_1$ and $c_2$) (see Figure 11a). Logistic GAMs were used for binary outcomes (pathogenic or likely pathogenic genotype) and Gaussian GAMs for continuous cardiac traits, with 5-fold cross-validation to assess predictive performance. A Cox proportional hazards model was trained using the DDRTree coordinates ($c_1$ and $c_2$) for each individual as predictors of time to major adverse cardiac events (MACE). Individual 5-year survival probabilities were derived from the fitted model and used to visualise spatial variation in MACE risk across the tree-structured latent space (see Figure 11a).

A separate generalised additive model (GAM) was fitted to model changes in expression (*LogFC*) from the start to the end of each lineage, allowing assessment of differential expression across pseudotime. P values were adjusted using the Benjamini–Hochberg method, and significant *LogFC* values were plotted for each branch (Figure 11b).

In addition, the logistic GAM for cardiac outcomes in Supplementary Figure 15a revealed a structured variation in risk across the latent space, indicating that individuals with similar latent coordinates tend to share comparable cardiac risk profiles. The cardiovascular outcomes observed in the cohort are summarised in Supplementary Table 11.

**Branch-level phenotypic association analyses.** To identify phenotypic heterogeneity across the tree, both continuous and discrete traits were tested for branch-level enrichment. Associations between tree branches and clinical, imaging-derived, and genetic traits were assessed using non-parametric tests. One-vs-all comparisons were performed to test for branch-level enrichment: for continuous variables, by the Wilcoxon rank-sum test; and for discrete variables, by Fisher's exact test (see Supplementary Figure 13). For continuous variables, a global Kruskal–Wallis test was first applied, followed by one-vs-all and pairwise Wilcoxon rank-sum tests to identify branch-specific differences (see Supplementary Figure 14). All *P* values were adjusted for multiple testing using the Benjamini–Hochberg false discovery rate (FDR) method.

**Associations between DDRTree axes and cardiovascular outcomes.** Associations between standardised DDRTree axes ($c_1$, $c_2$) and incident cardiovascular outcomes were evaluated using point–biserial correlations and univariate logistic regression (Supplementary Figure 15b,c). Each outcome was binarized (case/control), and effects were expressed as log-odds ratios with 95% confidence intervals per one–standard-deviation (1 SD) increase in axis value. Forest plots summarise the direction and magnitude of associations across outcomes.

**Generalisation of the trained tree structure to unseen participants.** To evaluate the generalisation of the learned 4D cardiac motion manifold, latent representations from unseen UK Biobank participants were projected into the low-dimensional space defined by the DDRTree model trained on the development cohort. The linear mapping $W$ learned during training was applied to obtain low-dimensional representations of the latent motion features, which were then used to predict the corresponding DDRTree coordinates.

Two Random Forest regression models were trained separately to predict the tree coordinates ($c_1$ and $c_2$) from these mapped features. Each model was implemented using 100 decision trees with a maximum depth of 10 and a fixed random seed to ensure reproducibility. Model performance was evaluated using five-fold cross-validation on the training cohort, computing the coefficient of determination ($R^2$), mean squared error (MSE), and mean absolute error (MAE) across training and validation folds (reported in Supplemental Table 8). The trained Random Forest models were subsequently applied to the unseen test cohort to infer tree coordinates based on their mapped latent motion representations.

Faithfulness of the tree mapping for unseen UK Biobank participants was evaluated by assessing the statistical consistency between the training and test cohorts (see Supplementary Table 8). The Spearman correlation of coordinate summary statistics was computed to quantify concordance between cohorts. In addition, trustworthiness metric was used to evaluate how well local neighborhood relationships from the original high-dimensional latent space ($z$) were preserved in the low-dimensional tree embedding ($c$), with values ranging from 0 to 1 (higher values indicating greater faithfulness of the mapping). Distributional similarity was assessed using the two-sample Kolmogorov–Smirnov test for differences in cumulative distributions and the Wilcoxon rank-sum test for median differences. To further quantify divergence between coordinate distributions, we calculated the Wasserstein (Earth Mover's) distance, Jensen–Shannon divergence, and Cohen's $d$ effect size. Finally, an overlap coefficient was computed to estimate the proportion of shared density between the training and test distributions. Together, these complementary metrics characterise the consistency and faithfulness of the DDRTree manifold mapping when applied to unseen 4D cardiac motion data.

**Risk prediction and model evaluation** Risk prediction for 5-year major adverse cardiovascular events (MACE) was assessed using Cox proportional hazards models. Models were constructed using conventional imaging measures (LVEF, LVEDVi and Ell global), motion-derived latent coordinates ($c_1$ and $c_2$), and their combination. Model discrimination was evaluated using the C-index and time-dependent area under the receiver operating characteristic curve (AUC) at 5 years. The C-index was corrected for optimism using bootstrap resampling (100 iterations). Differences in AUC between models were assessed using pairwise comparisons on the same dataset with the *riskRegression* package.

### Supplemental Figures

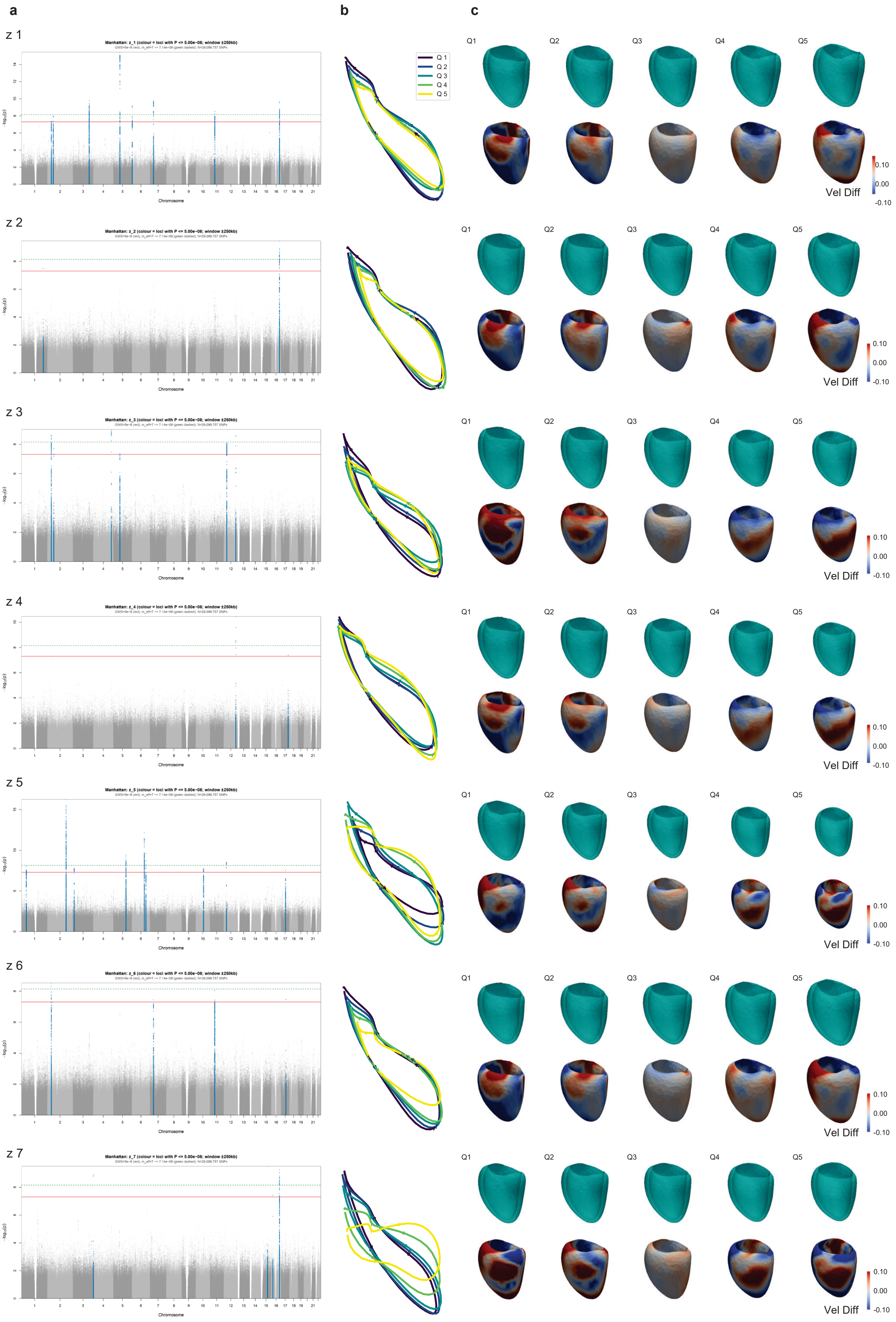


**Supplementary Figure 1. Genetic associations and phenotypic variation of latent motion components.**

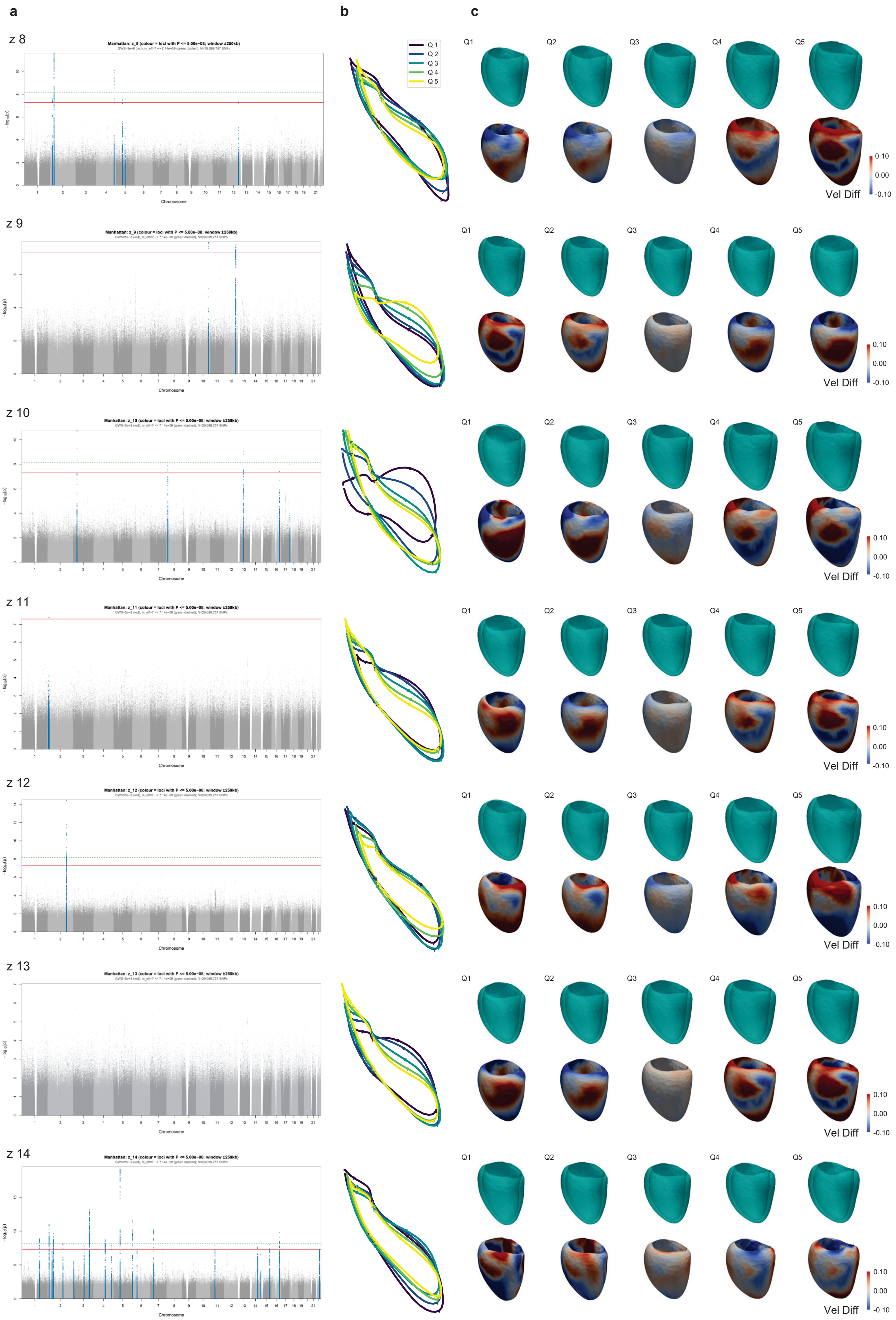


**Supplementary Figure 1. Genetic associations and phenotypic variation of latent motion components.**

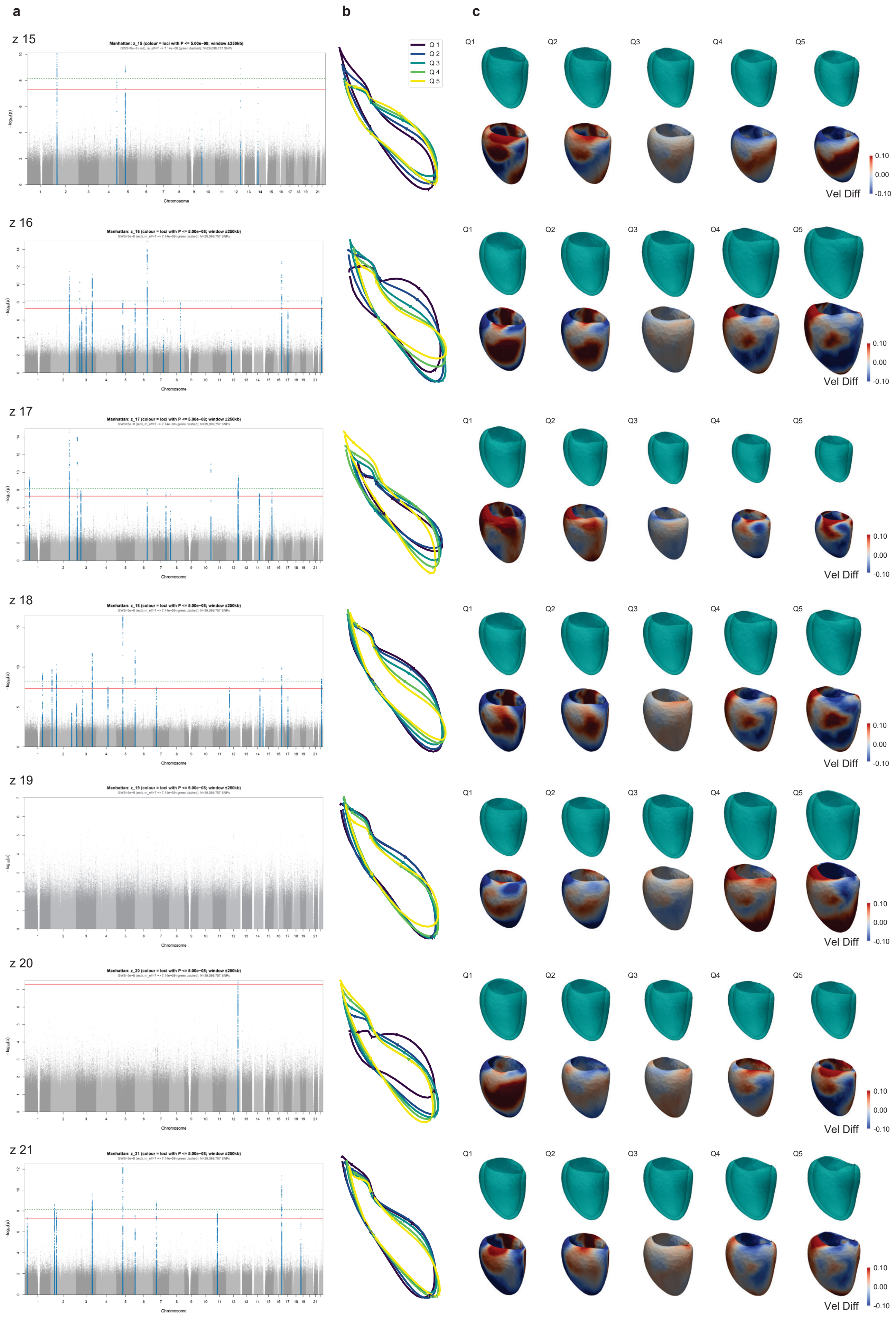


**Supplementary Figure 1. Genetic associations and phenotypic variation of latent motion components.**

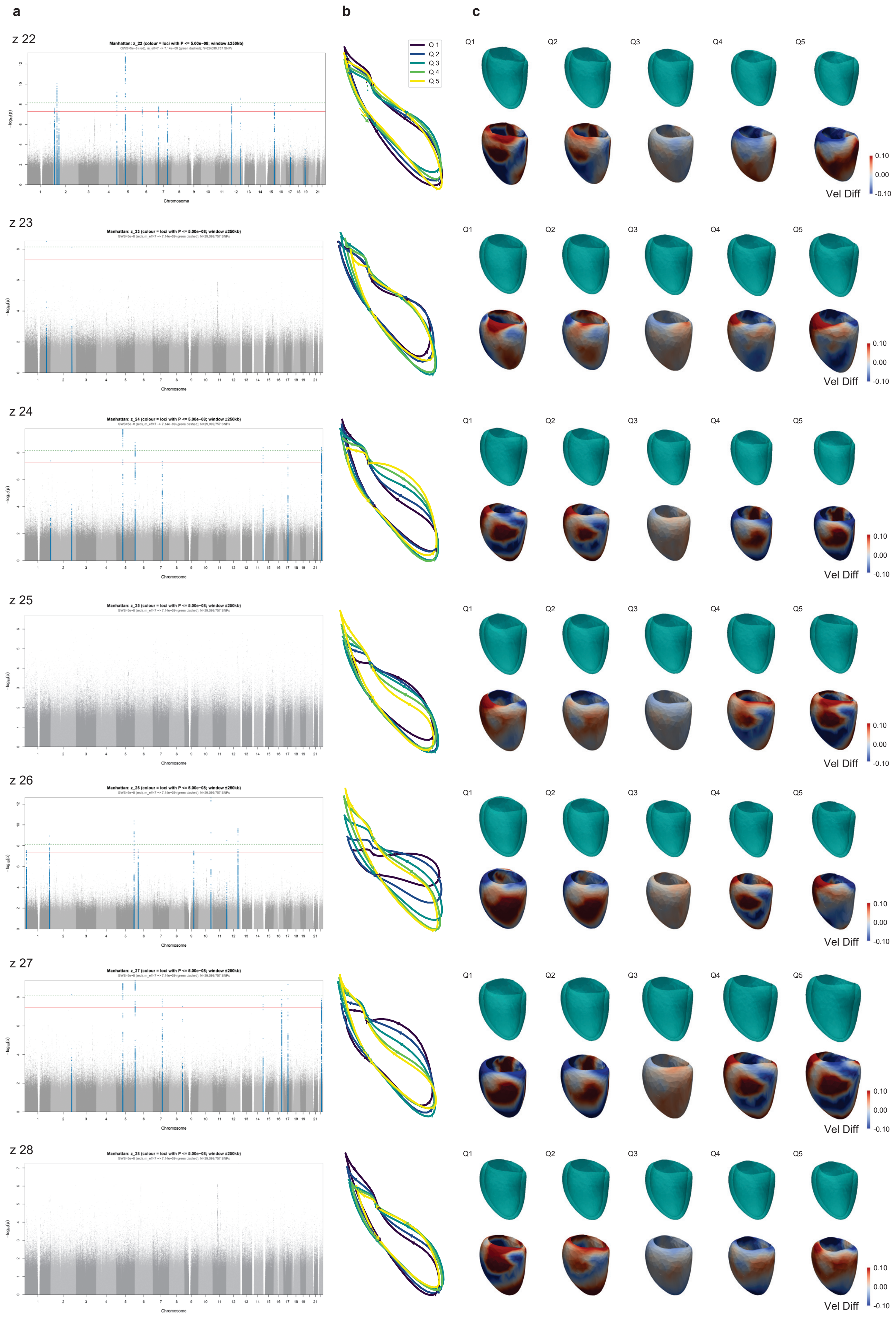


**Supplementary Figure 1. Genetic associations and phenotypic variation of latent motion components.**

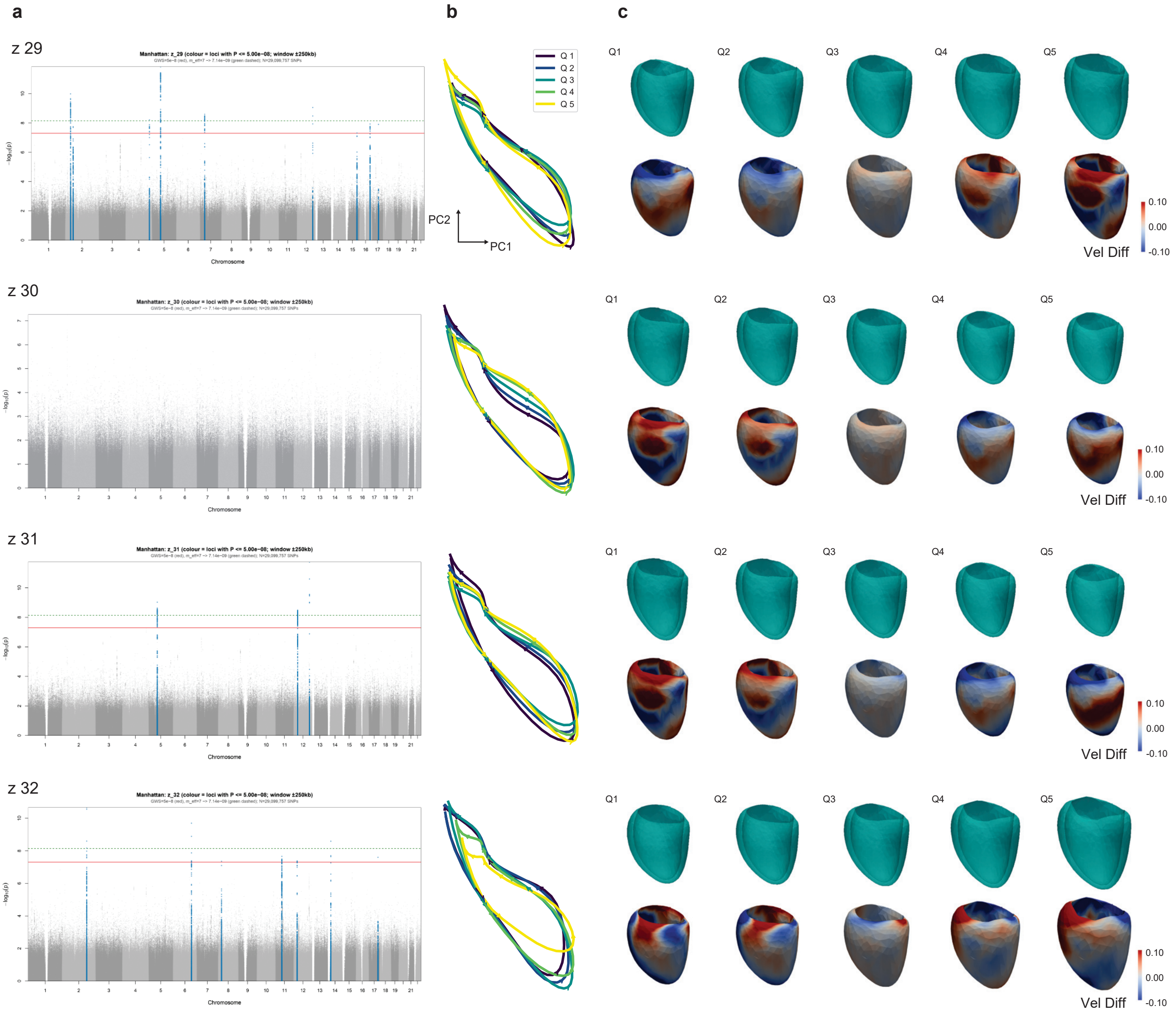


**Supplementary Figure 1. Genetic associations and phenotypic variation of latent motion components ($z_1$-$z_{32}$). a,** Manhattan plots from genome-wide association studies (GWAS) performed on latent motion components, highlighting loci surpassing genome-wide significance. The red line denotes the conventional genome-wide significance threshold ($P = 5 \times 10^{-8}$; $-\log_{10} P = 7.3$), and the green dashed line indicates the $M_{\text{eff}}$-corrected threshold ($P = 7.14 \times 10^{-9}$; $-\log_{10} P = 8.15$). **b,** Two-dimensional PCA representations of average cardiac motion trajectories across quantile ranges of each latent componenet (Quantiles from Q1 to Q5: [0.00–0.02], [0.09–0.11], [0.49–0.51], [0.89–0.91], and [0.98–1.00]), illustrating progressive variation along the latent dimension. **c,** Corresponding cardiac shapes and motion patterns across quantile ranges of each latent componenet. Cyan colored meshes show the mean end-diastolic (ED) geometry. Color-coded meshes show velocity differences (mm/frame) at ED relative to a control (non-carrier) reference cohort, with warm and cool colors indicating increased and decreased velocities, respectively.

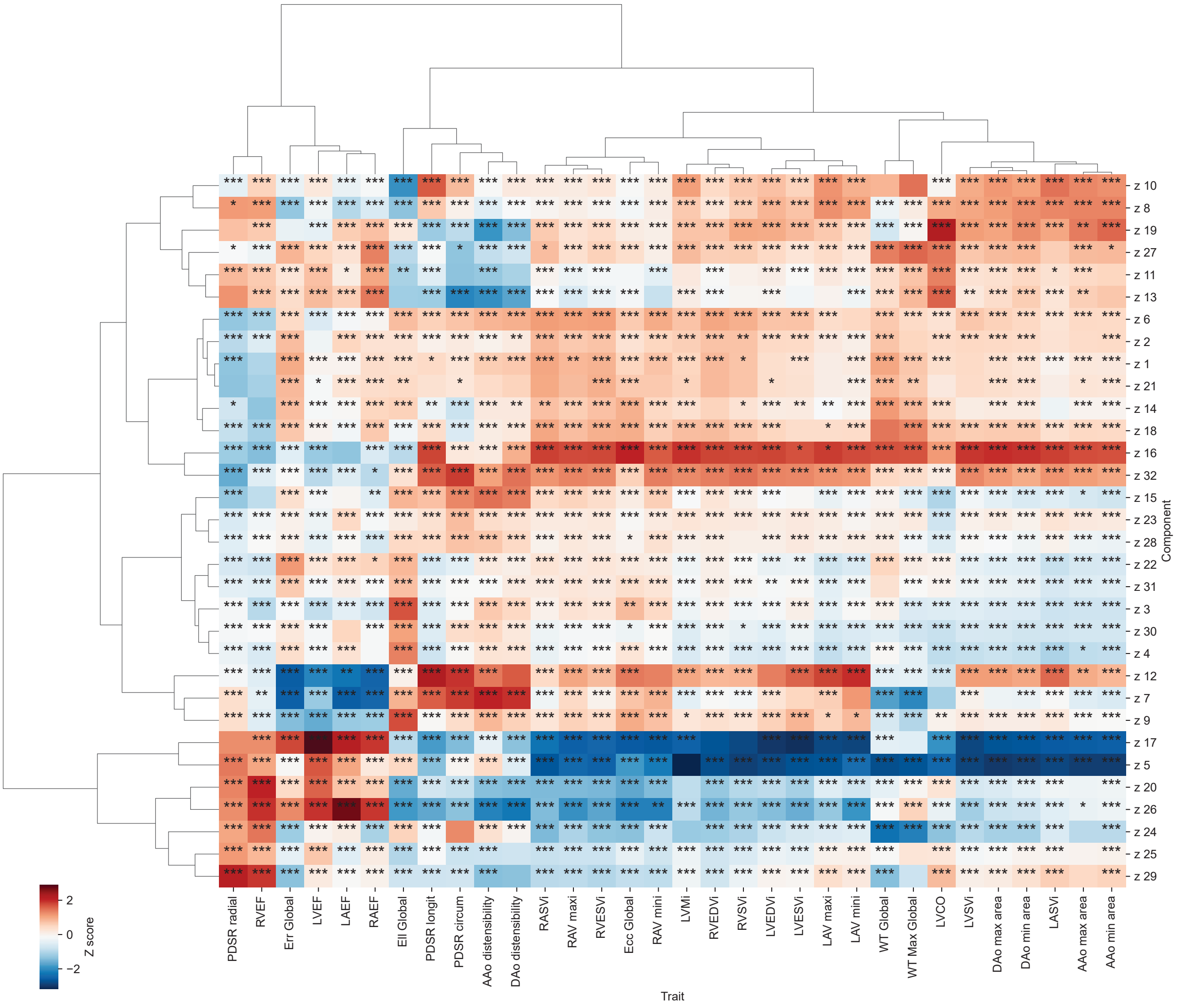


**Supplementary Figure 2. FDR corrected significant association of latent components with Imaging traits.** Asterisks denote FDR-significant associations (*FDR $\leq$ 0.05, **FDR $\leq$ 0.01, ***FDR $\leq$ 0.001). Colors indicate standardised beta values (Z scores). AAo, ascending aorta; DAo, descending aorta; Ell, longitudinal strain; Ecc, circumferential strain; Err, radial strain; FDR, false-discovery rate; LAEF, left atrial ejection fraction; LASVi, left atrial stroke volume indexed; LAVmax, maximum left atrial volume indexed; LAVmin, minimum left atrial volume indexed; LV, left ventricle; LVCO, left ventricular cardiac output; LVCi, left ventricular cardiac index; LVEDVi, left ventricular end-diastolic volume indexed; LVEF, left ventricular ejection fraction; LVESVi, left ventricular end-systolic volume indexed; LVMi, left ventricular mass indexed; LVSVi, left ventricular stroke volume indexed; PDSR, peak diastolic strain rate; RAEF, right atrial ejection fraction; RASVi, right atrial stroke volume indexed; RAVmax, maximum right atrial volume indexed; RAVmin, minimum right atrial volume indexed; RV, right ventricle; RVEDVi, right ventricular end-diastolic volume indexed; RVEF, right ventricular ejection fraction; RVESVi, right ventricular end-systolic volume indexed; RVSVi, right ventricular stroke volume indexed; WT, myocardial wall thickness.

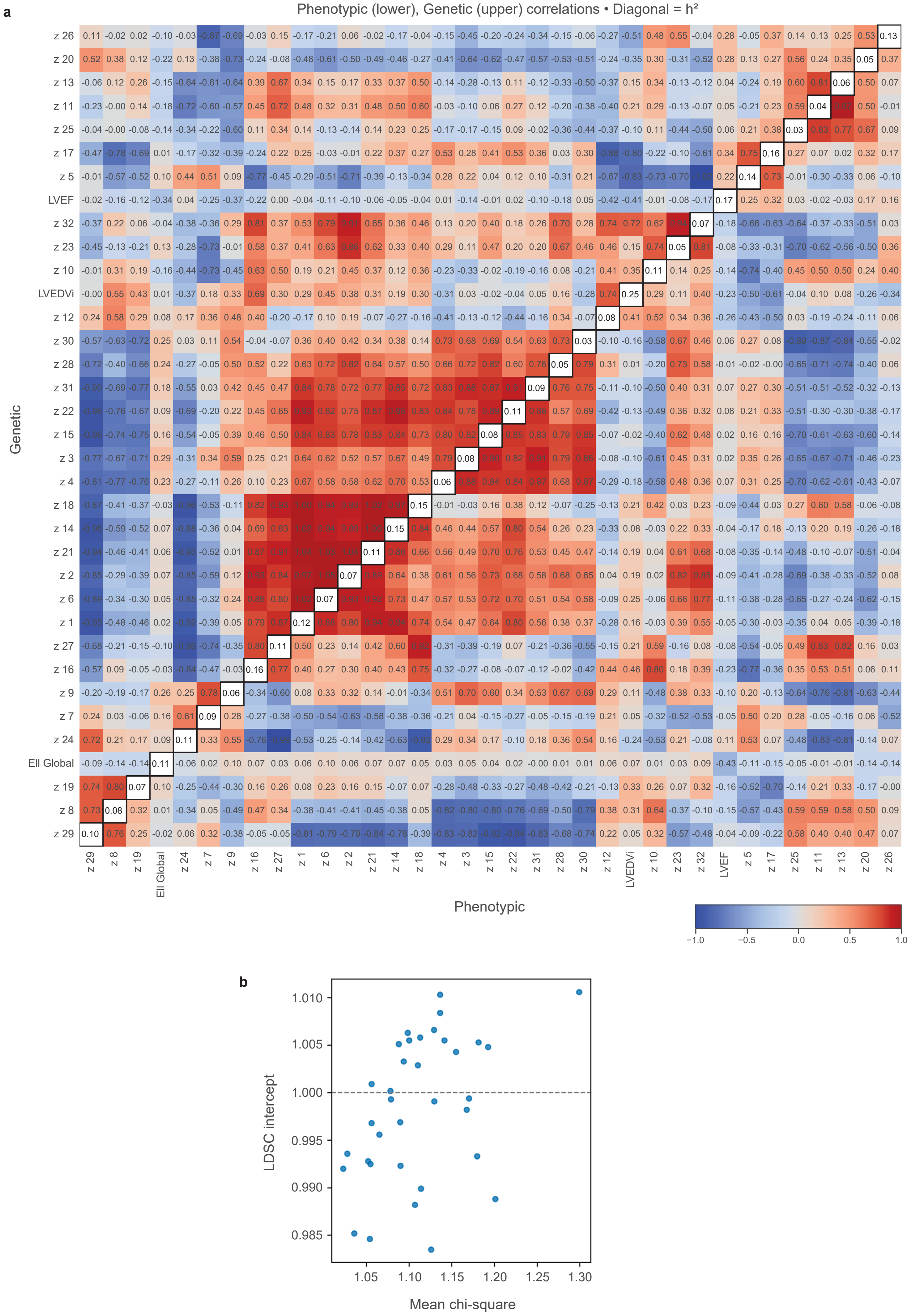

a
Phenotypic (lower), Genetic (upper) correlations • Diagonal = h²
Genetic
Phenotypic
z 26
z 20
z 13
z 11
z 25
z 17
z 5
LVEF
z 32
z 23
z 10
LVEDVi
z 12
z 30
z 28
z 31
z 22
z 15
z 3
z 4
z 18
z 14
z 21
z 2
z 6
z 1
z 27
z 16
z 9
z 7
z 24
EII Global
z 19
z 8
z 29
−1.0
−0.5
0.0
0.5
1.0
b
LDSC intercept
Mean chi-square
1.010
1.005
1.000
0.995
0.990
0.985
1.05
1.10
1.15
1.20
1.25
1.30

**Supplementary Figure 3. Phenotypic and genetic architecture of left-ventricular motion traits.** **a**, Heat map showing pairwise correlations among 32 latent motion traits ($z_1 - z_{32}$) and three conventional indices (LVEF, Ell Global, LVEDVi), with traits ordered by hierarchical clustering on the bivariate genetic correlation matrix (average-linkage clustering using the distance $D = (1 - r_g)/2$). The lower triangle displays pairwise phenotypic correlations (Spearman's $\rho$), whereas the upper triangle displays genetic correlations estimated by bivariate LD score regression from the corresponding pairs of GWAS summary statistics. The diagonal is annotated with SNP heritability ($h^2$) for each phenotype derived from univariate LDSC. The near-identical clustering structure between phenotypic and genetic matrices suggest that the latent representation reflects stable, biologically grounded axes of cardiac motion. **b**, LDSC model diagnostics showing the relationship between mean $\chi^2$ and LDSC intercept across all 35 traits. Intercepts cluster tightly around 1.0, indicating minimal confounding from population stratification or model misspecification and supporting the validity of the genetic correlation estimates.

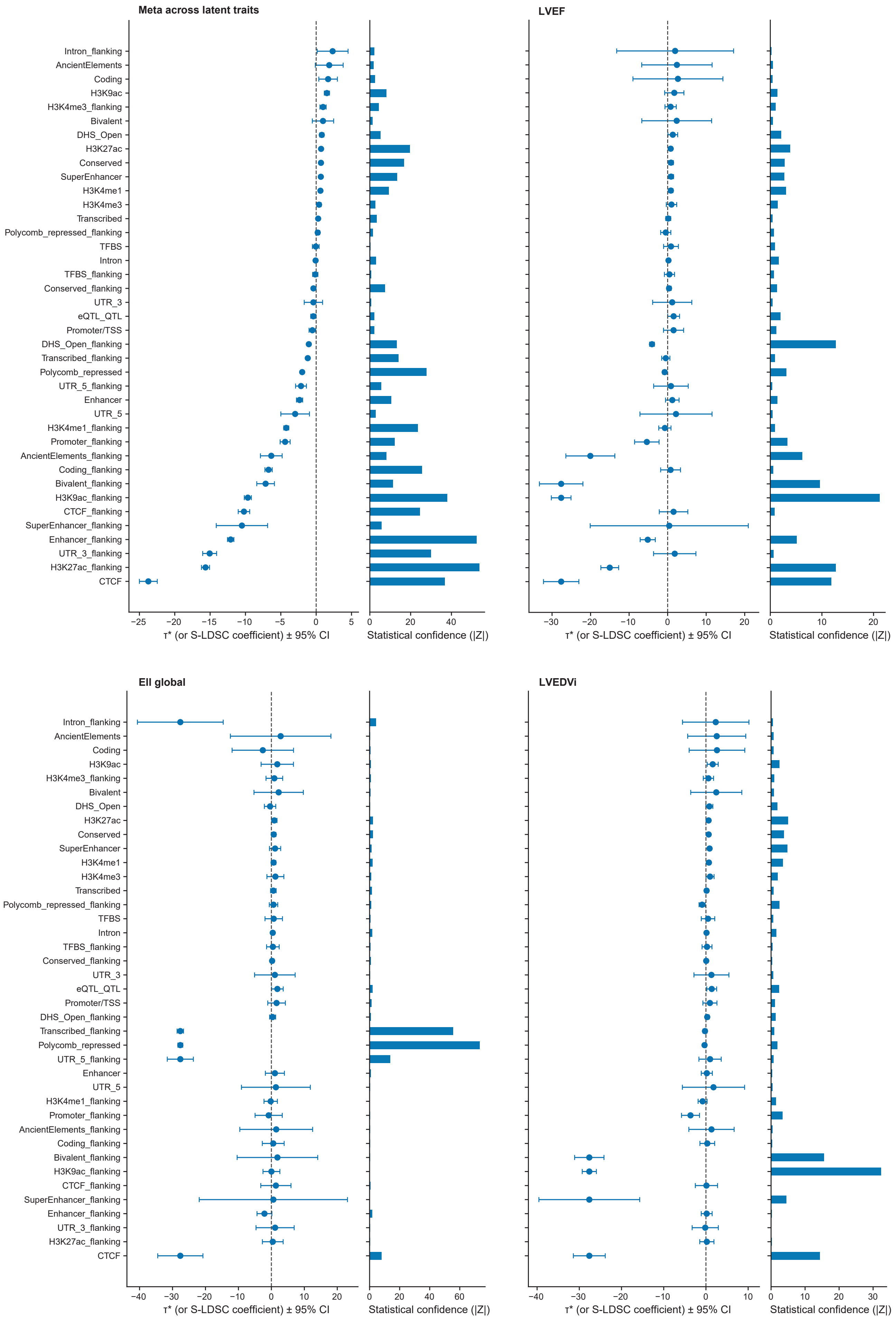

Meta across latent traits
LVEF
EII global
LVEDVi
Intron_flanking
AncientElements
Coding
H3K9ac
H3K4me3_flanking
Bivalent
DHS_Open
H3K27ac
Conserved
SuperEnhancer
H3K4me1
H3K4me3
Transcribed
Polycomb_repressed_flanking
TFBS
Intron
TFBS_flanking
Conserved_flanking
UTR_3
eQTL_QTL
Promoter/TSS
DHS_Open_flanking
Transcribed_flanking
Polycomb_repressed
UTR_5_flanking
Enhancer
UTR_5
H3K4me1_flanking
Promoter_flanking
AncientElements_flanking
Coding_flanking
Bivalent_flanking
H3K9ac_flanking
CTCF_flanking
SuperEnhancer_flanking
Enhancer_flanking
UTR_3_flanking
H3K27ac_flanking
CTCF
τ* (or S-LDSC coefficient) ± 95% CI
Statistical confidence (|Z|)

**Supplementary Figure 4. Partitioning heritability across regulatory annotations with S-LDSC.** Grouped functional annotations from the baselineLD v2.2 model were used to quantify the contribution of distinct regulatory features to SNP heritability for the 32 latent motion traits and three conventional metrics (LVEF, Ell Global, LVEDVi). Annotations were collapsed into biologically coherent families, and were ranked according to the meta-analysis across latent traits for comparability. For each annotation, the point estimate represents the stratified LD score regression coefficient ($\tau^*$) with 95% confidence intervals, and the accompanying horizontal bar encodes statistical confidence as the absolute Z-score. These analyses demonstrate that motion-derived latent traits index reproducible and biologically interpretable components of regulatory architecture, supporting their use as sensitive endophenotypes for downstream functional mapping.

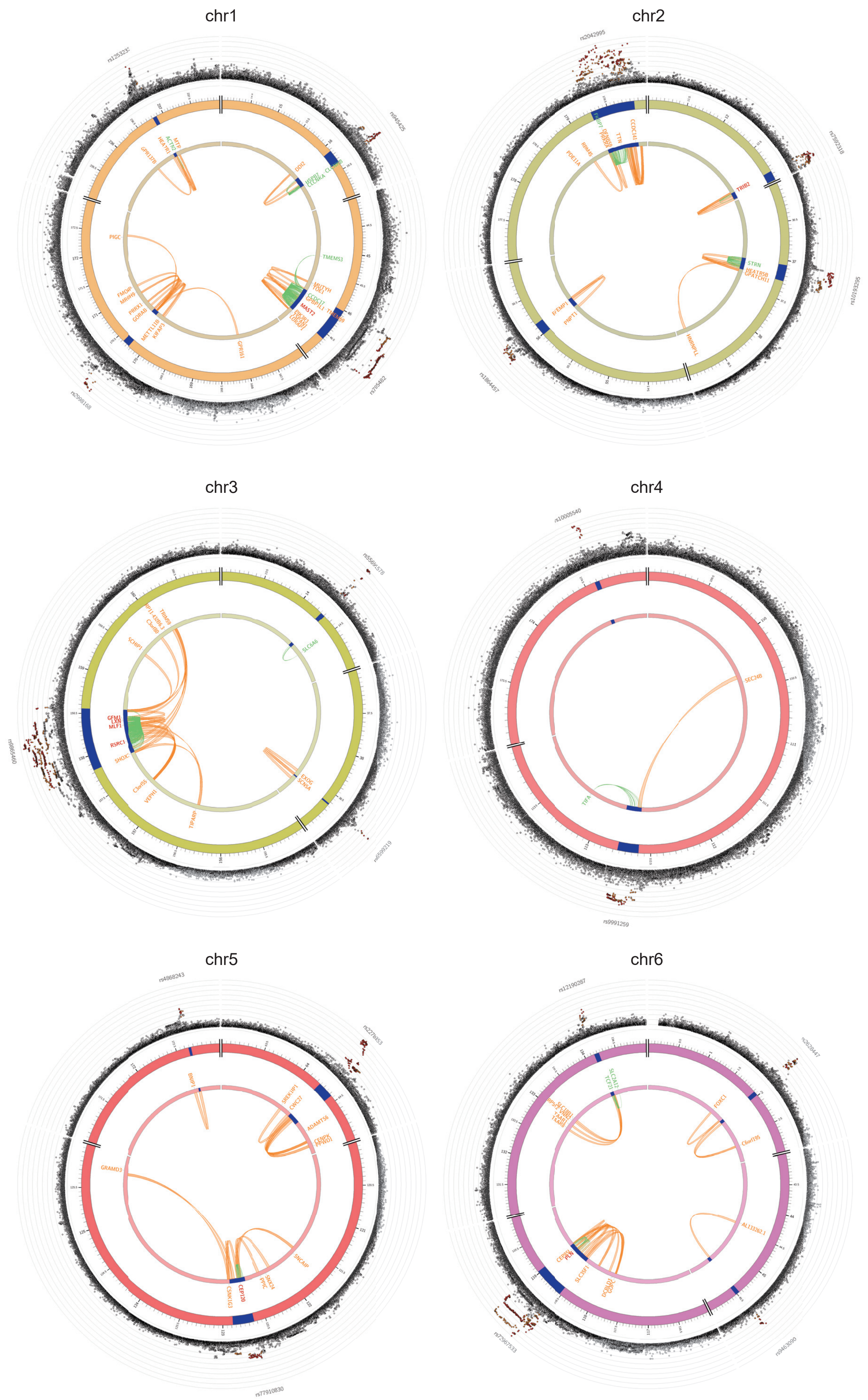


**Supplementary Figure 5. Circos plots of genomic loci and functional mapping for latent motion traits.**

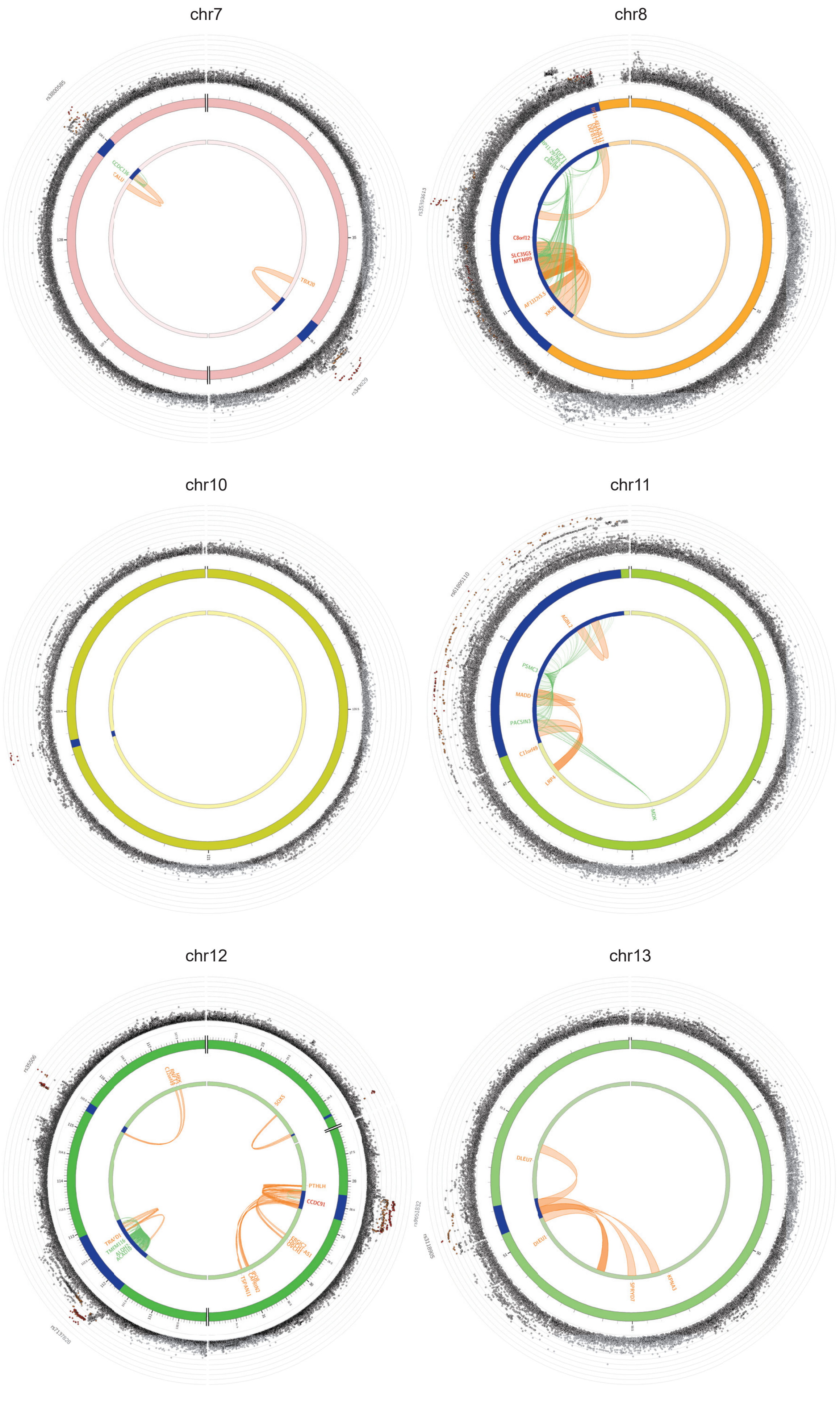


**Supplementary Figure 5. Circos plots of genomic loci and functional mapping for latent motion traits.**

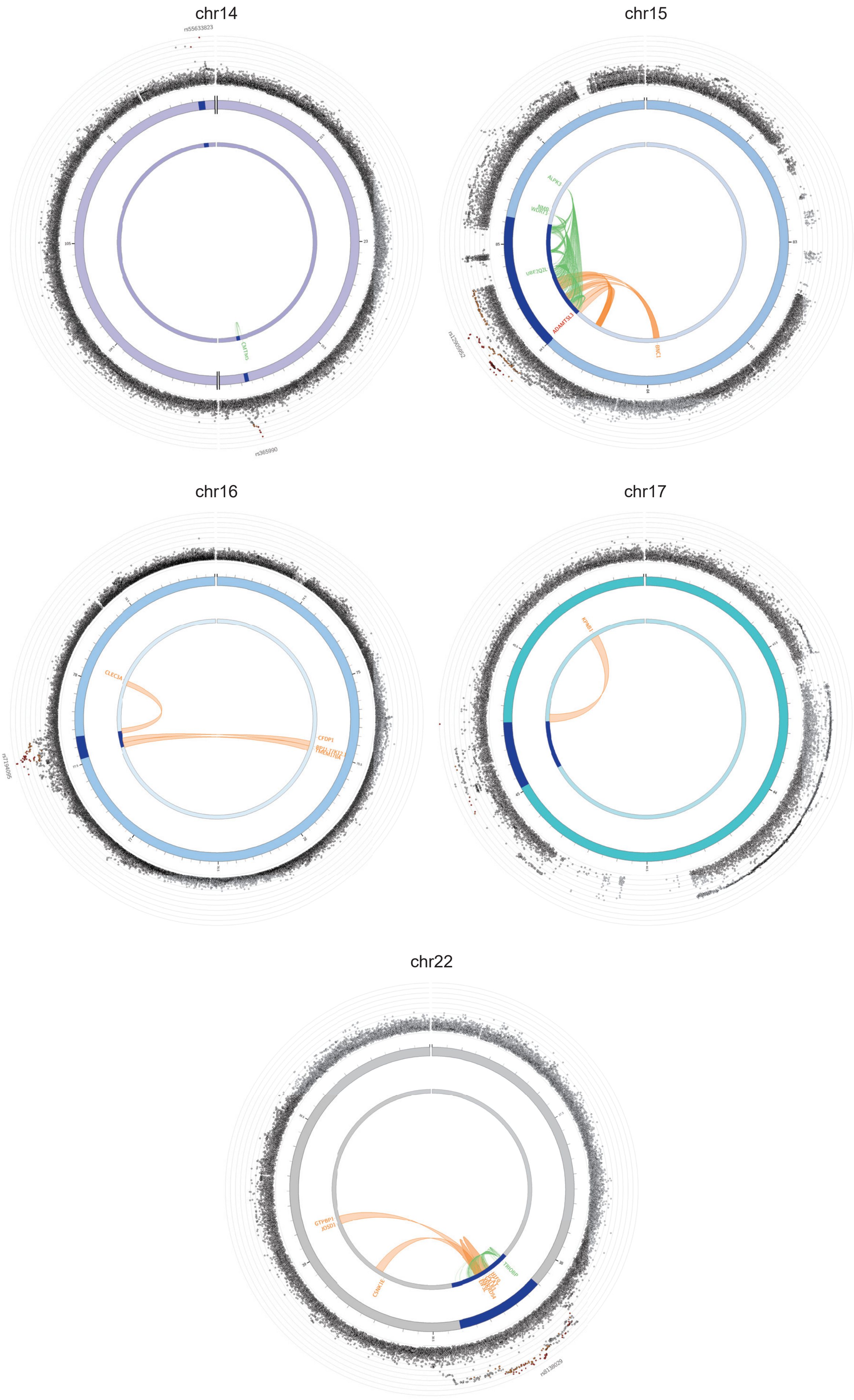
chr14
rs55633823
CMTM5
rs365990
chr15
ALPK3
NMB
WDR73
UBE2Q2L
ADAMTSL3
BNC1
chr16
CLEC3A
CFDP1
chr17
KPNB1
chr22
GTPBP1
JOSD1
CSNK1E
TRIOBP

**Supplementary Figure 5. Circos plots of genomic loci and functional mapping for latent motion traits.** The layers are defined from the outside in: **Outer layer:** Manhattan plot of variants with $P < 0.05$. SNPs are coloured based on linkage disequilibrium ($r^2$) with the independent significant lead variant: red ($r^2 > 0.8$), orange ($r^2 > 0.6$), green ($r^2 > 0.4$), blue ($r^2 > 0.2$), and grey ($r^2 \leq 0.2$). The y-axis scale ranges from 0 to the maximum $-\log_{10}(P)$ of the variants. Lead variants are labelled with rsIDs. **Second layer:** Chromosome ring with genomic risk loci highlighted in blue. **Inner ring:** Mapped effector genes. Genes implicated solely by chromatin interactions are coloured orange, those solely by eQTLs are coloured green, and genes supported by both evidence streams are coloured red (e.g., *MAST2* on chr1). **Centre:** Functional connections. Orange arcs represent chromatin interactions (derived from Left Ventricle Hi-C data; GSE87112), and green arcs represent eQTL associations (derived from GTEx v8 Heart Atrial Appendage and Heart Left Ventricle).

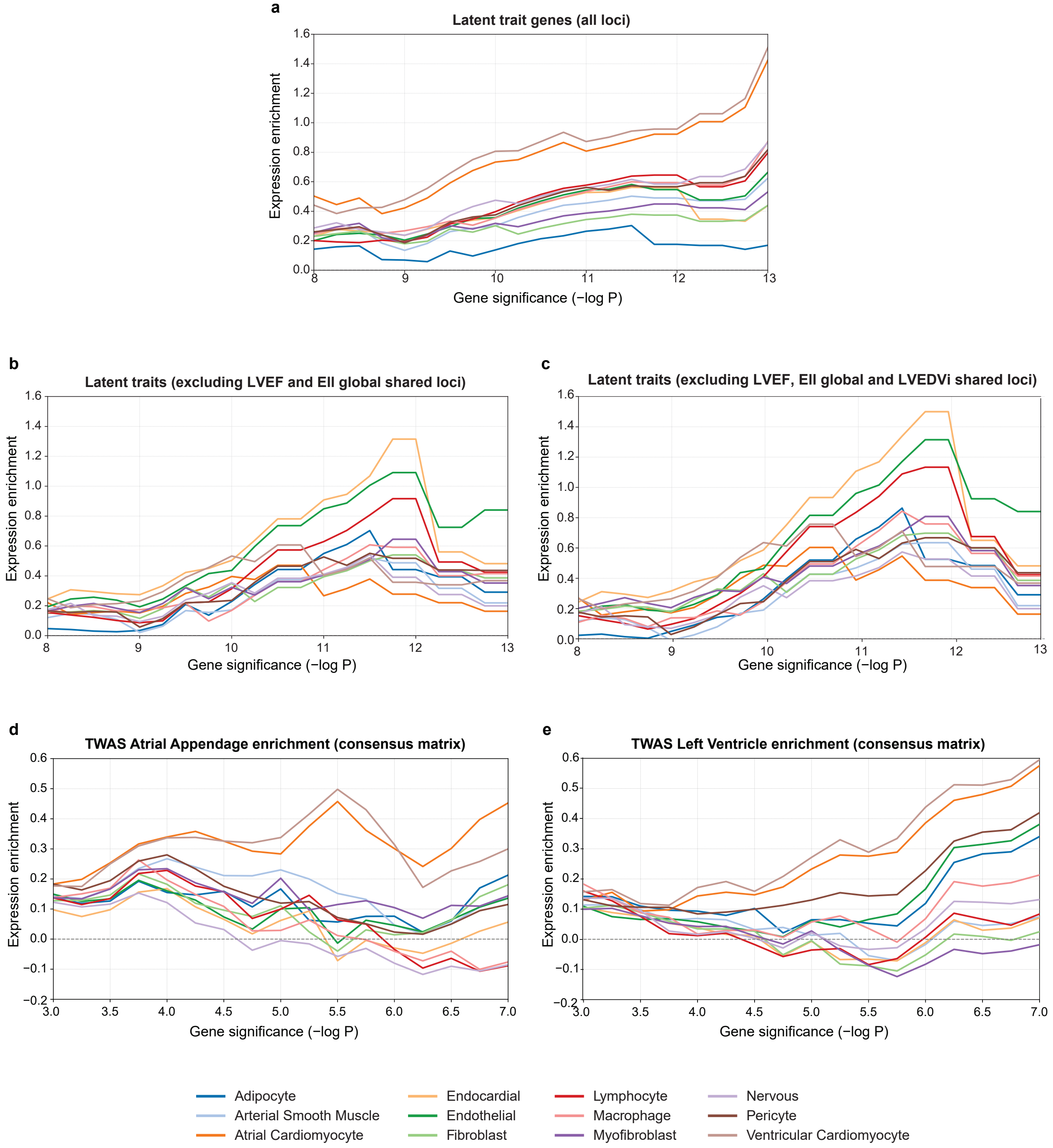


**Supplementary Figure 6. Sliding-threshold enrichment of latent motion trait genes across cardiac cell types.** As in Figure 4e, curves show the standardised mean difference in expression of GWAS-prioritised latent motion genes relative to all expressed genes in the corresponding cell type. **a**, Enrichment profile for genes at all loci associated with the latent left-ventricular motion traits, showing a dominant ventricular and atrial cardiomyocyte signal. **b**, The same sliding-threshold analysis after excluding loci shared with LVEF and Ell global, which modestly reduces the cardiomyocyte signal and relatively accentuates enrichment in several non-myocyte lineages, including fibroblast, endothelial and immune cells (as in Figure 4e). **c**, Further restriction to loci that are not shared with the addition of LVEDVi leads to a more pronounced effect of the non-myocyte enrichment. **d,** TWAS-derived gene sets from the atrial appendage (AA) show enrichment in atrial cardiomyocytes at higher significance thresholds, with endocardial cells emerging as a secondary signal. At lower thresholds, enrichment is broadly distributed across cell types. **e,** TWAS-derived gene sets from the left ventricle (LV) retain a positive ventricular cardiomyocyte signature across thresholds, consistent with the LV origin of the motion phenotype. Endocardial enrichment is also evident at intermediate thresholds, while adipocyte and lymphocyte signals remain near or below zero throughout.

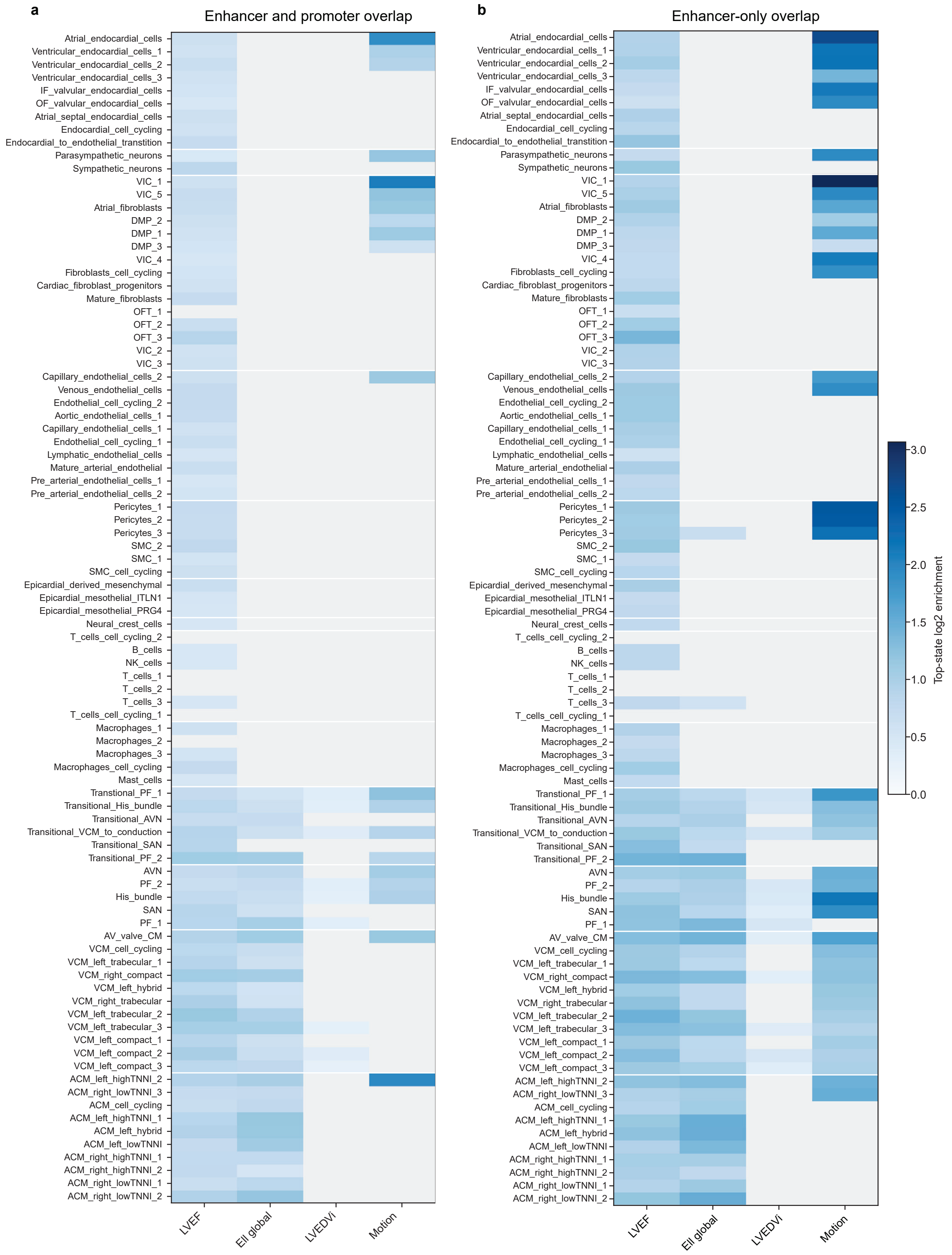


**Supplementary Figure 7. Full 90-state fine-mapped enrichment heatmaps. a,** Full 90-state all-regulatory heatmap. State-level enrichment derived from the all-regulatory analysis (enhancers plus promoters). Only states with $q < 0.05$ are coloured. The Motion column summarises the combined motion phenotype, and colour denotes representative state-level $\log_2$ enrichment. **b,** Full 90-state enhancer-only heatmap. The same visualisation generated from the all-regulatory analysis. This panel shows the enriched cell type specificity in an enhancer-only context and is presented relative to the more general promoter-inclusive view as a sensitivity analysis.

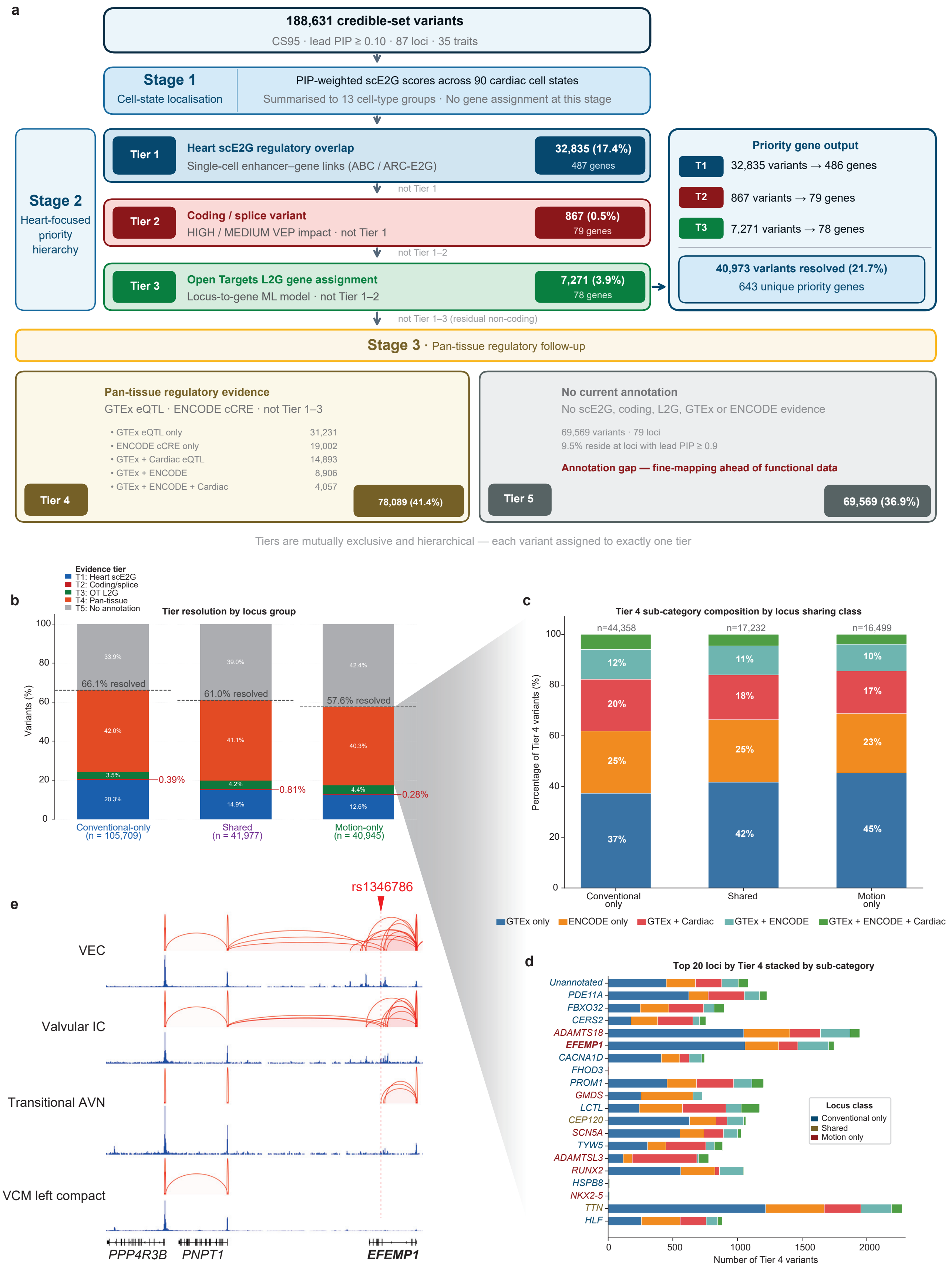
a
188,631 credible-set variants
CS95 · lead PIP ≥ 0.10 · 87 loci · 35 traits
Stage 1
Cell-state localisation
PIP-weighted scE2G scores across 90 cardiac cell states
Summarised to 13 cell-type groups · No gene assignment at this stage
Stage 2
Heart-focused priority hierarchy
Tier 1
Heart scE2G regulatory overlap
Single-cell enhancer–gene links (ABC / ARC-E2G)
32,835 (17.4%)
487 genes
not Tier 1
Tier 2
Coding / splice variant
HIGH / MEDIUM VEP impact · not Tier 1
867 (0.5%)
79 genes
not Tier 1–2
Tier 3
Open Targets L2G gene assignment
Locus-to-gene ML model · not Tier 1–2
7,271 (3.9%)
78 genes
Priority gene output
T1 32,835 variants → 486 genes
T2 867 variants → 79 genes
T3 7,271 variants → 78 genes
40,973 variants resolved (21.7%)
643 unique priority genes
not Tier 1–3 (residual non-coding)
Stage 3 · Pan-tissue regulatory follow-up
Pan-tissue regulatory evidence
GTEx eQTL · ENCODE cCRE · not Tier 1–3
• GTEx eQTL only 31,231
• ENCODE cCRE only 19,002
• GTEx + Cardiac eQTL 14,893
• GTEx + ENCODE 8,906
• GTEx + ENCODE + Cardiac 4,057
Tier 4
78,089 (41.4%)
No current annotation
No scE2G, coding, L2G, GTEx or ENCODE evidence
69,569 variants · 79 loci
9.5% reside at loci with lead PIP ≥ 0.9
Annotation gap — fine-mapping ahead of functional data
Tier 5
69,569 (36.9%)
Tiers are mutually exclusive and hierarchical — each variant assigned to exactly one tier
b
Evidence tier
T1: Heart scE2G
T2: Coding/splice
T3: OT L2G
T4: Pan-tissue
T5: No annotation
Tier resolution by locus group
Variants (%)
33.9%
39.0%
42.4%
66.1% resolved
61.0% resolved
57.6% resolved
42.0%
41.1%
40.3%
3.5%
4.2%
4.4%
20.3%
14.9%
12.6%
0.39%
0.81%
0.28%
Conventional-only (n = 105,709)
Shared (n = 41,977)
Motion-only (n = 40,945)
c
Tier 4 sub-category composition by locus sharing class
Percentage of Tier 4 variants (%)
n=44,358
n=17,232
n=16,499
12%
11%
10%
20%
18%
17%
25%
25%
23%
37%
42%
45%
Conventional only
Shared
Motion only
GTEx only
ENCODE only
GTEx + Cardiac
GTEx + ENCODE
GTEx + ENCODE + Cardiac
d
Top 20 loci by Tier 4 stacked by sub-category
Unannotated
PDE11A
FBXO32
CERS2
ADAMTS18
EFEMP1
CACNA1D
FHOD3
PROM1
GMDS
LCTL
CEP120
SCN5A
TYW5
ADAMTSL3
RUNX2
HSPB8
NKX2-5
TTN
HLF
Locus class
Conventional only
Shared
Motion only
Number of Tier 4 variants
e
rs1346786
VEC
Valvular IC
Transitional AVN
VCM left compact
PPP4R3B
PNPT1
EFEMP1

**Supplementary Figure 8. Hierarchical variant prioritisation framework and pan-tissue regulatory rescue. a,** Three-stage pipeline applied to 188,631 credible-set variants across 87 loci and 35 traits. Stage 1, PIP-weighted scE2G scoring across 90 cardiac cell states. Stage 2, mutually exclusive hierarchical assignment to Tier 1 (heart scE2G regulatory overlap), Tier 2 (coding/splice; VEP HIGH/MODERATE), or Tier 3 (Open Targets L2G), resolving 40,973 variants (21.7%) to 643 priority genes. Stage 3, pan-tissue regulatory rescue via GTEx eQTL or ENCODE cCRE overlap (Tier 4; 78,089 variants, 41.4%); Tier 5, unannotated. **b,** Tier composition across locus-sharing classes. Motion-only loci carry the lowest Tier 1 fraction (12.6% vs. 20.3% for conventional-only) and the largest unannotated tail (42.4%), consistent with the relative novelty of motion-trait associations. **c,** Tier 4 sub-category composition by locus-sharing class. GTEx-only variants predominate across all groups; multi-evidence variants (GTEx + ENCODE + cardiac eQTL) account for 10–12%. **d,** Top 20 loci ranked by Tier 4 variant count, stacked by sub-category. Motion-only loci (*ADAMTS18, EFEMP1, GMDS, SCN5A*) dominate the high-PIP end; an unannotated chr19 locus (PIP = 0.99) and *PDE11A* (PIP = 0.79) lack heart-specific gene assignment and represent priority candidates for targeted cardiac regulome integration. **e,** Representative latent-only locus prioritised in both Tier 1 and Tier 4. scE2G predictions and normalised ATAC-seq signals are shown for a variant that falls within a predicted enhancer inside the *EFEMP1* gene body. Ventricular endocardial cells (VEC), valvular interstitial cells (Valvular IC), and transitional atrioventricular node (Transitional AVN) all link the enhancer to the *EFEMP1* promoter, whereas ventricular cardiomyocytes show no predicted enhancer activity at this position. In VEC and Valvular IC, the same regulatory neighbourhood also connects to the *PNPT1* promoter, which shows promoter–promoter connectivity to *PPP4R3B*.

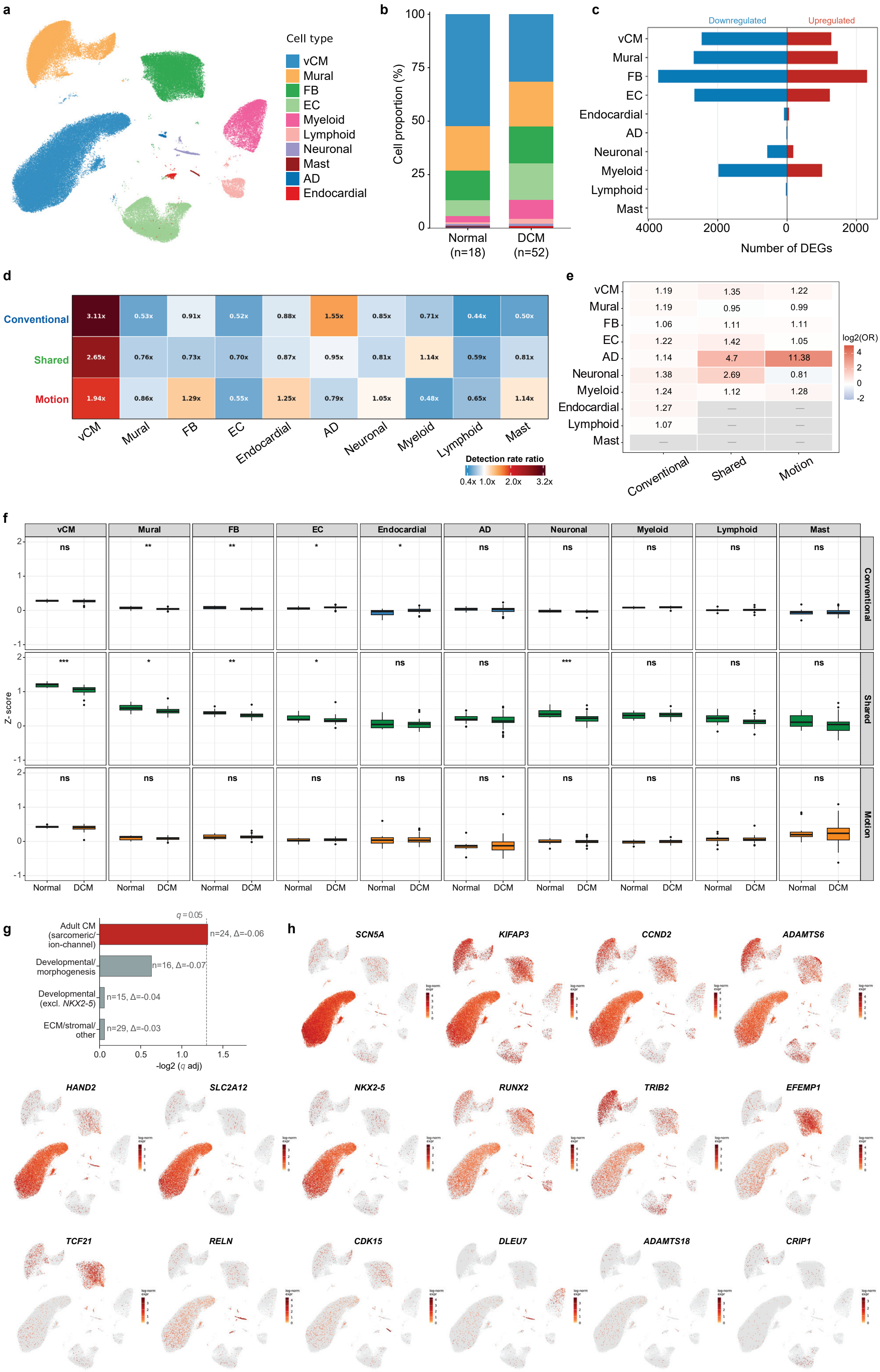
a
Cell type
vCM
Mural
FB
EC
Myeloid
Lymphoid
Neuronal
Mast
AD
Endocardial
b
Cell proportion (%)
Normal (n=18)
DCM (n=52)
c
Downregulated
Upregulated
Number of DEGs
d
Conventional
Shared
Motion
Detection rate ratio
e
log2(OR)
Conventional
Shared
Motion
f
Z-score
Normal
DCM
g
Adult CM (sarcomeric/ion-channel)
Developmental/morphogenesis
Developmental (excl. NKX2-5)
ECM/stromal/other
-log2 (q adj)
h
SCN5A
KIFAP3
CCND2
ADAMTS6
HAND2
SLC2A12
NKX2-5
RUNX2
TRIB2
EFEMP1
TCF21
RELN
CDK15
DLEU7
ADAMTS18
CRIP1

**Supplementary Figure 9. Single-nucleus RNA-seq validation of prioritised motion, shared, and conventional gene sets in normal and dilated cardiomyopathy left ventricle.** Analysis of 612,457 left ventricular nuclei from 78 donors (Normal, $n = 18$; DCM, $n = 52$; ARVC, $n = 8$) from Reichart et al. 2022[18]. **a**, UMAP embedding of 100,000 randomly downsampled nuclei coloured by annotated cell type. **b**, Cell-type composition in Normal and DCM hearts. **c**, Number of differentially expressed genes (DEGs) per cell type in DCM versus Normal ($p_{\mathrm{adj}} < 0.05$, $|\log_2 \mathrm{FC}| > 0.25$; pseudobulk DESeq2). **d**, Cell-type preference of each gene set in Normal hearts, computed as the detection rate ratio (observed / mean across all cell types; $1.0\times$ = no preference). **e**, Enrichment of each gene set among cell-type DEGs in DCM versus Normal (Fisher's exact test, Benjamini−Hochberg corrected across 30 tests). No test survived correction (best $p_{\mathrm{adj}} = 0.17$, Neuronal/Shared); Odds ratios shown for reference. Dashes indicate insufficient DEGs for testing. **f**, Per-donor Z-score of each gene set (rows) across all ten cell types (columns) in Normal (grey) and DCM (coloured) donors. Z-scores computed per donor as the mean normalised expression of genes within each set, standardised across all genes. Boxes show interquartile range across donors; median line and individual donor points overlaid. Significance reflects pairwise Wilcoxon rank-sum tests with Benjamini−Hochberg correction across 30 comparisons. $^{***}\ p_{\mathrm{adj}} < 0.001$, $^{**}\ p_{\mathrm{adj}} < 0.01$, $^{*}\ p_{\mathrm{adj}} < 0.05$, ns = not significant. **g**, Within-set dissociation of the Motion gene set in DCM ventricular cardiomyocytes. Bars show $-\log_{10}(q_{\mathrm{adj}})$ for Wilcoxon rank-sum tests of per-donor Z-scores (DCM vs. Normal) for four functional subcategories. The adult cardiomyocyte subset is significantly downregulated in DCM ($q_{\mathrm{adj}} = 0.048$, red), whereas the developmental subset shows no coordinated change and becomes definitively null once *NKX2-5* is excluded. Annotations indicate subcategory size ($n$) and median $\Delta$ Z-score. Dashed line, $q = 0.05$. **h**, UMAP feature plots of selected Motion genes (from Tiers 1 and 2), ordered by percentage of expressing cells in Normal donors. Colour scale represents log-normalised expression; grey indicates non-detection.

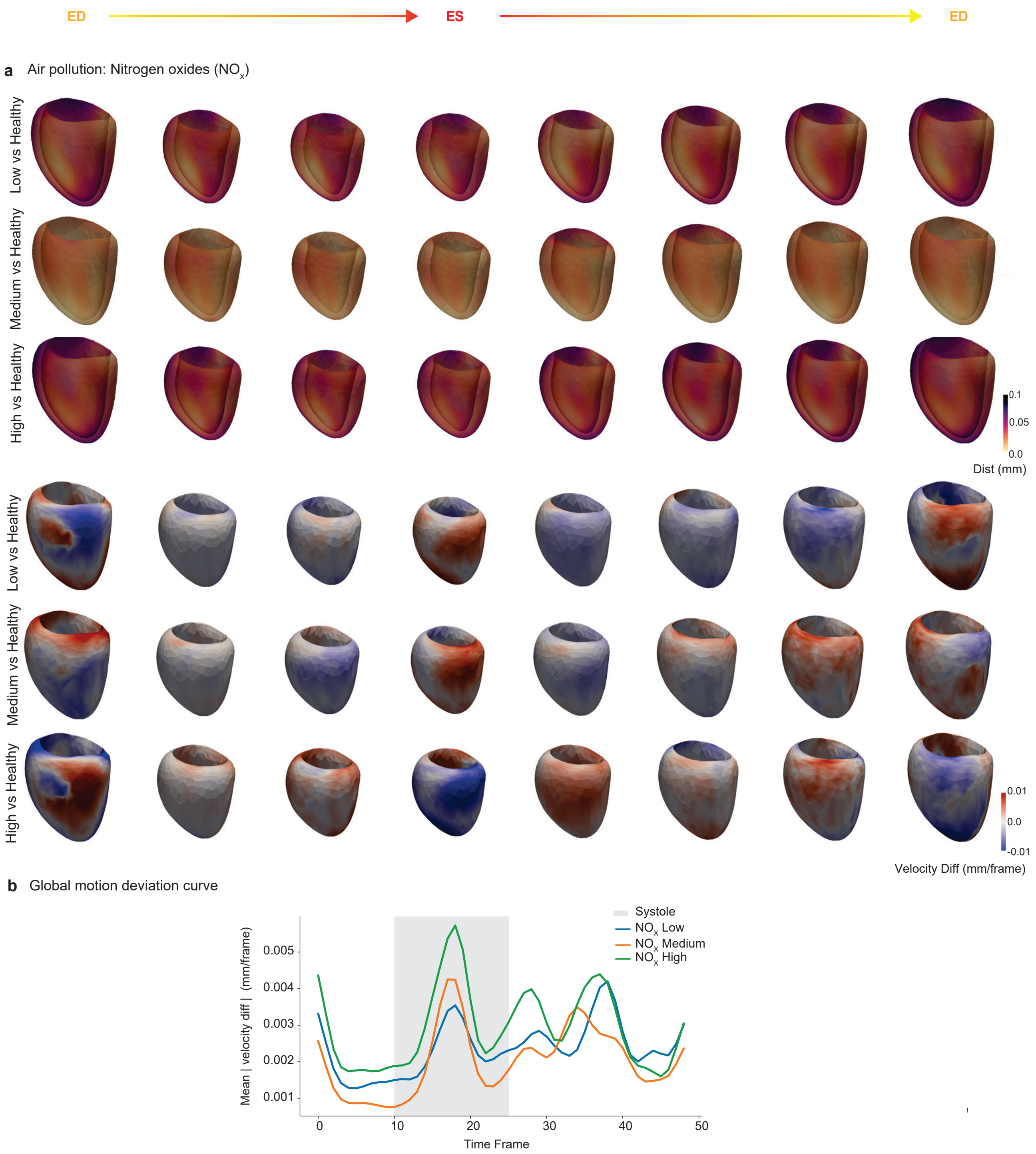


**Supplementary Figure 10. Spatiotemporal differences in left ventricular motion and deformation ccross environmental exposure to Nitrogen Oxides**.

**a**, Local displacement magnitude maps (Dist in mm) showing pointwise deviations in average left ventricular (LV) geometry relative to the control group across the cardiac cycle. Warmer colours indicate regions of greater displacement, reflecting more pronounced deformation relative to controls. Corresponding maps of local myocardial velocity differences (Velocity Diff in mm/frame) between exposure groups and controls. Red regions indicate faster motion and blue regions indicate slower motion compared with the reference. **b**, Global motion deviation curves showing the average magnitude of verte-xwise velocity differences (mm/frame) between each exposure group and the control cohort.

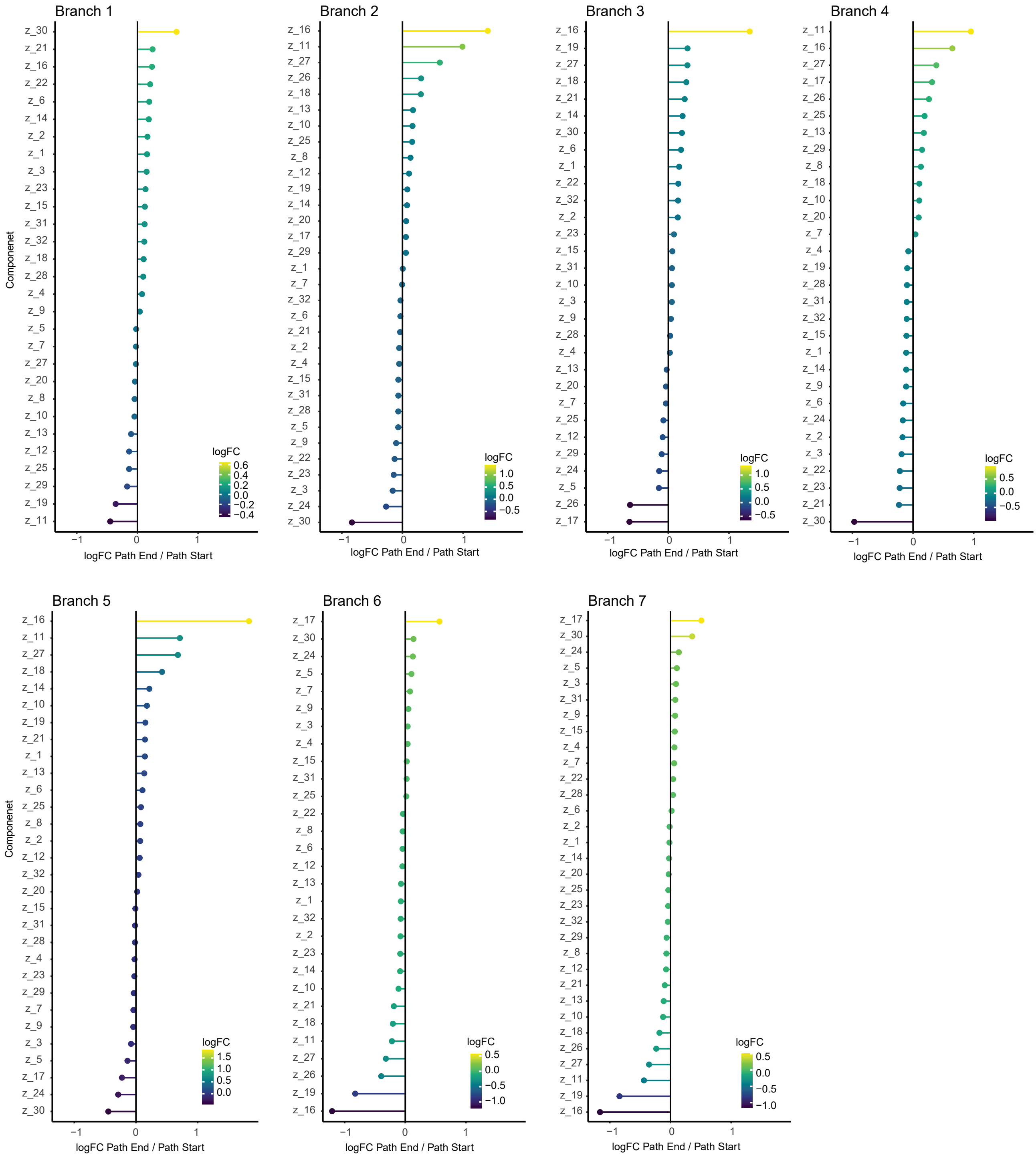


**Supplementary Figure 11. Latent motion feature changes along lineage pseudotime.** Differential expression of the 32-dimensional latent cardiac motion features $z$ was assessed between the start and end points of each lineage. Variables showing a statistically significant log fold change (LogFC) along pseudotime across the seven branches are plotted.

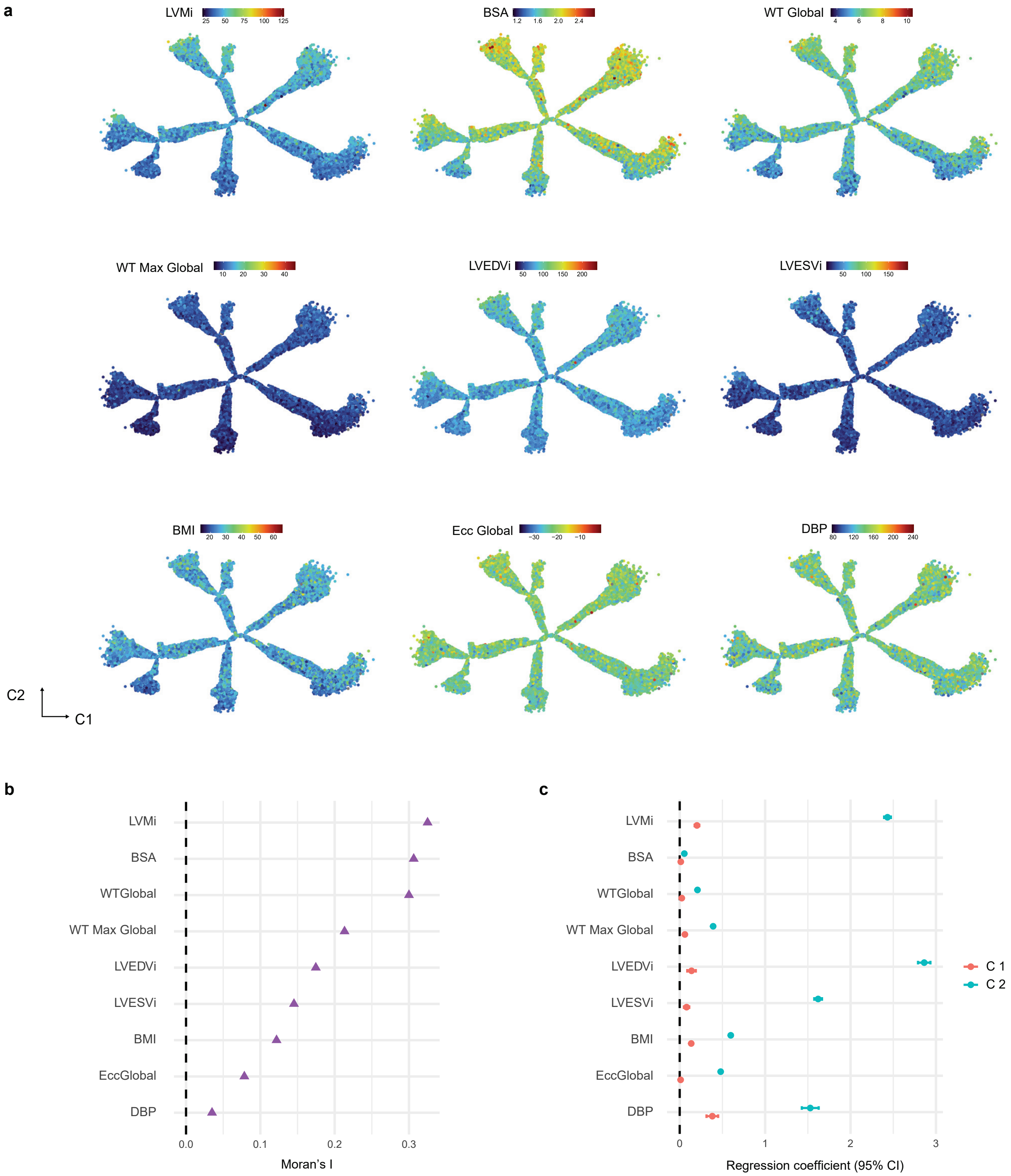


**Supplementary Figure 12. Tree-Structured spatial patterns in motion features. a**. Phenotypes overlaid on the motion tree structure, showing spatial gradients. **b**. Moran's I values quantifying spatial clustering strength. **c**. Regression coefficients showing directional loadings on tree axes. BMI, body mass index; BSA, body surface area; DBP, diastolic blood pressure; Ecc Global, global circumferential strain; LVEDVi, left ventricular end-diastolic volume indexed; LVESVi, left ventricular end-systolic volume indexed; LVMi, left ventricular mass indexed; WT Global, global wall thickness; WT Max Global, global maximum wall thickness.

**a**

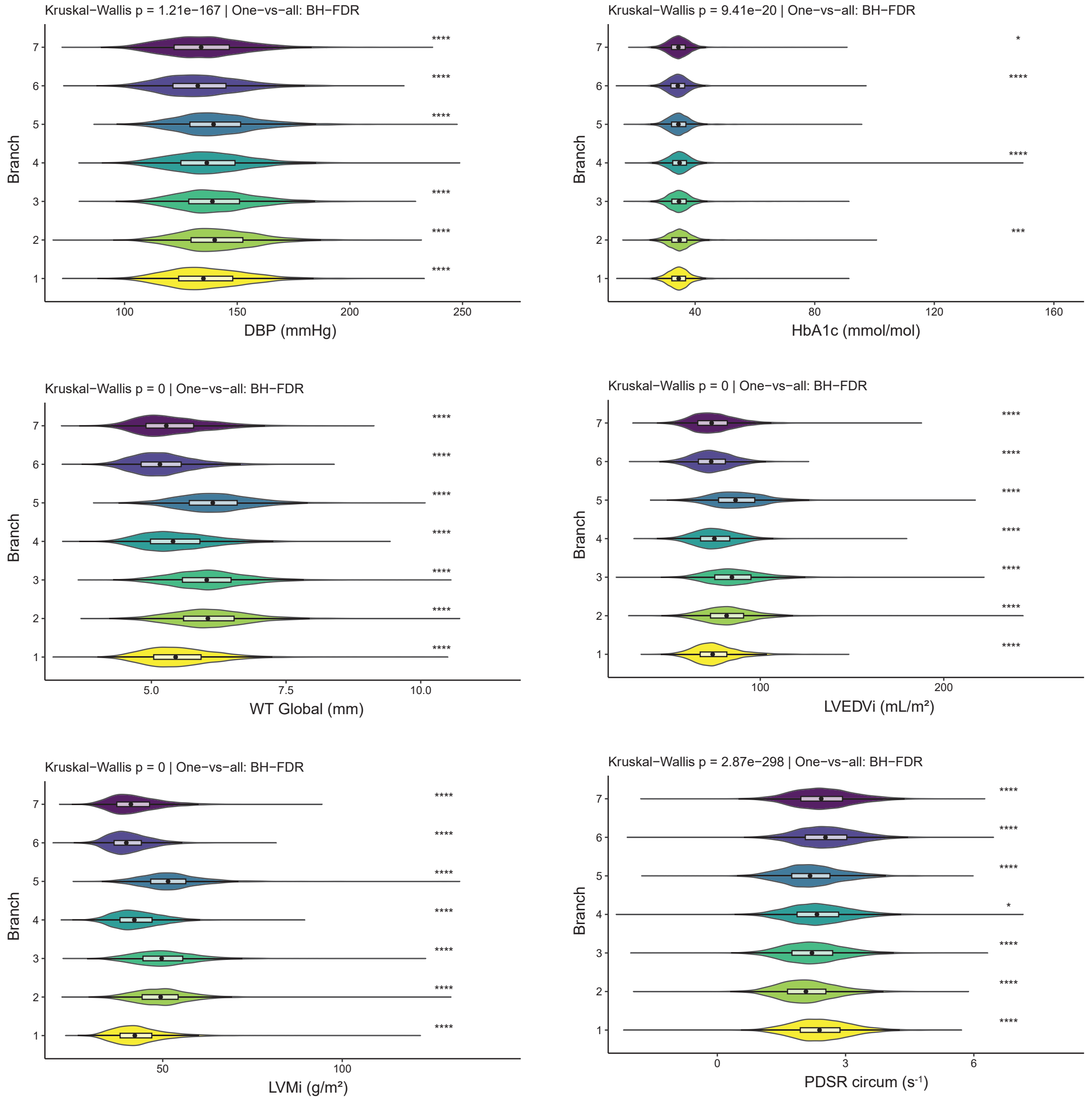


**b**

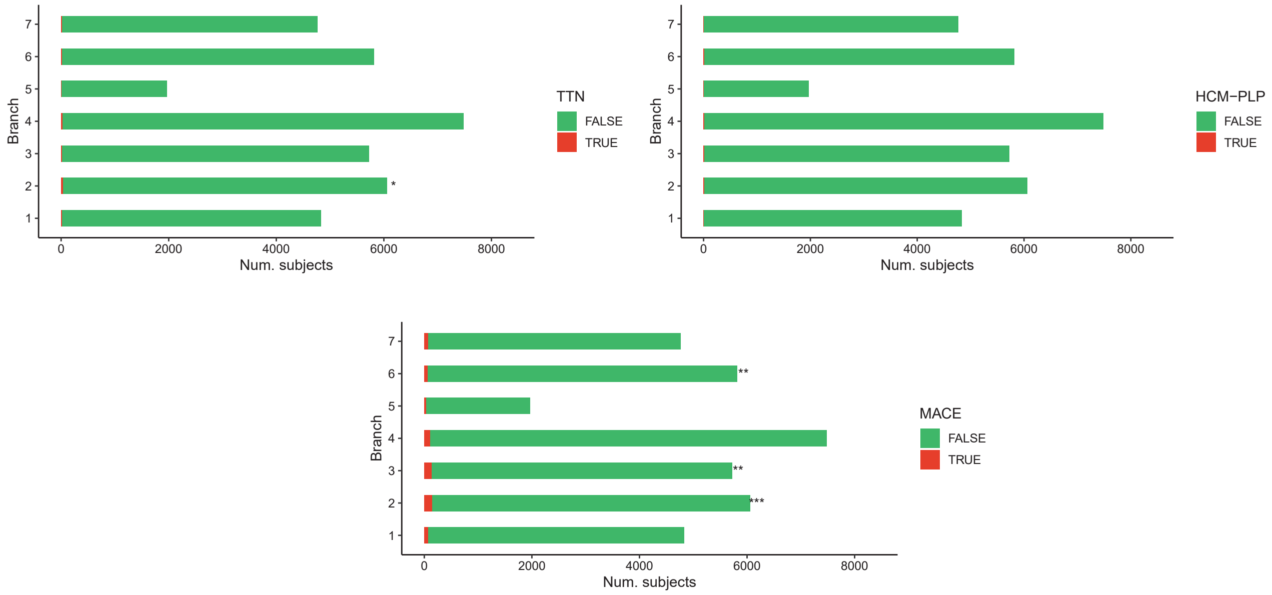


**Supplementary Figure 13. Branch-level enrichment analysis comparing each data-driven cardiac morphology branch with all other participants. a**. Continuous traits were compared using Wilcoxon rank-sum tests; violin plots show distribution per branch with median points and FDR-adjusted significance (Benjamini−Hochberg adjusted *p*). **b**. Binary traits (e.g., genotype carrier status, MACE occurrence) were compared using Fisher's exact tests; bar heights indicate prevalence within each branch and stars denote branches with significant enrichment after FDR correction. Asterisks indicate FDR-adjusted significance levels: $^{*}P\leq0.05$, $^{**}P\leq10^{-3}$, $^{***}P\leq10^{-4}$, $^{****}P\leq10^{-6}$. BH, Benjamini Hochberg, DBP, diastolic blood pressure; FDR: false-discovery rate; HbA1c, glycated haemoglobin; HCM, hypertrophic cardiomyopathy; LVEDVi, left ventricular end-diastolic volume indexed; LVMi, left ventricular mass indexed; MACE, major adverse cardiac event; PDSR circum, circumferential peak diastolic strain rate; WT Global, global wall thickness.

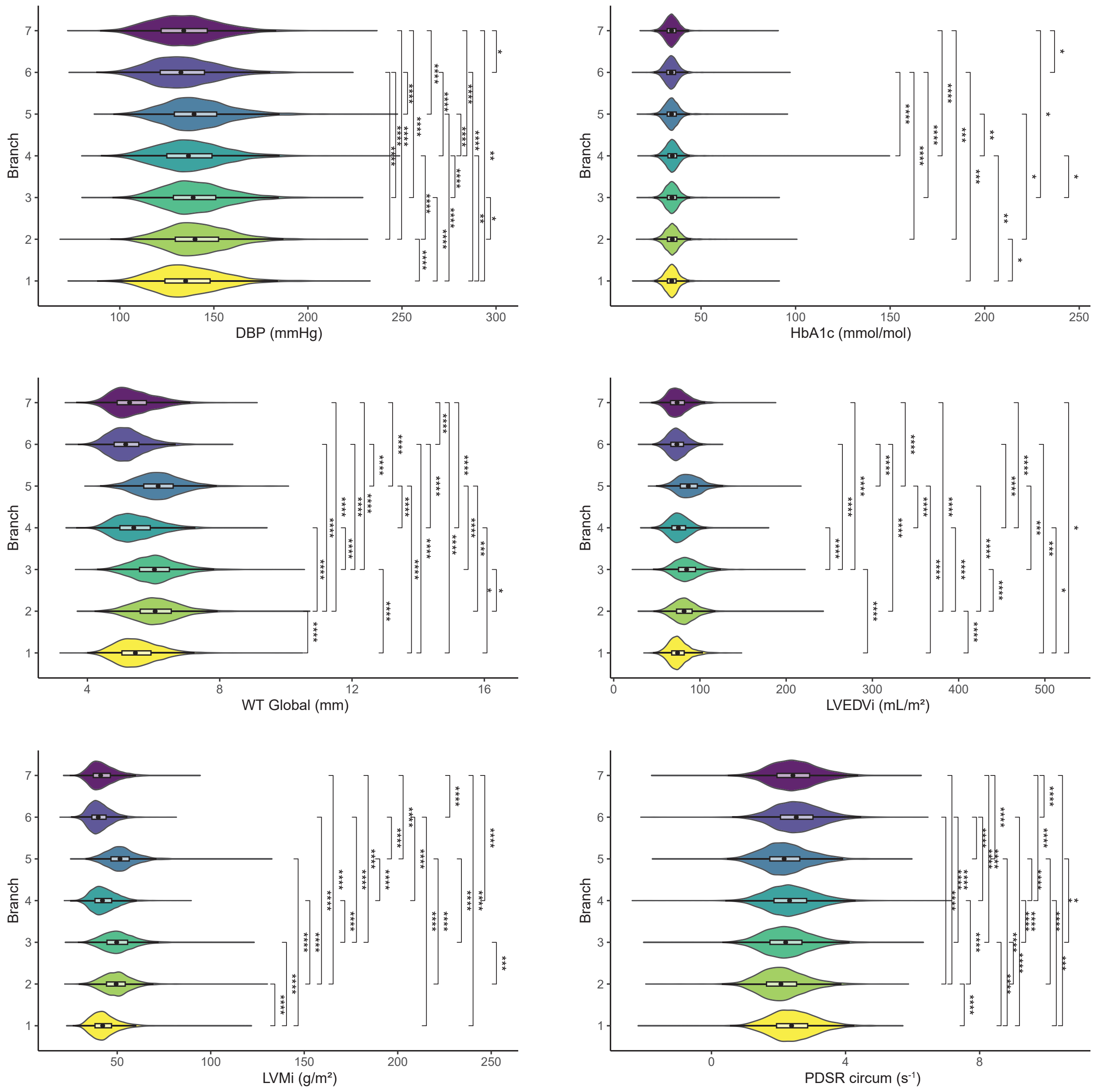


**Supplementary Figure 14. Pairwise branch comparison of continuous phenotypes.** Violin plots show the distribution of each phenotype across branches with median markers. Pairwise Wilcoxon rank-sum tests were applied following a global Kruskal–Wallis test; significance brackets indicate FDR-adjusted p-values. Asterisks denote FDR-adjusted significance levels: $^{*}P \leq 0.05$, $^{**}P \leq 10^{-3}$, $^{***}P \leq 10^{-4}$, $^{****}P \leq 10^{-6}$. DBP, diastolic blood pressure; FDR: false-discovery rate; HbA1c, glycated haemoglobin; HCM, hypertrophic cardiomyopathy; LVEDVi, left ventricular end-diastolic volume indexed; LVMi, left ventricular mass indexed; MACE, major adverse cardiac event; PDSR circum, circumferential peak diastolic strain rate; WT Global, global wall thickness.

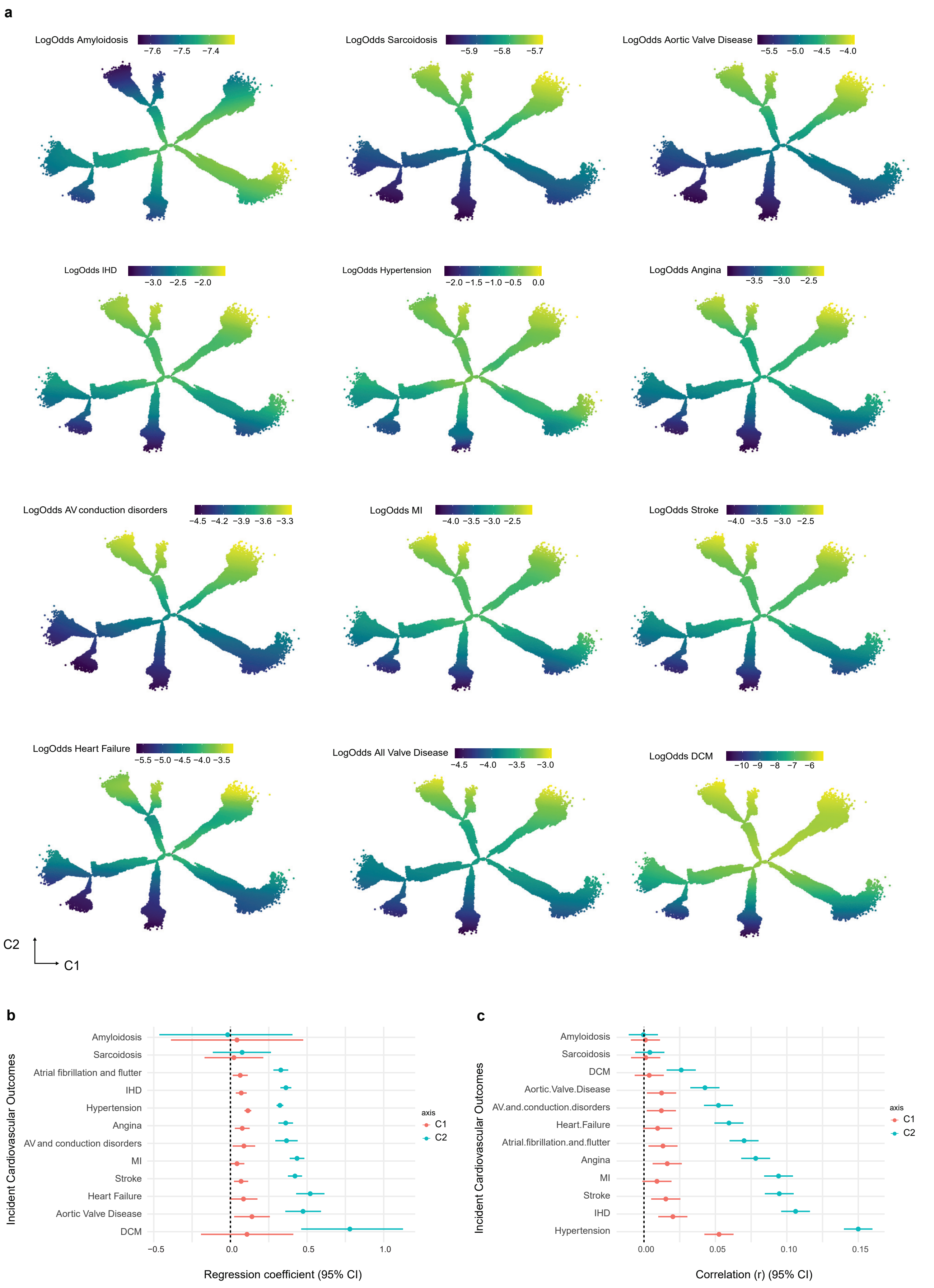


**Supplementary Figure 15. Mapping cardiovascular outcome variation across the DDRTree latent space a**. Logistic generalised additive model (GAM) fitted on DDRTree coordinates ($c_1$, $c_2$) showing spatial variation in cardiac outcome risk. **b,c**. Associations between standardised DDRTree axes and incident cardiovascular outcomes, assessed using univariate logistic regression (**b**) and point−biserial correlation (**c**). Forest plots display regression coefficients and correlation estimates (95% CI), illustrating how variation along DDRTree axes captures clinically relevant gradients in cardiovascular disease risk. Cardiovascular outcomes in the cohort are summarised in Supplementary Table 11. AV, atrioventricular; DCM, dilated cardiomyopathy; IHD, ischaemic heart disease; MI, myocardial infarction.

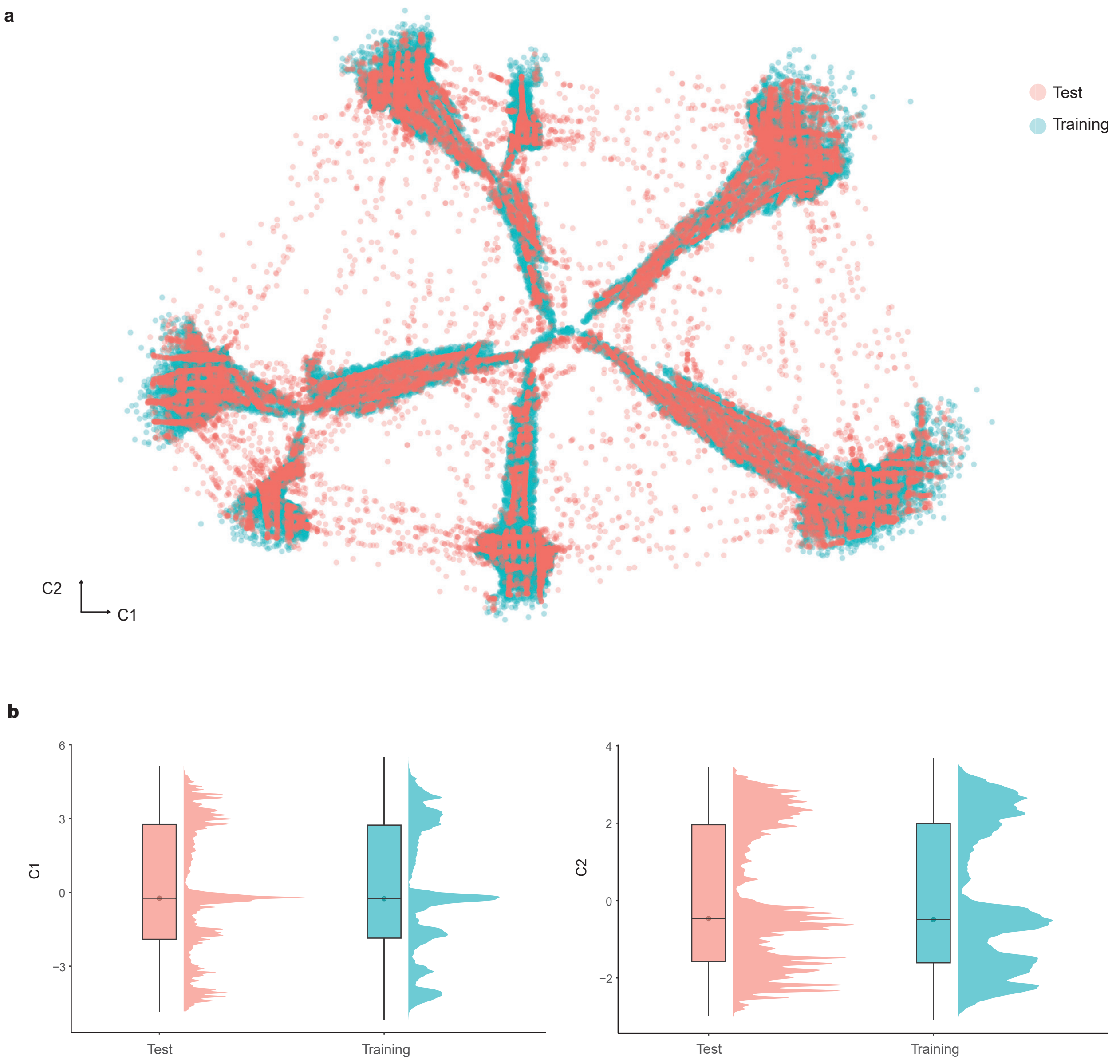


**Supplementary Figure 16. Generalisation of the DDRTree embedding across participants. a**. Using the trained DDRTree model, the $41,257$ test participants were projected onto the manifold learned from $36,856$ training individuals. **b**. Raincloud plots showing the distribution of training and test participants along the two tree axes $(c_1, c_2)$ demonstrates that the low-dimensional manifold of 4D cardiac motion traits is stable and reproducible across unseen UK Biobank participants; quantitative comparison is reported in Table 8.

**a**

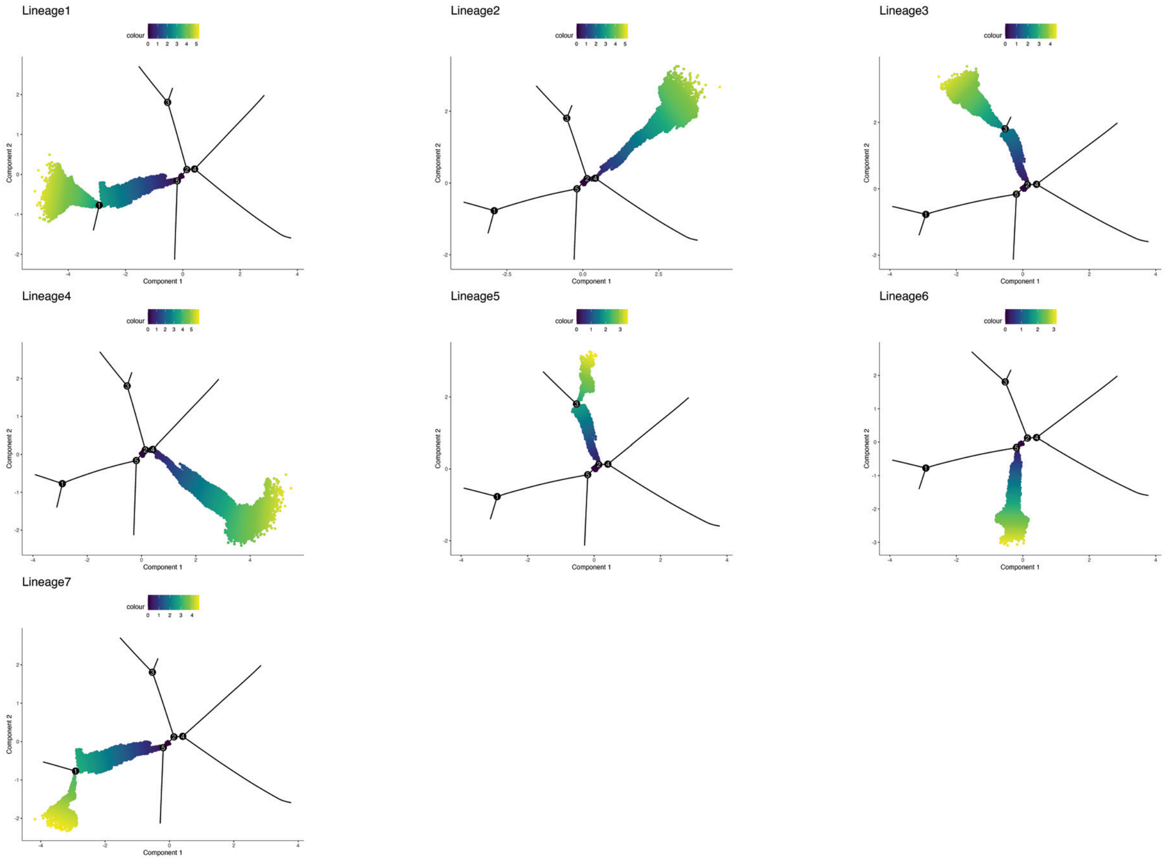


**b**

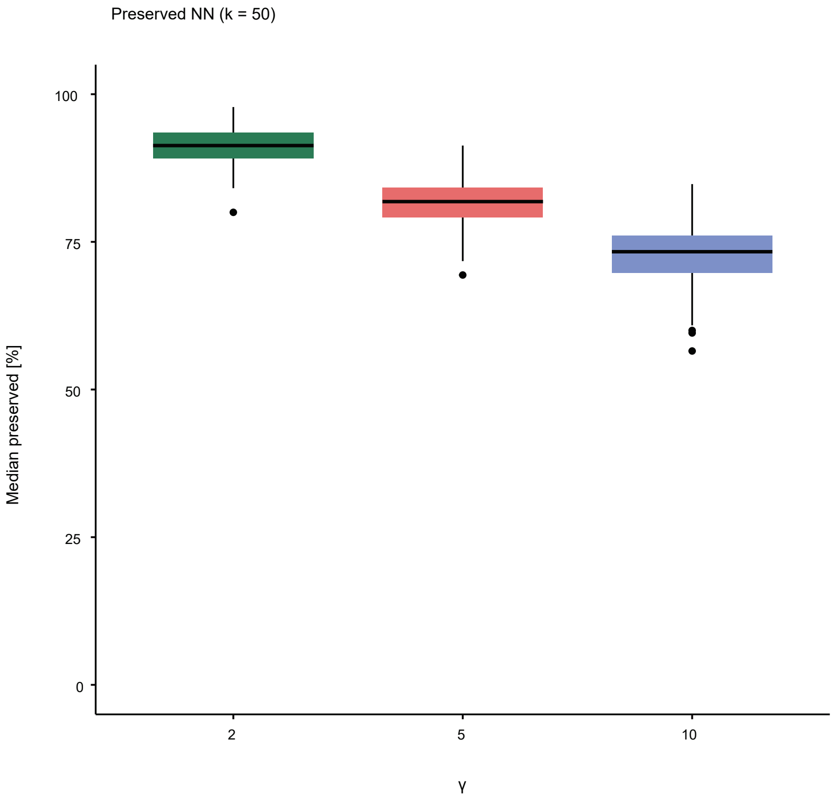


**Supplementary Figure 17. Lineage structure and model stability.a**. Lineages for the seven identified branches are shown, beginning at the root node, which represents the subject with the most average phenotype in the model, and extending outward to the periphery. Each lineage is colored according to pseudotime, which indicates how far each subject is positioned along the branch relative to the root node. **b**. Stability analysis using different model parameters of $\gamma(2, 5, 10)$ to estimate the median percentage of nearest neighbours (NN%) over $500$ replicates.

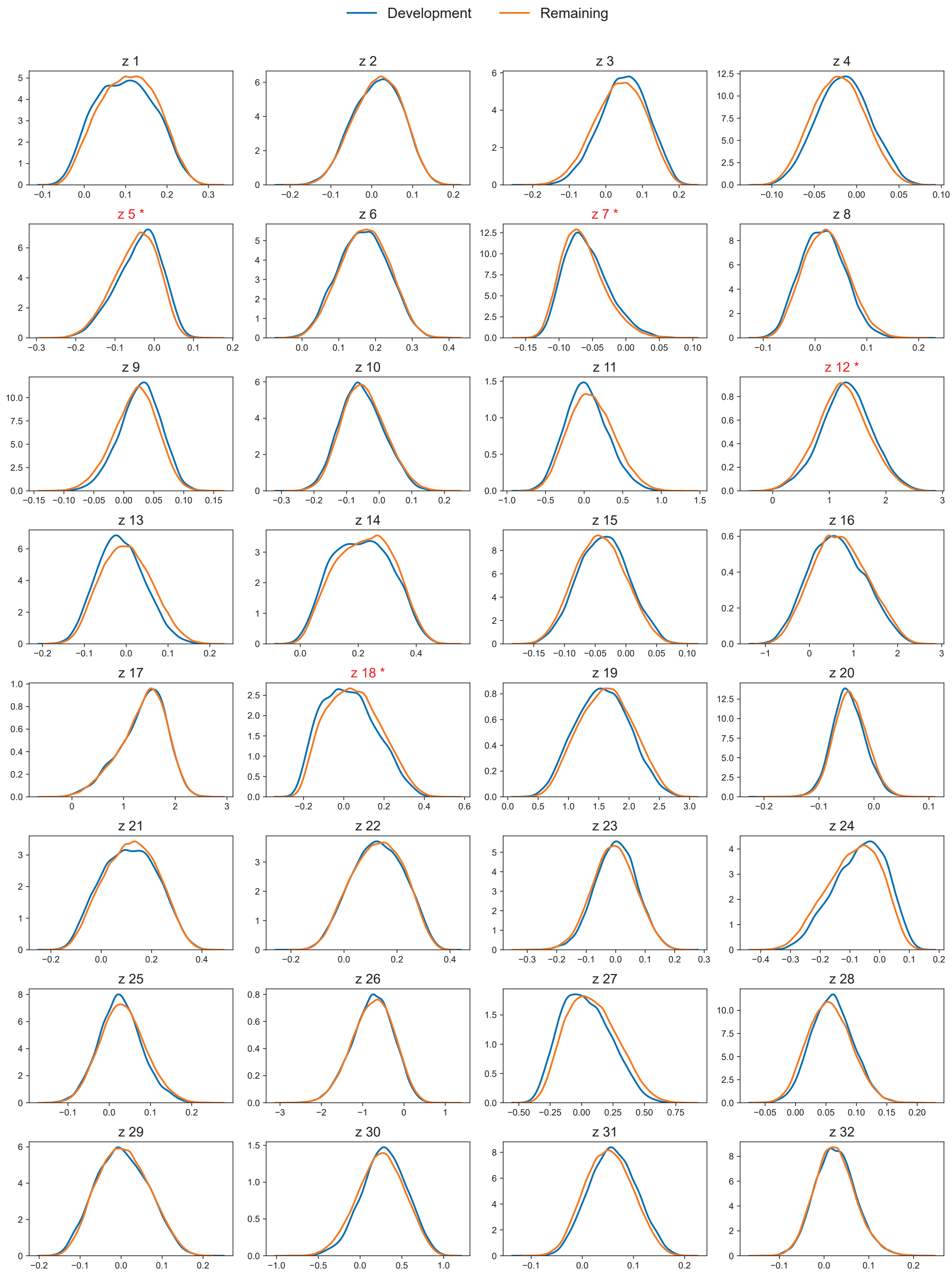


**Supplementary Figure 18.** Distribution of the 32 latent variables in the model-development cohort (blue) and remaining UK Biobank participants (orange). Red stars indicate the four latent dimensions with the highest proportion of participants outside the range observed during model development (see Supplementary Table 12). Even for these latent dimensions, the distributions remained highly consistent between cohorts.

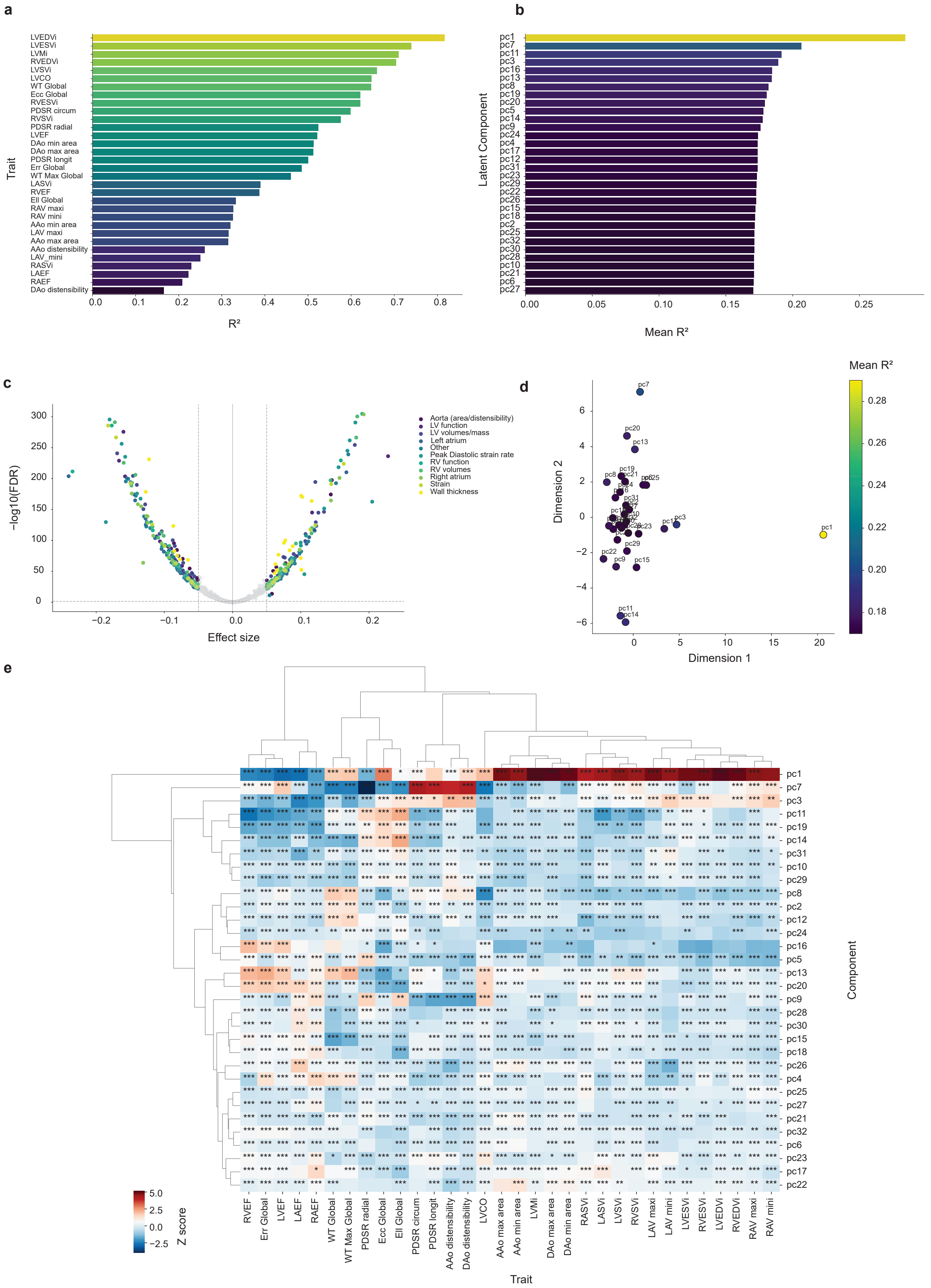

**Supplementary Figure 19. Predictive power and trait associations of PCA-derived motion components**. Panel **a** is based on the multivariable model including all 32 principle components, whereas panels **b–e** are based on analyses of individual principle components. **a**, Imaging trait predictability from motion-derived principle components, presented by variance explained ($R^2$). **b**, Mean variance explained ($R^2$) by each component across traits, each bar represents the mean variance explained by a single compoent across all CMR traits. **c**, Volcano plot of component–trait associations colored by trait category, revealing numerous robust associations across cardiac and vascular traits. **d**, Two-dimensional projection of standardised effect-size profiles across traits, showing similarity relationships among the 32 principal components, coloured by mean $R^2$. **e**, Hierarchically clustered statistical heatmap of covariate-adjusted component–trait associations (Euclidean distance; Ward linkage); Asterisks denote FDR-significant associations (*FDR $\leq$ 0.05, **FDR $\leq$ 0.01, ***FDR $\leq$ 0.001).

### Supplemental Tables

| Trait | Mean ± SD | Units |
|---|---|---|
| White ethnicity | 96.41 | % |
| Female sex | 52.93 | % |
| Male sex | 47.07 | % |
| Age at MRI | 66.22 ± 8.01 | years |
| BSA | 1.87 ± 0.21 | $m^2$ |
| DBP | 78.87 ± 10.16 | mmHg |
| SBP | 141.96 ± 19.57 | mmHg |
| *Left ventricular deformation and strain* | | |
| WT Global | 5.69 ± 0.77 | mm |
| WT Max Global | 9.50 ± 1.75 | mm |
| Ecc Global | −22.35 ± 3.59 | % |
| Err Global | 45.73 ± 8.78 | % |
| Ell Global | −18.50 ± 2.95 | % |
| PDSR longitudinal | 1.62 ± 0.58 | $s^{-1}$ |
| PDSR radial | −5.75 ± 1.87 | $s^{-1}$ |
| PDSR circumferential | 2.26 ± 0.75 | $s^{-1}$ |
| *Ventricular volumes and mass* | | |
| LVEDVi | 77.53 ± 14.33 | $mL/m^2$ |
| LVESVi | 31.52 ± 8.98 | $mL/m^2$ |
| LVMi | 45.04 ± 8.45 | $g/m^2$ |
| RVEDVi | 82.25 ± 15.46 | $mL/m^2$ |
| RVESVi | 35.37 ± 9.43 | $mL/m^2$ |
| RVSVi | 46.87 ± 9.12 | $mL/m^2$ |
| LVSVi | 46.01 ± 8.57 | $mL/m^2$ |

| Trait | Mean ± SD | Units |
|---|---|---|
| *Atrial volumes and function* | | |
| RAV min | 24.99 ± 10.10 | $mL/m^2$ |
| RAV max | 46.45 ± 14.25 | $mL/m^2$ |
| RASVi | 21.46 ± 6.92 | $mL/m^2$ |
| LAV min | 16.35 ± 8.84 | $mL/m^2$ |
| LAV max | 39.24 ± 12.00 | $mL/m^2$ |
| LASVi | 22.88 ± 5.72 | $mL/m^2$ |
| *Global cardiac function* | | |
| LVEF | 59.65 ± 6.44 | % |
| LVCO | 5.37 ± 1.24 | L/min |
| RVEF | 57.25 ± 6.50 | % |
| LAEF | 59.99 ± 9.76 | % |
| RAEF | 46.78 ± 9.11 | % |
| *Aortic dimensions and distensibility* | | |
| AAo max area | 865.16 ± 192.68 | $mm^2$ |
| AAo min area | 790.68 ± 187.12 | $mm^2$ |
| AAo distensibility | 1.65 ± 2.74 | $10^{-3}$ $mmHg^{-1}$ |
| DAo max area | 480.38 ± 99.51 | $mm^2$ |
| DAo min area | 426.46 ± 94.19 | $mm^2$ |
| DAo distensibility | 2.16 ± 1.53 | $10^{-3}$ $mmHg^{-1}$ |

**Supplementary Table 1. Participant characteristics.** Demographic and imaging-derived cardiac parameters for the study participants. Values are presented as mean ± SD (SD denotes standard deviation). BSA, body surface area; DBP, diastolic blood pressure; SBP, systolic blood pressure; AAo, ascending aorta; DAo, descending aorta; Ell, longitudinal strain; Ecc, circumferential strain; Err, radial strain; LAEF, left atrial ejection fraction; LASVi, left atrial stroke volume indexed; LAVmax, maximum left atrial volume indexed; LAVmin, minimum left atrial volume indexed; LVCO, left ventricular cardiac output; LVCi, left ventricular cardiac index; LVEDVi, left ventricular end-diastolic volume indexed; LVEF, left ventricular ejection fraction; LVESVi, left ventricular end-systolic volume indexed; LVMi, left ventricular mass indexed; LVSVi, left ventricular stroke volume indexed; PDSR, peak diastolic strain rate; RAEF, right atrial ejection fraction; RASVi, right atrial stroke volume indexed; RAVmax, maximum right atrial volume indexed; RAVmin, minimum right atrial volume indexed; RVEDVi, right ventricular end-diastolic volume indexed; RVEF, right ventricular ejection fraction; RVESVi, right ventricular end-systolic volume indexed; RVSVi, right ventricular stroke volume indexed; WT, myocardial wall thickness.

| | TTN | MYH7 | BAG3 | DCM P/LP | HCM P/LP |
|---|---|---|---|---|---|
| **Count** | 252 | 5 | 3 | 323 | 128 |

**Supplementary Table 2.** Summary of genotype frequencies and pathogenic or likely pathogenic (P/LP) variant classifications among 78,113 UK Biobank participants.

| **Exposure factor** | | **Count** |
|---|---|---|
| **Air Pollution ($\mu$g/m$^3$)** | | |
| | Low: $0.00 \leq NO_2 < 98.11$ | 29,140 |
| $NO_2$ (2005–2007 and 2010) | Medium: $98.11 \leq NO_2 < 125.32$ | 25,213 |
| | High: $125.32 \leq NO_2 \leq 500.50$ | 23,684 |
| | Low: $19.74 \leq NO_x < 37.36$ | 28,957 |
| $NO_x$ (2010) | Medium: $37.36 \leq NO_x < 47.58$ | 24,591 |
| | High: $47.58 \leq NO_x \leq 265.94$ | 23,489 |
| **Smoking Status** | | |
| | Former smokers | 25,804 |
| | Current smokers | 4,986 |

**Supplementary Table 3.** Air pollution exposure tertiles and smoking status among 78,113 UK Biobank participants. $NO_x$, Nitric oxides; $NO_2$, Nitric dioxide.

| Characteristic | Estimate ($\beta$) | 95% CI | p-value |
|---|---|---|---|
| (Intercept) | 137.69 | (137.52, 137.87) | < 2e−16 |
| $c_1$ | 0.58 | (-0.17, 1.34) | 0.1293 |
| $c_2$ | 5.79 | (-0.68, 12.26) | 0.0796 |
| $s(c_1)$ | — | — | 0.449 |
| $s(c_2)$ | — | — | < 2e−16 |

**(a) DBP** (Adj. $R^2 = 0.0298$, Dev. explained = 3.03%)

| Characteristic | Estimate ($\beta$) | 95% CI | p-value |
|---|---|---|---|
| (Intercept) | 34.89 | (34.84, 34.94) | < 2e−16 |
| $c_1$ | 0.09 | (0.06, 0.11) | 1.05e−13 |
| $c_2$ | 0.41 | (-0.02, 0.85) | 0.0639 |
| $s(c_1)$ | — | — | 0.969 |
| $s(c_2)$ | — | — | < 2e−16 |

**(b) HbA1c** (Adj. $R^2 = 0.00427$, Dev. explained = 0.443%)

| Characteristic | Estimate ($\beta$) | 95% CI | p-value |
|---|---|---|---|
| (Intercept) | 5.66 | (5.65, 5.66) | < 2e−16 |
| $c_1$ | 0.15 | (-0.04, 0.35) | 0.125 |
| $c_2$ | 0.27 | (-0.07, 0.60) | 0.124 |
| $s(c_1)$ | — | — | < 2e−16 |
| $s(c_2)$ | — | — | < 2e−16 |

**(c) WT Global** (Adj. $R^2 = 0.266$, Dev. explained = 26.7%)

| Characteristic | Estimate ($\beta$) | 95% CI | p-value |
|---|---|---|---|
| (Intercept) | 78.52 | (78.40, 78.65) | < 2e−16 |
| $c_1$ | -1.66 | (-6.24, 2.92) | 0.477 |
| $c_2$ | 14.45 | (9.30, 19.61) | 3.83e−08 |
| $s(c_1)$ | — | — | < 2e−16 |
| $s(c_2)$ | — | — | < 2e−16 |

**(d) LVEDVi** (Adj. $R^2 = 0.154$, Dev. explained = 15.4%)

| Characteristic | Estimate ($\beta$) | 95% CI | p-value |
|---|---|---|---|
| (Intercept) | 45.19 | (45.12, 45.26) | < 2e−16 |
| $c_1$ | -0.02 | (-2.56, 2.51) | 0.986467 |
| $c_2$ | 6.58 | (3.05, 10.10) | 0.000257 |
| $s(c_1)$ | — | — | < 2e−16 |
| $s(c_2)$ | — | — | < 2e−16 |

**(e) LVMi** (Adj. $R^2 = 0.289$, Dev. explained = 29.0%)

| Characteristic | Estimate ($\beta$) | 95% CI | p-value |
|---|---|---|---|
| (Intercept) | 2.35 | (2.34, 2.36) | < 2e−16 |
| $c_1$ | -0.17 | (-0.26, -0.07) | 0.000963 |
| $c_2$ | -0.18 | (-0.45, 0.08) | 0.179172 |
| $s(c_1)$ | — | — | < 2e−16 |
| $s(c_2)$ | — | — | < 2e−16 |

**(f) PDSR circum** (Adj. $R^2 = 0.0541$, Dev. explained = 5.47%)

**Supplementary Table 4. Results of statistical modeling using generalised additive models (GAMs) for continuous phenotypes**. Each subtable shows parameter estimates, 95% CI, and p-values). Adjusted $R^2$ and deviance explained are provided in captions.

| Characteristic | Estimate ($\beta$) | 95% CI | p-value |
|---|---|---|---|
| (Intercept) | -5.71 | (-5.89, -5.52) | < 2e−16 |
| $c_1$ | 0.03 | (-0.04, 0.09) | 0.455 |
| $c_2$ | 0.06 | (-0.05, 0.17) | 0.278 |
| $s(c_1)$ | — | — | 0.995 |
| $s(c_2)$ | — | — | 0.957 |

**(a) TTN Variants** (Adj. $R^2 = 2.4 \times 10^{-6}$, Dev. explained = $0.13\%$)

| Characteristic | Estimate ($\beta$) | 95% CI | p-value |
|---|---|---|---|
| (Intercept) | -6.33 | (-6.57, -6.08) | < 2e−16 |
| $c_1$ | 0.01 | (-0.08, 0.10) | 0.801 |
| $c_2$ | 0.24 | (-0.32, 0.80) | 0.400 |
| $s(c_1)$ | — | — | 0.500 |
| $s(c_2)$ | — | — | 0.229 |

**(b) HCM P/LP Variants** (Adj. $R^2 = 1.4 \times 10^{-4}$, Dev. explained = $0.70\%$)

| Variable | Estimate ($\beta$) | HR | 95% CI | p-value |
|---|---|---|---|---|
| $c_1$ | 0.031 | 1.03 | 1.00–1.06 | 0.0466 |
| $c_2$ | 0.170 | 1.19 | 1.14–1.24 | $1.4 \times 10^{-15}$ |
| **Model:** | n = 35,495; events = 625; Concordance = 0.602; LR test $p = 2 \times 10^{-15}$ | | | |
| **PH test:** | $p_{c1} = 0.45$, $p_{c2} = 0.72$, Global $p = 0.70$ (no violation) | | | |

**(c) MACE (Cox Model)**

**Supplementary Table 5. Results of statistical modeling using logistic and Cox regression for genotypic and clinical outcomes. a–b**. Logistic regression models by different genotypes (each subtable shows parameter estimates, 95% CI, and p-values; adjusted $R^2$ and deviance explained are provided in captions). **c**. Cox proportional hazards model for MACE (parameter estimates, 95% CI, and p-values; hazard ratios (HR) and proportional hazards test results are shown).

| Model | C-index | AUC (95% CI) |
|---|---|---|
| $c_1 + c_2$ | 0.6014 | 0.5773 (0.5773–0.6228) |
| $c_1 + c_2$ + covariates | 0.691 | 0.716 (0.6933–0.7386) |
| LVEF (base) | 0.5810 | 0.5804 (0.5548–0.6060) |
| LVEF + $c_1 + c_2$ | 0.6179 | **0.6181** (0.5942–0.6420) |
| LVEF + covariates | 0.7049 | 0.7252 (0.7031 – 0.7473) |
| LVEF+$c_1 + c_2$ + covariates | 0.7068 | **0.728** (0.7057 – 0.7503) |
| LVEDVi (base) | 0.5574 | 0.5532 (0.5279–0.5785) |
| LVEDVi + $c_1 + c_2$ | 0.5938 | 0.5932 (0.5693–0.6171) |
| LVEDVi + covariates | 0.7004 | 0.7189 (0.6959 – 0.7418) |
| LVEDVi + $c_1 + c_2$ + covariates | 0.6994 | 0.7206 (0.6977 – 0.7435 |
| Ell global (base) | 0.5993 | 0.5996 (0.5736–0.6257) |
| Ell global + $c_1 + c_2$ | 0.6295 | 0.6071 (0.6071–0.6567) |
| Ell global + covariates | 0.703 | 0.7248 (0.7024 – 0.7472 ) |
| Ell global + $c_1 + c_2$ + covariates | 0.7046 | 0.729 (0.7061 – 0.7518 ) |

**Supplementary Table 6.** Discriminatory performance of 5-year MACE prediction models, summarised by optimism-corrected C-index and time-dependent AUC at 5 years (95% CI). covariates included age at MRI, sex, body surface area (BSA), low-density lipoprotein cholesterol (LDL), and systolic blood pressure (SBP).

| **Variable** | $\beta$ | **HR** | **95% CI** | $p$ |
|---|---|---|---|---|
| $\mathbf{c_1}$ | -0.071 | 0.93 | 0.79–1.09 | 0.3835 |
| $\mathbf{c_2}$ | 0.314 | 1.37 | 1.12–1.68 | **0.0026** |
| Age | 0.969 | 2.63 | 2.24–3.09 | < **0.0001** |
| Sex | 0.356 | 1.43 | 1.12–1.82 | 0.0041 |
| BSA | 0.237 | 1.27 | 1.07–1.50 | 0.0061 |
| SBP | 0.103 | 1.11 | 0.98–1.25 | 0.1007 |
| LDL | 0.006 | 1.01 | 0.89–1.13 | 0.9234 |

**(a)** $c_1 + c_2$ + covariates

| Variable | $\beta$ | HR | 95% CI | $p$ |
|---|---|---|---|---|
| **LVEF** | -0.357 | 0.70 | 0.63–0.77 | < 0.0001 |
| Age | 0.962 | 2.62 | 2.23–3.07 | < 0.0001 |
| Sex | 0.332 | 1.39 | 1.10–1.76 | 0.0055 |
| BSA | 0.262 | 1.30 | 1.10–1.53 | 0.0016 |
| SBP | 0.077 | 1.08 | 0.96–1.22 | 0.2191 |
| LDL | 0.020 | 1.02 | 0.91–1.15 | 0.7397 |

**(b)** LVEF + covariates

| Variable | $\beta$ | HR | 95% CI | $p$ |
|---|---|---|---|---|
| $\mathbf{c_1}$ | -0.051 | 0.95 | 0.81–1.12 | 0.5311 |
| $\mathbf{c_2}$ | 0.313 | 1.37 | 1.11–1.68 | **0.0026** |
| **LVEF** | -0.355 | 0.70 | 0.63–0.78 | **<0.0001** |
| Age | 0.964 | 2.62 | 2.24–3.08 | **<0.0001** |
| Sex | 0.224 | 1.25 | 0.98–1.60 | 0.0736 |
| BSA | 0.211 | 1.24 | 1.04–1.46 | 0.0143 |
| SBP | 0.072 | 1.07 | 0.95–1.21 | 0.2510 |
| LDL | 0.019 | 1.02 | 0.91–1.15 | 0.7528 |

**(c)** LVEF + $c_1 + c_2$ + covariates

| Variable | $\beta$ | HR | 95% CI | $p$ |
|---|---|---|---|---|
| **LVEDVi** | 0.308 | 1.36 | 1.23–1.51 | < 0.0001 |
| Age | 1.040 | 2.83 | 2.41–3.33 | < 0.0001 |
| Sex | 0.226 | 1.25 | 0.98–1.60 | 0.0713 |
| BSA | 0.332 | 1.39 | 1.18–1.64 | < 0.0001 |
| SBP | 0.138 | 1.15 | 1.02–1.30 | 0.0271 |
| LDL | 0.026 | 1.03 | 0.91–1.15 | 0.6634 |

**(d)** LVEDVi + covariates

| Variable | $\beta$ | HR | 95% CI | $p$ |
|---|---|---|---|---|
| $\mathbf{c_1}$ | -0.069 | 0.93 | 0.79–1.10 | 0.4042 |
| $\mathbf{c_2}$ | 0.164 | 1.18 | 0.95–1.46 | 0.1315 |
| **LVEDVi** | 0.281 | 1.32 | 1.19–1.48 | < 0.0001 |
| Age | 1.039 | 2.83 | 2.40–3.33 | < **0.0001** |
| Sex | 0.192 | 1.21 | 0.94–1.56 | 0.1362 |
| BSA | 0.306 | 1.36 | 1.14–1.61 | 0.0005 |
| SBP | 0.133 | 1.14 | 1.01–1.29 | 0.0332 |
| LDL | 0.023 | 1.02 | 0.91–1.15 | 0.6957 |

**(e)** LVEDVi + $c_1 + c_2$ + covariates

| Variable | $\beta$ | HR | 95% CI | $p$ |
|---|---|---|---|---|
| **Ell Global** | 0.339 | 1.40 | 1.27–1.55 | < 0.0001 |
| Age | 0.934 | 2.55 | 2.17–2.99 | < 0.0001 |
| Sex | 0.320 | 1.38 | 1.09–1.75 | 0.0082 |
| BSA | 0.304 | 1.35 | 1.15–1.59 | 0.0003 |
| SBP | 0.068 | 1.07 | 0.95–1.21 | 0.2801 |
| LDL | -0.001 | 1.00 | 0.89–1.12 | 0.9825 |

**(f)** Ell Global + $c_1 + c_2$ + covariates

| Variable | $\beta$ | HR | 95% CI | $p$ |
|---|---|---|---|---|
| $\mathbf{c_1}$ | -0.026 | 0.97 | 0.83–1.15 | 0.7565 |
| $\mathbf{c_2}$ | 0.359 | 1.43 | 1.16–1.76 | **0.0007** |
| **Ell Global** | 0.348 | 1.42 | 1.28–1.57 | < **0.0001** |
| Age | 0.934 | 2.55 | 2.17–2.99 | < **0.0001** |
| Sex | 0.189 | 1.21 | 0.94–1.55 | 0.1393 |
| BSA | 0.244 | 1.28 | 1.08–1.51 | 0.0049 |
| SBP | 0.061 | 1.06 | 0.94–1.20 | 0.3361 |
| LDL | -0.002 | 1.00 | 0.89–1.12 | 0.9722 |

**(g)** Ell Global + $c_1 + c_2$ + covariates

**Supplementary Table 7. Cox proportional hazards models for MACE.** Comparison of multivariable Cox models including latent motion coordinates ($c_1$, $c_2$), conventional CMR biomarkers (LVEF,LVEDVi,Ell Global), and their combination. Covariates included age at MRI, sex, body surface area (BSA), systolic blood pressure (SBP), and low-density lipoprotein cholesterol (LDL). Hazard ratios (HRs) correspond to the interquartile-range contrast reported by `rms::summary`.

| Category | Metric | Value |
|---|---|---|
| **Cross-Validation** | $R^2$ | 0.9948 |
| | RMSE | 0.1567 |
| | MAE | 0.0471 |
| **Training vs Test** | Trustworthiness (Train 32d–2d) | 0.8801 |
| | Trustworthiness (Test 32d–2d) | 0.8773 |
| | Spearman (Summary Stats) | 1.0000 |
| **Training vs Test tree axes** | $c_1$ Spearman (Summary) | 1.0000 |
| | $c_2$ Spearman (Summary) | 1.0000 |
| | $c_1$ KS Test p-value | p=0.002758 (D=0.0130) |
| | $c_2$ KS Test p-value | p=0.000000 (D=0.0214) |
| | $c_1$ Wasserstein Distance | 0.0377 |
| | $c_2$ Wasserstein Distance | 0.0458 |
| | $c_1$ Jensen–Shannon Divergence | 0.00328 |
| | $c_2$ Jensen–Shannon Divergence | 0.00345 |
| | $c_1$ Cohen's d (Effect Size) | 0.00230 |
| | $c_2$ Cohen's d (Effect Size) | -0.00624 |
| | $c_1$ overlapping coefficient (OVL) | 0.8921 |
| | $c_2$ overlapping coefficient (OVL) | 0.8744 |

**Supplementary Table 8. Summary of DDRTree model evaluation and generalisation results.** The model was trained on 36,856 participants and validated on a unseen independent set of 41,257 UK Biobank participants. Higher values of Spearman correlation, and overlap coefficient (OVL) indicate greater consistency between training and test cohorts, whereas lower values of Kolmogorov–Smirnov (KS) statistic, Wasserstein distance, Jensen–Shannon divergence, and Cohen's d denote smaller distributional differences. The results indicate that the test and training embeddings are highly similar, demonstrating stable and generalisable performance.

| Endpoint | Definition | UKB data-field ID(s) | ICD-10 Code(s) |
|---|---|---|---|
| *Patient Characteristics* | | | |
| Ethnicity | Self-reported Ethnicity | 21000 | - |
| Sex | Biological Sex at Birth | 31 | - |
| Age | Age at Imaging Visit | 12697 | - |
| SBP | Systolic Blood Pressure | 4080, 93 | - |
| DBP | Diastolic Blood Pressure | 4079, 94 | - |
| BSA | Body Surface Area | Derived from 21002 (weight), 50 (height) | - |
| Hypertension | Diagnosed Hypertension | 131286, 6150, 41202, 41204 | I10-I15 |
| Valvular Disease | Any Valvular Pathology | 41202, 41204 | I05 |
| HbA1c | - | 30750 | - |
| *Clinical Outcomes* | | | |
| All-cause Mortality | Death from any Cause | 40000 (date), 40001 / 40002 (cause) | Any |
| HF | First HF / Cardiomyopathy Record | 131298, 41202, 41204, 42018 | I11, I13, I50, I42 |
| Atrial fibrillation or flutter | First Arrhythmia Event | 131424, 41202, 41204, 42018 | I47-I49 |
| AV conduction disorder | First diagnosis | - | I44, I45 |
| Stroke | First Ischaemic or Haemorrhagic Stroke | 131040, 41202, 41204, 42018 | I60-I64 |
| Cardiac arrest | First Sudden Cardiac arrest / Ventricular stand-still | 41202, 41204, 40000 | I46, I49.0 |
| Angina | First diagnosis | - | I20 |
| Ischaemic heart disease | First diagnosis | 42006 | I20-I25 |
| MI | First diagnosis | 42006 | I21-I23 |
| MACE | Composite of Stroke, MI, HF, Arrest, and cardiovascular mortality | 131040, 131298, 131424, 41202, 41204, 42018 | I42, I46, I47-I49, I48, I50, I60-I64 |
| Amyloidosys | First diagnosis | - | E85 |
| Sarcoidosis | First diagnosis | - | D86 |
| Aortic valve disease | First diagnosis | - | I06, I35 |
| DCM | First diagnosis | - | I42.0, I42.6, I42.7 |
| HCM | First diagnosis | - | I42.1, I42.2 |
| *CMR-Derived Variables* | | | |
| CMR variables | All Variables derived from cardiac images | 20207 | - |

**Supplementary Table 9. UK Biobank study variable definitions, data-field IDs, and ICD-10 codes for cardiovascular outcomes analysis.** AV, atrioventricular; BSA, body surface area; CMR, cardiac magnetic resonance; DBP, diastolic blood pressure; DCM, dilated cardiomyopathy; HbA1c, glycated haemoglobin; HCM, hypertrophic cardiomyopathy; HF, heart failure; ICD-10, International Classification of Diseases, 10th Revision; IHD, ischaemic heart disease; MACE, major adverse cardiovascular event; MI, myocardial infarction; SBP, systolic blood pressure; UKB, UK Biobank.

| Exposure factor | UKB data-field ID(s) |
|---|---|
| **Household** | |
| Own or rent | 680 |
| Household income | 737 |
| **Electronics use** | |
| Weekly mobile phone use | 1120 |
| **Sleep** | |
| Sleep difficulties | 1200 |
| Snoring | 1210 |
| Sleep duration | derived |
| **Smoking** | |
| Pack years | 20161 |
| Smoking status | 20116 |
| **Alcohol** | |
| Alcohol drinking frequency | 1558 |
| **Diet** | |
| Processed meat | 1349 |
| Cheese | 1408 |
| Coffee | 1498 |
| Red meat | derived |
| **Physical environment** | |
| Nitric dioxide (2005) | 24016 |
| Nitric dioxide (2006) | 24017 |
| Nitric dioxide (2007) | 24018 |
| Nitric dioxide (2010) | 24003 |
| Nitric oxides (2010) | 24004 |
| $PM_{2.5}$ (2010) | 24004 |

**Supplementary Table 10.** Data dictionary of exposure variables with UK Biobank field IDs. The 19 field IDs listed were used to derive the 25 exposure factors included in the analysis as described in the Supplementary Method. PM, particulate matter with a diameter of 2.5 $\mu$m

| Outcome | Cases / Events | Controls |
|---|---|---|
| Hypertension | 11,379 | 25,477 |
| Ischemic heart disease | 3,328 | 33,528 |
| Myocardial infarction | 1,735 | 35,121 |
| Angina | 1,727 | 35,129 |
| Atrial fibrillation and flutter | 1,665 | 35,191 |
| Stroke | 1,873 | 34,983 |
| Heart failure | 469 | 36,387 |
| Aortic valve disease | 288 | 36,568 |
| Atrioventricular conduction disorders | 723 | 36,133 |
| Dilated Cardiomyopathy | 43 | 36,813 |
| Amyloidosis | 21 | 36,835 |
| Sarcoidosis | 105 | 36,751 |
| Major adverse cardiac events | 625 | 35,495 |

**Supplementary Table 11. Summary of cardiovascular outcomes analyzed among 36,856 UK Biobank participants in the training set.** Major adverse cardiac events is a composite outcome comprising stroke, myocardial infarction, heart failure, cardiac arrest, and cardiovascular mortality.

| Latent | Dev Min | Dev Max | Remaining UKB Min | Remaining UKB Max | Outside Dev Range(%) |
|---|---|---|---|---|---|
| $z_{12}$ | -0.03186 | 2.64236 | -0.26750 | 2.672071 | 0.121233 |
| $z_{5}$ | -0.23077 | 0.120789 | -0.27706 | 0.15881 | 0.069887 |
| $z_{18}$ | -0.27414 | 0.438472 | -0.28810 | 0.537932 | 0.051346 |
| $z_{7}$ | -0.13986 | 0.090862 | -0.15926 | 0.096616 | 0.049919 |
| $z_{15}$ | -0.15974 | 0.084325 | -0.17169 | 0.103745 | 0.047067 |
| $z_{24}$ | -0.36433 | 0.141856 | -0.40929 | 0.151214 | 0.045641 |
| $z_{17}$ | -0.16587 | 2.598466 | -0.50369 | 2.792556 | 0.037083 |
| $z_{4}$ | -0.10709 | 0.076895 | -0.11667 | 0.080575 | 0.037083 |
| $z_{27}$ | -0.45348 | 0.749456 | -0.46652 | 0.858901 | 0.035657 |
| $z_{13}$ | -0.18210 | 0.185488 | -0.17928 | 0.213879 | 0.034230 |
| $z_{14}$ | -0.03680 | 0.482962 | -0.04832 | 0.521758 | 0.028525 |
| $z_{25}$ | -0.13149 | 0.223924 | -0.15406 | 0.259848 | 0.028525 |
| $z_{21}$ | -0.19584 | 0.406962 | -0.19846 | 0.452962 | 0.025673 |
| $z_{2}$ | -0.20646 | 0.183019 | -0.19495 | 0.19880 | 0.021394 |
| $z_{26}$ | -2.58707 | 0.796805 | -2.95042 | 1.211731 | 0.019968 |
| $z_{16}$ | -1.05550 | 2.509423 | -1.03377 | 2.642020 | 0.017115 |
| $z_{19}$ | 0.291606 | 2.892362 | 0.230368 | 2.990627 | 0.017115 |
| $z_{9}$ | -0.11484 | 0.142137 | -0.13076 | 0.154905 | 0.014263 |
| $z_{6}$ | -0.03913 | 0.395601 | -0.04128 | 0.410482 | 0.012836 |
| $z_{3}$ | -0.22314 | 0.218731 | -0.20233 | 0.228312 | 0.009984 |
| $z_{1}$ | -0.07620 | 0.298841 | -0.08166 | 0.315138 | 0.008558 |
| $z_{11}$ | -0.79438 | 1.128936 | -0.75239 | 1.377347 | 0.008558 |
| $z_{10}$ | -0.28528 | 0.202190 | -0.26612 | 0.228422 | 0.007131 |
| $z_{30}$ | -0.73467 | 1.072535 | -0.92412 | 1.096994 | 0.007131 |
| $z_{23}$ | -0.28255 | 0.243064 | -0.32659 | 0.243213 | 0.005705 |
| $z_{32}$ | -0.12167 | 0.224652 | -0.15219 | 0.231876 | 0.005705 |
| $z_{31}$ | -0.11415 | 0.197422 | -0.10230 | 0.210719 | 0.004279 |
| $z_{8}$ | -0.10599 | 0.210133 | -0.10776 | 0.202992 | 0.002853 |
| $z_{28}$ | -0.05896 | 0.191648 | -0.05937 | 0.217253 | 0.002853 |
| $z_{29}$ | -0.17057 | 0.219159 | -0.17688 | 0.203624 | 0.002853 |
| $z_{20}$ | -0.21175 | 0.089094 | -0.20259 | 0.100695 | 0.001426 |
| $z_{22}$ | -0.21061 | 0.392082 | -0.17518 | 0.391727 | 0.000000 |

**Supplementary Table 12. Comparison of latent variable ranges derived from *Cardio4D-VAE* between the model-development cohort (training/validation dataset,8000 sampels) and remaining UK Biobank cohorts (70,113 samples).** For each latent dimension, the minimum and maximum values observed in the model-development cohort are compared with those observed in the remaining UK Biobank participants, together with the percentage of participants whose latent values fall outside the range observed during model development.

### Supplemental Movies

**Movie S1: Spatiotemporal left ventricular displacement in TTN variant carriers.** Vertex-wise displacement maps comparing TTN variant carriers with non-carriers across the cardiac cycle. Warm colors indicate greater outward displacement relative to controls; cool colors indicate reduced displacement. Animated version of Figure 8a.

**Movie S2: Spatiotemporal left ventricular displacement in HCM P/LP sarcomeric variant carriers.** Vertex-wise displacement maps comparing HCM P/LP carriers with non-carriers across the cardiac cycle. Warm colors indicate greater outward displacement relative to controls; cool colors indicate reduced displacement. Animated version of Figure 8a.

**Movie S3: Spatiotemporal left ventricular velocity differences in TTN variant carriers.** Vertex-wise signed velocity difference maps comparing TTN carriers with non-carriers across the cardiac cycle. Warm colors indicate regions where carriers exhibit faster myocardial motion than controls, and cool colors indicate slower motion relative to controls (mm/frame). Animated version of Figure 8b.

**Movie S4: Spatiotemporal left ventricular velocity differences in HCM P/LP sarcomeric variant carriers.** Vertex-wise signed velocity difference maps comparing HCM P/LP carriers with non-carriers across the cardiac cycle. Warm colors indicate regions where carriers exhibit faster myocardial motion than controls, and cool colors indicate slower motion relative to controls (mm/frame). Animated version of Figure 8b.

### Supplemental Data

**Data S1: LD-clumped loci and lead variants for conventional and latent traits.** LD-clumped locus-trait pairs from GWAS of LVEF, Ell global, LVEDVi, and 32 latent LV motion traits, reporting GRCh38 coordinates, locus span, sentinel variant (rsID) and P-value, and the number of traits with an independent lead variant at each locus.

**Data S2: SNP-based heritability estimates for conventional and latent traits.** Summary statistics from linkage disequilibrium score regression (LDSC) quantifying the common variant heritability of all traits. Columns report the heritability point estimates and standard errors (`h2`, `h2_se`) derived from `N_SNP_used` variants. Diagnostic metrics for assessing population stratification and cryptic relatedness include the LDSC intercept (`intercept`, `intercept_se`) and the attenuation ratio. Model fit and genomic inflation are further quantified by the genomic control inflation factor (`lambda_GC`) and mean $\chi^2$ statistic.

**Data S3: Grouping of baselineLD v2.2 annotations into canonical regulatory families for S-LDSC.** Reference table mapping specific baselineLD v2.2 annotation tracks to canonical regulatory families and broader functional categories used in stratified LD score regression (S-LDSC). Columns list the track name, collapsed analysis group, and biological class. Flags indicate tracks derived from 500-bp flanking windows and distinguish families selected for enrichment visualisation from those used solely as baseline covariates.

**Data S4: FUMA-mapped genes at conventional and latent trait loci.** Genes mapped to trait loci ($z_1$–$z_{32}$, LVEF, Ell global, LVEDVi) by positional, eQTL, or chromatin-interaction evidence, with gene identifiers, GRCh37/GRCh38 coordinates, constraint metrics (pLI, ncRVIS), mapping evidence indicators, and overlap with loci for conventional cardiac traits.

**Data S5: Transcriptome-wide association statistics for conventional and latent LV motion traits.** TWAS results from FUSION using GTEx v8 expression models for Heart Left Ventricle and Heart Atrial Appendage, listing gene-tissue pairs with genomic coordinates, tissue model, TWAS $Z$-score, and nominal $P$-value.

**Data S6: Data package for the finemapped cell-state analysis.**
